# Integrated Cardiac and Circulating N-glycan Signatures Reflect Atrial Remodeling in Patients with Atrial Fibrillation

**DOI:** 10.64898/2026.01.29.26345171

**Authors:** Chi Him Kendrick Yiu, Jack Cheeseman, Georgia Elgood-Hunt, Chenhan Sam Ma, Abhirup Banerjee, Lucia M. Moreira, Aaron M. Johnston, Neelam Mehta, Kathryn Cox, Tim R. Betts, Kim Rajappan, Matthew Ginks, Michala Pedersen, Yaver Bashir, Rohan Wijesurendra, Rana Sayeed, George Krasopoulos, Vivek Srivastava, Antonios Kouliouros, Daniel I. R. Spencer, Svetlana Reilly

## Abstract

**Background:** Atrial adverse remodeling drives the maintenance and progression of atrial fibrillation (AF) through electrical and structural myocardial changes, often accompanied by inflammation. Circulating N-glycans are emerging as biomarkers in inflammatory diseases, yet their role in AF remains undefined.

**Methods:** We profiled the serum N-glycome of 138 patients with AF, non-AF arrhythmias, or sinus rhythm (SR) controls from peripheral venous (PV) and coronary sinus (CS) samples using hydrophilic interaction liquid chromatography coupled with high-resolution mass spectrometry. Glycan traits associated with AF were identified via logistic regression adjusted for clinical risk factors. Multivariate glycan scores were derived from PV and CS datasets using LASSO regression. In a subset (N=37), plasma proteome profiling was performed with the Olink Reveal panel.

**Results:** Sixty-two glycan peaks were detected; 27 in PV and 8 in CS serum differed significantly between AF and controls. PV and CS glycan scores accurately classified AF, with the PV score correlating with 11 plasma proteins linked to structural remodeling and thrombo-inflammatory processes. The most abundant glycan, A2G2S2 (peak 30), was associated with higher odds of AF after adjusting for confounders (OR 2.22 [95% CI: 1.40–3.75], P = 0.001). CS A2G2S2 correlated with C-reactive protein (R = 0.432, P = 0.0275) and was elevated in patients with left atrial enlargement (P = 0.0354), but unchanged in those with impaired left ventricular ejection fraction or hypertrophy.

**Conclusion:** Integrated profiling of peripheral and cardiac serum identifies novel N-glycosylation signatures in AF. Specific cardiac and circulating N-glycan signatures, including A2G2S2, are associated with AF and reflect inflammation-driven atrial remodeling, highlighting potential mechanistic pathways and biomarker applications.

## Introduction

Atrial fibrillation (AF) is the most common sustained arrhythmia, with a global prevalence of more than 59 million cases (1). AF increases the risk of mortality, primarily due to serious complications, such as stroke and heart failure (2). Moreover, patients with AF are 39% more likely to develop dementia and experience poorer quality of life (3,4). Multiple factors, including age, AF duration, inflammation, obesity, underlying heart disease, and prior surgery, promote pathological atrial remodeling in AF (5,6). This remodeling, represented by electrical and structural (fibrotic) changes in the atrial myocardium, promotes AF maintenance and progression, and hampers arrhythmia treatment. Thus, a better understanding of AF pathogenesis, through identification of novel molecular markers, may benefit the development of new clinical strategies to manage AF and improve its risk prediction.

Complex mechanisms driving the arrhythmogenic substrate include atrial fibrosis, oxidative stress, ion channel alterations, and inflammatory or cell-to-cell signaling processes (7). Atrial fibrosis, characterised by increased extracellular matrix (ECM) deposition in the atria (8), negatively impacts on the success of AF treatments with pharmacological strategies or catheter ablation (9). Thus, better understanding of the mechanisms contributing to fibrosis may aid the development of more successful therapeutics for AF. Besides known mediators, such as transforming growth factor-beta (TGF-β), angiotensin II–renin axis, and galectin-3, N-glycosylation has emerged as a key post-translational modification of ECM proteins, potentially driving atrial fibrotic remodeling (10–12).

N-glycosylation, a common post-translational modification, occurs on most secreted proteins and regulates critical processes including protein folding, intercellular communication, cell adhesion, and immune cell migration and activation (13,14). Changes in plasma and serum N-glycan concentrations have been linked to diseases including diabetes, cancer, liver fibrosis, and cardiovascular diseases (15–20), with sialylation levels often outperforming standard inflammatory biomarkers such as high-sensitivity C-reactive protein (CRP) (21,22). Alterations in *N*-glycosylation patterns have been found in CVD pathologies (e.g. myocardial infarction, stroke, coronary revascularisation, unstable angina requiring hospitalisation, death, resuscitation after cardiac arrest), as defined in the JUPITER and TNT trials (23). Furthermore, agalactosylation and asialylation of IgG glycans were shown to be associated with increased risk of CVD (23). In relation to AF, a recent study profiled plasma and IgG *N*-glycosylation levels, patterns and traits in patients with AF and found distinct *N*-glycan signatures attributed to the AF type, AF recurrence after ablation, and stroke risk (24). These findings indicate that *N*-glycans may emerge as valuable indications for AF and AF-associated remodeling, which necessitates their further exploration and validation in clinical cohorts.

Despite evidence for N-glycans in cardiovascular pathology, their role in AF and relationship to atrial remodeling remain unknown. To address this gap, we performed integrated profiling of peripheral and cardiac serum N-glycans in AF patients, aiming to identify novel molecular signatures linked to inflammation-driven atrial remodeling and potential biomarker development

## Methods

### Study design and patient population

This study was approved by the South Central-Berkshire B Research Ethics Committee (UK, 18/SC/0404 and 18/SC/0304) at John Radcliffe Hospital, Oxford. Patients who underwent elective cardiac surgery (coronary artery bypass graft, valve repair/replacement) or catheter ablation were recruited prospectively and all patients provided written informed consent before enrolment. Patients with AF (paroxysmal or persistent) were considered as the study group, which were compared with two control groups: 1) patients in sinus rhythm (SR), and 2) a non-AF group consisting of patients with supraventricular tachycardias (SVTs) (without AF) and atrial flutter. The latter (non-AF) patients were scheduled for routine electrophysiological (EP) study. Diagnosis of AF and arrhythmic events were determined according to the electronic patient records, the EP study and electrocardiogram (ECG) data.

Inclusion criteria were 1) males and females aged between 18 and 85 years old, 2) undergoing cardiac surgery for any heart condition for the first time in last six months or undergoing catheter ablation for AF or routine EP study (non-AF group), and 3) who were willing and able to give informed consent for participation. 138 patients, of whom 62 were in AF, 52 controls in SR and 20 non-AF controls were recruited between November 2018 and May 2024 (**Table 2-3**).

### Patient data collection

Demographic and clinical characteristics were collected from the NHS electronic patient records by the study team of qualified researchers and clinicians. Data included demographic characteristics such as age, sex, body mass index (BMI); medical history including hypertension, ischemic heart disease (IHD), stroke or transient ischaemic attack (TIA), myocardial infarction, peripheral arterial disease (PAD), heart failure, chronic obstructive pulmonary disease (COPD), diabetes, chronic kidney disease, thyroid disease and duration of AF, and medications. Biochemical measurements were recorded when available and included haemoglobin, white blood count, urea, creatinine, CRP, estimated glomerular filtration rate (eGFR), iron, transferrin level and saturation, and ferritin. Cardiac functional parameters, such as ECG measurements (heart rate, QT interval, QRS interval, PR interval), and echocardiography parameters (indexed left atrial volume, left atrial dimension, indexed left ventricular mass, left ventricular ejection fraction [LVEF]) were also collected.

### Blood collection and processing

In the cardiac surgery patient cohort, peripheral venous (PV) blood samples were taken via intravenous access. All blood samples were collected by the clinical team at three timepoints: the day of cardiac surgery, day 2 and day 4 after surgery. In the catheter cardiac ablation patient cohort, coronary sinus (CS) blood samples were taken through transfemoral access of intra-cardiac sites within the CS under ultrasound and fluoroscopy guidance using a diagnostic Amplatz AL1 catheter or Agilis sheath. The PV blood samples were collected from the femoral veins before and 2 hours post-ablation, and from the antecubital veins at two follow up visits 3 months and 12 months after the procedure. PV blood samples were collected using sterile syringes by the study’s clinical team.

For *N*-glycome profiling, PV and CS blood samples were immediately transferred to BD Vacutainer Yellow SST II advance tubes, left to clot at room temperature for 1 hour and then centrifuged at 3000 rpm for 15 minutes at room temperature. Upon layer separation, serum was collected from the top layer and stored at -80°C until use in experiments. For Olink assay, PV blood samples were immediately transferred to BD Vacutainer Citrate tubes, left to clot at room temperature for 1 hour and then centrifuged at 3000 rpm for 15 minutes at room temperature. Upon layer separation, plasma was collected from the top layer and stored at -80°C until use in experiments.

### Serum sample processing for N-glycome profiling

To each well of a skirted 96-well plate (4titude, 180 µL volume) 5 µL of either serum samples, plasma standards (Quadratech Diagnostics Ltd, UK) or IgG standards (Sigma Aldrich, UK) was added. The 96-well plate was transferred to a liquid handling robot (Hamilton Starlet) for all subsequent steps.

The volume was topped up to 9 µL by addition of 4 µL water. 1 µL of denaturation buffer (NEB, UK) was then added to each sample. The samples were vortexed and briefly centrifuged before incubation at 100^°^C for 10 minutes. After cooling to room temperature, the following was added to each sample: 5 µL water, 1 µL NP-40 (NEB, UK), 2 µL reaction buffer (NEB, UK) and 2 µL PNGase-F solution (P0709L, NEB, UK). The samples were vortexed and briefly centrifuged before incubation at 37^°^C for 16 hours. The resulting sample mixtures were dried using a vacuum centrifuge (Thermo Scientific) for 2 hours. The glycans were converted to aldoses by the addition of 20 µL 1% formic acid solution, vortexed, and briefly centrifuged before incubation at room temperature for 45 minutes. The samples were filtered through a LudgerClean 96-well protein binding membrane plate (LC-PBM-96, Ludger, UK) with water (2 x 100 µL) to remove excess protein. Subsequently, the resulting 200 µL solutions were transferred to a non-skirted 96-well plate (4titude, 300 µL volume) and dried down in a vacuum centrifuge for 9 hours. The dried glycans were subjected to fluorescent labelling by reductive amination with 20 µL procainamide labelling solution (LT-KPROC-96, Ludger, UK), vortexed, and briefly centrifuged before incubation for 1 hour at 65°C. The samples were then mixed with 80 µL acetonitrile before transferring to a LudgerClean procainamide clean-up plate where they were washed with acetonitrile (3 x 100 µL) to remove excess labelling solution. The labelled glycans were recovered by washing with water (2 x 100 µL). For liquid chromatography tandem mass spectrometry (LC-MS/MS) analysis, 100 µL of each sample was added to 300 µL of acetonitrile. System suitability standards (procainamide labelled plasma and IgG, and blanks) were prepared by adding 25 µL of each standard or water to 75 µL acetonitrile. Each standard or sample was analysed by hydrophilic interaction liquid chromatography with high-resolution mass spectrometry (HILIC-HRMS) with florescence detection. 20 μL of each standard or sample was injected into an ACQUITY UPLC BEH-Glycan 1.7μm, 2.1 mm×150 mm column (Waters) at 40°C on a Thermo Scientific Vanquish ultra-high performance liquid chromatography (UHPLC) instrument with a fluorescence detector (λex = 310 nm, λem = 370 nm), attached to a Thermo Scientific Orbitrap Exploris 120 mass spectrometer. The chromatography conditions were solvent A 50 mM ammonium formate pH 4.4 made from LudgerSep N-Buffer, and solvent B acetonitrile. Gradient conditions were 0 to 53.5 min, 76 to 51% B, 0.4 mL/min; 53.5 to 55.5 min, 51% to 0% B, 0.4 mL/min to 0.2 mL/min; 55.5 to 57.5 min,0% B at a flow rate of 0.2 mL/min; 57.5 to 59.5 min, 0 to 76% B, 0.2mL/min; 59.5 to 65.5 min, 76% B, 0.2 mL/min; 65.5 to 66.5 min,76% B, 0.2 mL/min to 0.4 mL/min; 66.5 to 70.0 min, 76% B, 0.4 mL/min. The Exploris 120 settings were source temperature, 300^°^C; ion transfer tube temperature, 325^°^C; sheath gas, 50 arbitrary units (Arb); auxiliary gas, 10 Arb; sweepgas, 1 Arb; capillary voltage, 3500 V; positive ion mode; mass range scanned, 500-2500 m/z; resolution, 120000, data dependent acquisition mode; RF level, 70%.

### N-glycome data processing

The fluorescence data was integrated and quantified using HappyTools (version 0.1-beta1, build 190115a) as previously described (25). The relative abundance of each glycan trait in a serum sample was expressed as the ratio of the specific peak area over the total area of all peaks. In addition to the sixty-two detected glycan traits in human serum, twelve *N*-glycosylation traits were derived and listed as follows: oligomannose (O), no galactosylation (G0), diantennary galactosylation (G2), bisected galactosylation (B), triantennary galactosylation (G3), tetrantennary galactosylation (G4), diantennary sialylation (S2), bisected sialylation (SB), triantennary sialylation (S3), tetrantennary sialylation (S4), antenna fucosylation (AFu) and core fucosylation (CFu). Derived traits were expressed as a sum of relative areas of glycans that share common biochemical structures and the formulas of calculating these derived traits were provided in **Table 1**.

**Table 1:** Derived *N* -glycosylation traits in human serum.

| Derived traits | Formula |
| --- | --- |
| O: Oligomannose | Peak 5+Peak 13+Peak 21 |
| G0: No galactosylation | (Peak 1 / 2)+Peak 2+Peak 3+Peak 6 |
| G2: Diantennary galactosylation | (Peak 7+Peak 8+Peak 10+ Peak 15+Peak 16+Peak 20+Peak 22) / 2 + Peak 14+Peak 17+Peak 24+Peak 26+ Peak 27+Peak 29+Peak 30+Peak 32+Peak 33+Peak 36 |
| B: Bisected galactosylation | (Peak 9+Peak 12+Peak 19) / 2 + Peak 18+Peak 23+Peak 25+Peak 31+Peak 34 |
| G3: Triantennary galactosylation | Peak 35+Peak 37+Peak 38+Peak 39+Peak 40+Peak 41+Peak 42+Peak 43+Peak 44+Peak 45+Peak 55+Peak 46+Peak 47+Peak 48+Peak 49 |
| G4: Tetrantennary galactosylation | Peak 62+Peak 61+Peak 60+Peak 59+Peak 58+Peak 57+Peak 56+Peak 54+Peak 53+Peak 52+Peak 51+Peak 50 |
| S2: Diantennary sialylation | (Peak 15+Peak 16+Peak 24+Peak 27+Peak 32+Peak 36+Peak 22) / 2 + Peak 20+Peak 26+Peak 29+Peak 30+Peak 33 |
| SB: Bisected sialylation | (Peak 23+Peak 25) / 2 + Peak 31+Peak 34+Peak 19 |
| S3: Triantennary sialylation | (Peak 35+Peak 37+Peak 38+Peak 39+Peak 40+Peak 49) * 2 / 3 + Peak 41+Peak 42+Peak 43+Peak 44+Peak 45+Peak 55+Peak 46+Peak 47+Peak 48+Peak 50+Peak 51+Peak 52+Peak 53 |
| S4: Tetrantennary sialylation | Peak 62+Peak 61+Peak 60+Peak 59+Peak 52+Peak 53+Peak 54+Peak 56+Peak 57+Peak 58 |
| AFu: Antenna fucosylation | Peak 48+Peak 62 |
| CFu: Core fucosylation | Peak 1+Peak 3+Peak 6+Peak 10+Peak 11+Peak 12+Peak 17+Peak 18+Peak 20+Peak 24+Peak 27+Peak 25+Peak 29+Peak 33+Peak 34+Peak 39+Peak 40+Peak 43+Peak 44+Peak 46+Peak 47+Peak 48+Peak 59+Peak 60+Peak 61+Peak 62 |

The mass spectrometry data for *N*-glycans was analysed using Thermo Scientific Freestyle software (version 1.8) and the structures were manually assigned. Assignments were made based on exact mass, and the accuracy of the actual mass compared to the theoretical mass being below 20ppm, fragmentation data and retention order. Further to this, reference was made to previously reported glycan structures found in plasma and serum (21,26). *N*-glycan compositions were illustrated to the Consortium for Functional Glycomics (CFG) notation (27): *N*-acetylglucosamine (N; blue square), fucose (F; red triangle), galactose (H; yellow circle), mannose (H; green circle), *N*-acetylneuraminic acid (S; purple diamond).

### Statistical analysis

Statistical analyses and data visualisation were performed using GraphPad Prism 10.4.1 and R version 4.3.2. Batch correction was conducted using the analysis of covariance (ANCOVA) framework method to minimise technical variations between batches (28). Normal distribution was determined by performing a Shapiro-Wilk test. For two-group comparisons, normally distributed data were analysed by unpaired t-test or Welch’s t-test (for two groups with unequal variances), and non-parametric data were analysed by Mann-Whitney test. In paired samples, normally distribute data were analysed by paired t-test (for two-group comparisons) and repeated-measures one-way analysis of variance (ANOVA) with post-hoc Tukey test (multiple-group comparisons). Correlation between two continuous variables were tested using Pearson correlation test for normally distributed data, and Spearman correlation test for non-normally distributed data. Differences between two categorical variables were tested using Fisher’s exact test. The statistical significance was reached when *P*-value < 0.05. Receiver Operating Characteristic (ROC) curve analysis was performed with a five-fold cross-validation approach to give an averaged area under the ROC curve (AUROC) value. Analyses on binary logistic regression were done in R using the package gtsummary (v2.0.4) and finalfit (v1.0.8). Glycan values were normalised to z-scores for regression analyses, and thus ORs are expressed as a function of a 1 standard deviation change. Check for multicollinearity in regression models was completed using the vif() function from the car package (v3.1.2).

### N-glycan scores

The multivariate *N*-glycan scores for AF were analysed and generated in R version 4.4.1. Prior to modelling, all glycan peak intensities were standardised using z-score transformation. Separate models were developed for the PV and CS datasets. Two levels of prediction tasks were examined: 1) binary classification of AF vs. no AF, and 2) multiclass classification of AF subtypes: no AF, paroxysmal AF and persistent AF.

To identify informative glycans while controlling for collinearity, least absolute shrinkage and selection operator (LASSO) regression was applied with unpenalised clinical covariates. In the binary classification, LASSO logistic regression was used with age, sex, BMI, hypertension and IHD (for PV model) or age, sex and BMI (for CS model) specified as unpenalised variables. In the multiclass classification, a multinomial LASSO model was fitted using the same covariate sets. The optimal penalty parameter (λ) was selected via ten-fold cross-validation, and glycans with non-zero coefficients at λ_min were deemed selected.

Predicted probabilities were generated for: *clinical model* including only the fixed covariates, *glycan score model* based solely on the selected glycans, and *combined model* incorporating both. Model discrimination was quantified using the AUROC for binary tasks and the macro-averaged AUROC for multiclass outcomes.

To evaluate the incremental predictive value of adding glycans to the clinical model, the continuous net reclassification improvement (NRI) and integrated discrimination improvement (IDI) were calculated. The NRI quantified the change in directional risk classification between the combined and clinical models, while the IDI measured the mean improvement in separation between predicted probabilities for AF and non-AF cases. Overall predictive accuracy was measured by the Brier score, defined as the mean squared error between predicted and observed outcomes. For interpretability, model coefficients were visualised using bar plots, where standardised LASSO coefficients were displayed by magnitude and direction (positive or negative log-odds).

### Plasma sample and data processing for Olink

Plasma proteomes in baseline samples were profiled using the Olink Reveal panel with the proximity extension assay (PEA) technology (Olink Proteomics AB, Uppsala, Sweden). In brief, 40 μl per well of plasma was loaded to the wells in a clean 96-well PCR-plate with full skirt (#AB0800, ThermoFisher Scientific, US). Sealed plate was transferred on dry ice to the Multiomics Technology Platform Group (Oxford, UK), followed by the generation and processing of sequencing data. Next generation sequencing (NGS) was then performed on the plasma samples using Illumina’s NovaSeq 6000 platform. Finally, protein expression was measured and quantified as the normalised protein expression (NPX) values from Olink.

## Results

### Patient characteristics

Study workflow is presented in **Supplementary Fig. 1**. In this study we recruited 138 patients who were assigned to 3 groups: AF (N=66), non-AF (N=20), or sinus rhythm (SR) (N=52) controls for subsequent blood collection and analysis of *N*-glycosylation of serum proteins. The baseline characteristics between SR and AF patients are summarised in **Table 2**. While there was no difference in age, sex, BMI, diabetes, heart failure, stroke or TIA, PAD, COPD and thyroid disease between the groups, a larger proportion of patients in SR group suffered with comorbidities, including hypertension, myocardial infarction, IHD, and chronic kidney disease. Similarly, administration of antiplatelets, nitrates, statins, and angiotensin-converting enzyme inhibitor (ACEi) or angiotensin II receptor blockers (ARBs) were also significantly higher in the SR patients. Expectedly, patients with AF were more frequently prescribed β-blockers, amiodarone, flecainide, anticoagulants, digoxin and diuretics.

**Table 2:**
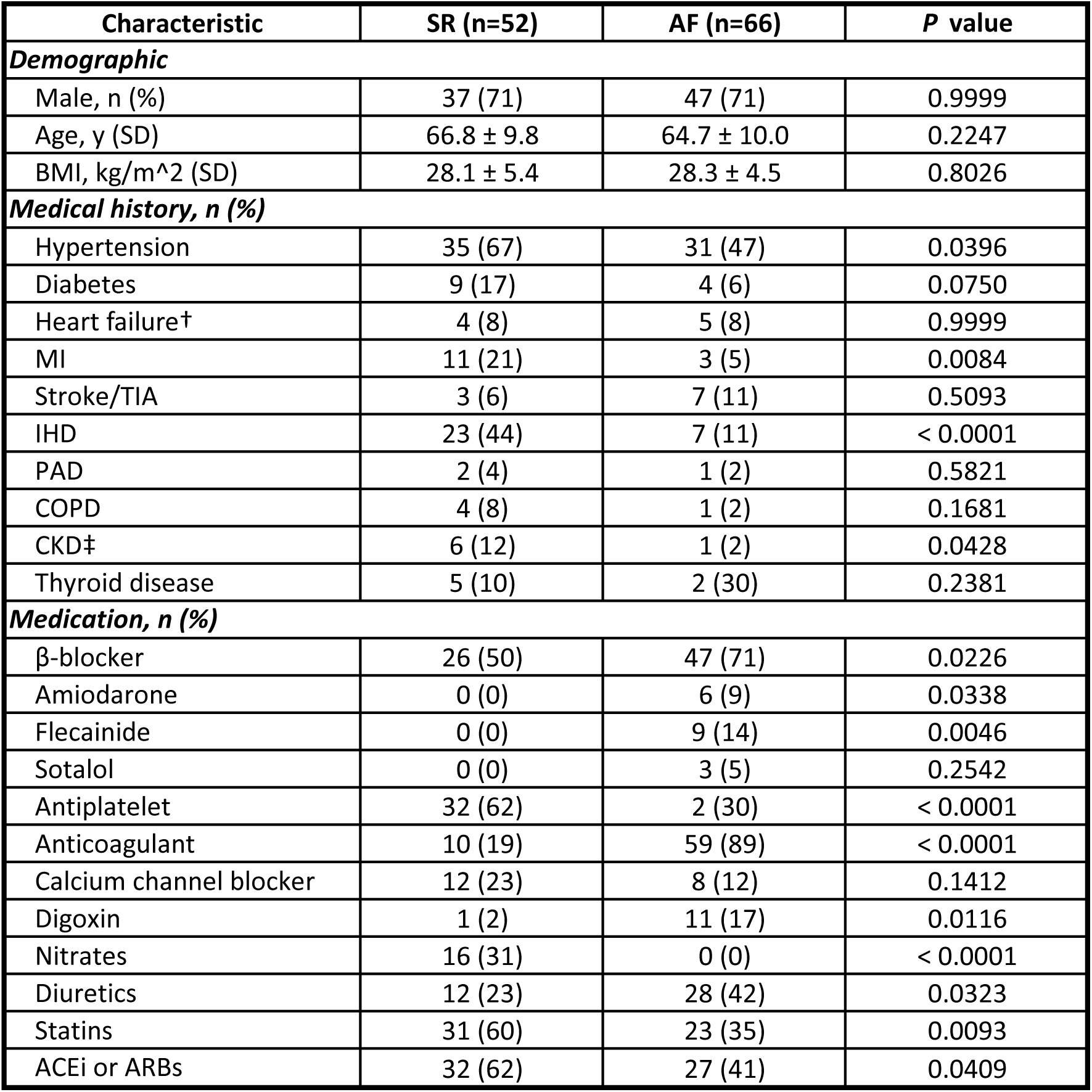
Baseline characteristics of patients with PV serum samples. ACEi, angiotensin-converting enzyme inhibitor; AF, atrial fibrillation; ARBs, angiotensin II receptor blockers; AVR, aortic valve replacement; BMI, body mass index; CABG, coronary artery bypass graft; CKD, chronic kidney disease; COPD, chronic obstructory pulmonary disease; IHD, ischemic heart disease; MI, myocardial infarction; MVR, mitral valve repair/replacement; PAD, peripheral arterial disease; SR, sinus rhythm; TIA, transient ischemic attack *P* values were calculated by the Mann-Whitney test for Age, unpaired t-test for BMI, and Fisher’s exact test for other characteristics. † Heart failure is determined by ejection fraction < 40%. ‡ CKD is determined by eGFR < 60 ml/min/1.73 m^2.

A comparison of characteristics between non-AF and AF patients is summarised in **Table 3**. The characteristics did not differ between the two groups with the exceptions that AF patients were older than non-AF patients (non-AF 55.3 ± 14.0, AF 62.9 ± 10.7; *P* = 0.0208) and more frequently prescribed anticoagulants (non-AF 40%, AF 93%, *P* < 0.0001).

**Table 3:** Baseline characteristics of patients with CS serum samples. ACEi, angiotensin-converting enzyme inhibitor; AF, atrial fibrillation; ARBs, angiotensin II receptor blockers; AVR, aortic valve replacement; BMI, body mass index; CABG, coronary artery bypass graft; CKD, chronic kidney disease; COPD, chronic obstructory pulmonary disease; IHD, ischemic heart disease; MI, myocardial infarction; MVR, mitral valve repair/replacement; PAD, peripheral arterial disease; TIA, transient ischemic attack *P* values were calculated by the unpaired t-test for Age, Mann-Whitney test for BMI, and Fisher’s exact test for other characteristics. † Heart failure is determined by ejection fraction < 40%. ‡ CKD is determined by eGFR < 60 ml/min/1.73 m^2.

| Characteristic | Non-AF (n=20) | AF (n=42) | *P* value |
| --- | --- | --- | --- |
| **Demographic** |  |  |  |
| Male, n (%) | 13 (65) | 28 (67) | 0.9999 |
| Age, y (SD) | 55.3 ± 14.0 | 62.9 ± 10.7 | 0.0208 |
| BMI, kg/m<sup>2</sup> (SD) | 29.7 ± 12.8 | 28.4 ± 4.7 | 0.4941 |
| **Medical history, n (%)** |  |  |  |
| Hypertension | 4 (20) | 14 (33) | 0.3748 |
| Diabetes | 0 (0) | 3 (7) | 0.5447 |
| Heart failure† | 1 (5) | 3 (7) | 0.9999 |
| MI | 1 (5) | 0 (0) | 0.3226 |
| Stroke/TIA | 1 (5) | 2 (5) | 0.9999 |
| IHD | 0 (0) | 3 (7) | 0.5447 |
| PAD | 0 (0) | 0 (0) | N/A |
| COPD | 0 (0) | 2 (5) | 0.9999 |
| CKD‡ | 0 (0) | 0 (0) | N/A |
| Thyroid disease | 0 (0) | 2 (5) | 0.9999 |
| **Medication, n (%)** |  |  |  |
| β-blocker | 11 (55) | 27 (64) | 0.5801 |
| Amiodarone | 2 (10) | 5 (12) | 0.9999 |
| Flecainide | 2 (10) | 8 (19) | 0.4776 |
| Sotalol | 0 (0) | 3 (7) | 0.5447 |
| Antiplatelet | 2 (10) | 0 (0) | 0.1005 |
| Anticoagulant | 8 (40) | 39 (93) | <0.0001 |
| Calcium channel blocker | 1 (5) | 6 (14) | 0.4119 |
| Digoxin | 3 (15) | 6 (14) | 0.9999 |
| Nitrates | 0 (0) | 0 (0) | N/A |
| Diuretics | 1 (5) | 9 (21) | 0.1463 |
| Statins | 6 (30) | 12 (29) | 0.9999 |
| ACEi or ARBs | 4 (20) | 13 (31) | 0.5439 |

### Associations of serum N-glycans and derived serum N-glycosylation traits with AF

UHPLC analysis identified sixty-two relatively quantified chromatographic peaks in the human serum, corresponding to 62 *N*-glycan traits (**Fig. 1a**). We evaluated *N*-glycan peaks in the PV serum samples between SR controls and AF patients (**Table 4**.) AF was associated with 27 individual glycan peaks which were differentially abundant (*P* < 0.05). Of these, 17 peaks were increased, while 10 peaks were decreased in PV serum (**Fig. 1b** and **Supplementary Fig. 2a**). In addition to peripheral changes, to understand the modifications of *N*-glycans locally in the heart in AF, we performed the analysis of *N*-glycans in the cardiac CS serum samples collected from non-AF controls and AF patients (**Table 5**). We found that 8 glycan peaks were significantly associated with AF (*P* < 0.05), including 2 upregulated and 6 downregulated peaks (**Fig. 1c** and **Supplementary Fig. 2**).

**Figure 1.**
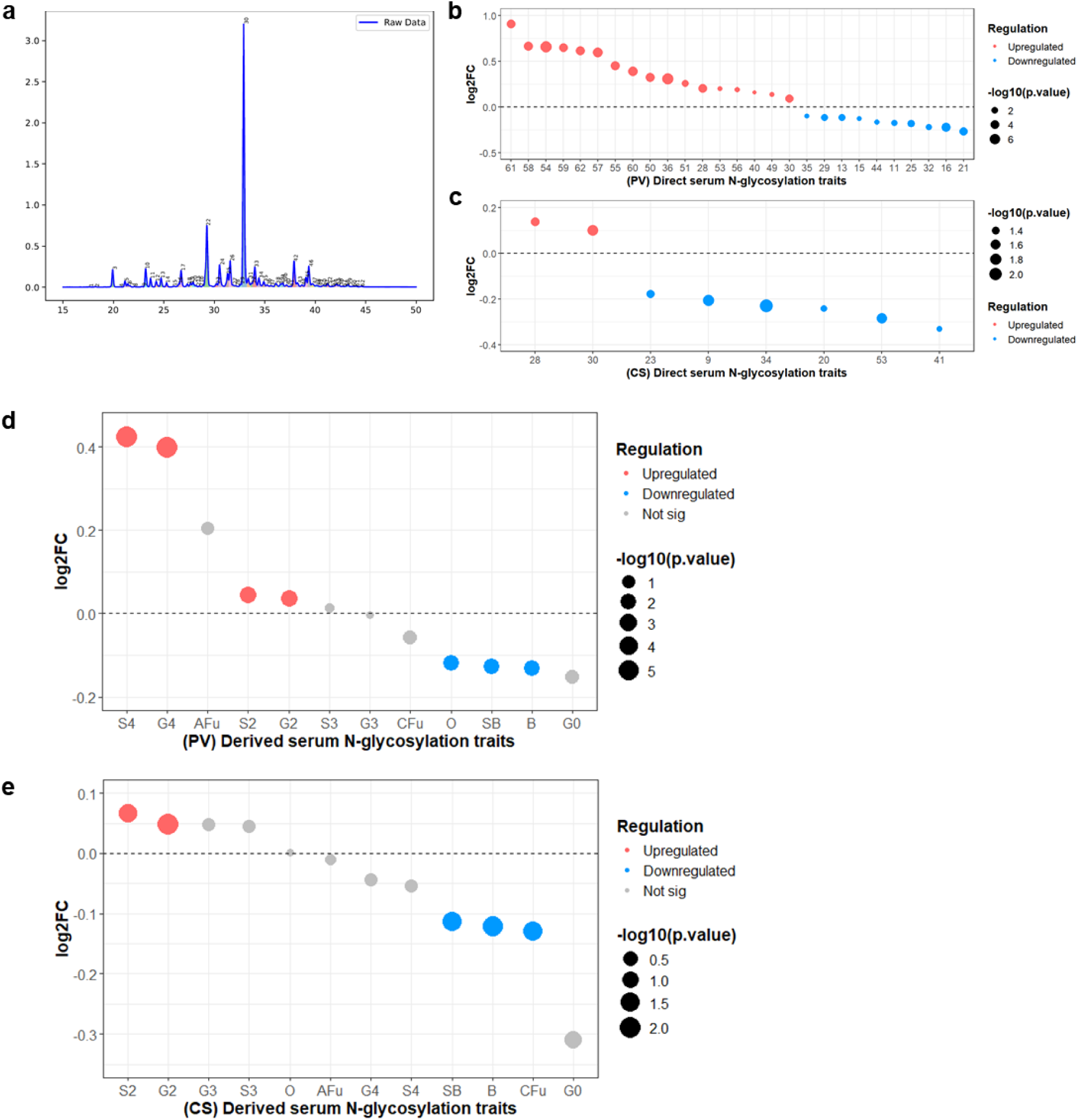
Associations of serum *N*-glycans and derived traits with AF. (**a**) Chromatogram of glycan peaks in human serum identified by UHPLC analysis. (**b**) Significantly regulated *N*-glycan peaks in PV serum between SR (N=52) and AF (N=66). (**c**) Significantly regulated *N*-glycan peaks in CS serum between non-AF (N=20) and AF (N=42) groups. (**d**) Significantly regulated derived traits in PV serum between SR and AF. (**e**) Significantly regulated derived traits in CS serum between non-AF and AF groups. Full name of derived traits is described in Table 1. Data are presented as dot plots and expressed as a fold change of the control group (SR or non-AF). *P* values were determined by unpaired t-test, Welch’s t-test or Mann-Whitney test. AF, atrial fibrillation; CS, coronary sinus; PV, peripheral venous; SR, sinus rhythm; UHPLC, ultra-high performance liquid chromatography.

**Table 4:** Glycan profiling of baseline PV serum samples. Glycan LC peaks were extracted from the fluorescence data analysis and quantified using HappyTools. AF, atrial fibrillation; log2FC, log2 fold change; LC, liquid chromatography; SR, sinus rhythm

| LC Peak | SR<br>(mean ± SD, n=52) | AF<br>(mean ± SD, n=66) | log2FC | P value | Test |
| --- | --- | --- | --- | --- | --- |
| 1 | 0.000874±0.0002 | 0.000863±0.0004 | -0.018273 | 0.2917 | Mann-Whitney test |
| 2 | 0.00169±0.0009 | 0.001648±0.0013 | -0.036307 | 0.2095 | Mann-Whitney test |
| 3 | 0.040151±0.0125 | 0.035804±0.0120 | -0.165315 | 0.0533 | Mann-Whitney test |
| 4 | 0.001133±0.0003 | 0.001042±0.0004 | -0.120793 | 0.1528 | Welch Two Sample t-test |
| 5 | 0.009576±0.0020 | 0.00913±0.0017 | -0.068808 | 0.2917 | Mann-Whitney test |
| 6 | 0.008873±0.0028 | 0.008117±0.0027 | -0.128475 | 0.1905 | Mann-Whitney test |
| 7 | 0.003315±0.0008 | 0.003185±0.0017 | -0.057715 | 0.0547 | Mann-Whitney test |
| 8 | 0.001269±0.0004 | 0.001265±0.0008 | -0.004555 | 0.1426 | Mann-Whitney test |
| 9 | 0.002274±0.0006 | 0.002121±0.0007 | -0.100488 | 0.0706 | Mann-Whitney test |
| 10 | 0.031081±0.0070 | 0.029163±0.0066 | -0.091894 | 0.1299 | Two Sample t-test |
| 11 | 0.01465±0.0038 | 0.012981±0.0032 | -0.174499 | 0.0162 | Mann-Whitney test |
| 12 | 0.01258±0.0024 | 0.011969±0.0029 | -0.071829 | 0.2274 | Two Sample t-test |
| 13 | 0.015273±0.0024 | 0.014086±0.0018 | -0.116721 | 0.0058 | Mann-Whitney test |
| 14 | 0.007307±0.0014 | 0.007466±0.0027 | 0.031056 | 0.3199 | Mann-Whitney test |
| 15 | 0.003076±0.0005 | 0.002817±0.0008 | -0.126896 | 0.0422 | Welch Two Sample t-test |
| 16 | 0.005895±0.0010 | 0.005055±0.0011 | -0.221781 | 4.73E-05 | Two Sample t-test |
| 17 | 0.021273±0.0060 | 0.021808±0.0067 | 0.035834 | 0.9805 | Mann-Whitney test |
| 18 | 0.005593±0.0013 | 0.005133±0.0013 | -0.123820 | 0.0537 | Two Sample t-test |
| 19 | 0.007737±0.0013 | 0.007409±0.0016 | -0.062495 | 0.0574 | Mann-Whitney test |
| 20 | 0.007772±0.0016 | 0.007602±0.0017 | -0.031907 | 0.5102 | Mann-Whitney test |
| 21 | 0.003503±0.0008 | 0.002902±0.0007 | -0.271543 | 0.0003 | Mann-Whitney test |
| 22 | 0.10919±0.0117 | 0.106717±0.0113 | -0.033051 | 0.2481 | Two Sample t-test |
| 23 | 0.008222±0.0023 | 0.007794±0.0021 | -0.077125 | 0.2917 | Mann-Whitney test |
| 24 | 0.037154±0.0079 | 0.035877±0.0072 | -0.050458 | 0.3592 | Two Sample t-test |
| 25 | 0.029467±0.0066 | 0.025923±0.0055 | -0.184868 | 0.0020 | Mann-Whitney test |
| 26 | 0.029447±0.0061 | 0.028336±0.0059 | -0.055485 | 0.3194 | Two Sample t-test |
| 27 | 0.002352±0.0006 | 0.002341±0.0006 | -0.006763 | 0.9805 | Mann-Whitney test |
| 28 | 0.001337±0.0003 | 0.00154±0.0003 | 0.203931 | 0.0001 | Mann-Whitney test |
| 29 | 0.003591±0.0005 | 0.003313±0.0005 | -0.116247 | 0.0044 | Two Sample t-test |
| 30 | 0.317546±0.0309 | 0.337865±0.0274 | 0.089481 | 0.0002 | Two Sample t-test |
| 31 | 0.012668±0.0016 | 0.012197±0.0016 | -0.054662 | 0.1516 | Mann-Whitney test |
| 32 | 0.005587±0.0016 | 0.004797±0.0017 | -0.219941 | 0.0175 | Mann-Whitney test |
| 33 | 0.036311±0.0099 | 0.035416±0.0073 | -0.036005 | 0.9805 | Mann-Whitney test |
| 34 | 0.024942±0.0087 | 0.0223±0.0066 | -0.161533 | 0.1532 | Mann-Whitney test |
| 35 | 0.01258±0.0028 | 0.011732±0.0033 | -0.100683 | 0.0424 | Mann-Whitney test |
| 36 | 0.001954±0.0004 | 0.002421±0.0005 | 0.309173 | 3.17E-08 | Mann-Whitney test |
| 37 | 0.001727±0.0003 | 0.001643±0.0004 | -0.071936 | 0.1999 | Mann-Whitney test |
| 38 | 0.010522±0.0025 | 0.01017±0.0022 | -0.049089 | 0.4211 | Two Sample t-test |
| 39 | 0.010679±0.0020 | 0.010645±0.0021 | -0.004601 | 0.9292 | Two Sample t-test |
| 40 | 0.002018±0.0005 | 0.002253±0.0007 | 0.158921 | 0.0481 | Welch Two Sample t-test |
| 41 | 0.000541±0.0002 | 0.001222±0.0016 | 1.175544 | 0.2403 | Mann-Whitney test |
| 42 | 0.048208±0.0140 | 0.045553±0.0147 | -0.081727 | 0.1999 | Mann-Whitney test |
| 43 | 0.006736±0.0015 | 0.007228±0.0014 | 0.101704 | 0.0745 | Two Sample t-test |
| 44 | 0.003995±0.0011 | 0.003564±0.0012 | -0.164698 | 0.0320 | Mann-Whitney test |
| 45 | 0.016555±0.0059 | 0.017112±0.0042 | 0.047741 | 0.5636 | Welch Two Sample t-test |
| 46 | 0.028963±0.0120 | 0.029473±0.0124 | 0.025183 | 0.8222 | Two Sample t-test |
| 47 | 0.008044±0.0016 | 0.008519±0.0014 | 0.082771 | 0.0882 | Two Sample t-test |
| 48 | 0.00267±0.0013 | 0.002565±0.0014 | -0.057881 | 0.4760 | Mann-Whitney test |
| 49 | 0.000843±0.0002 | 0.000925±0.0002 | 0.133921 | 0.0390 | Two Sample t-test |
| 50 | 0.001894±0.0007 | 0.002364±0.0006 | 0.319794 | 0.0001 | Two Sample t-test |
| 51 | 0.001863±0.0007 | 0.002228±0.0006 | 0.258122 | 0.0030 | Two Sample t-test |
| 52 | 0.004439±0.0018 | 0.004913±0.0017 | 0.146370 | 0.1270 | Mann-Whitney test |
| 53 | 0.000428±0.0001 | 0.000492±0.0002 | 0.201048 | 0.0352 | Welch Two Sample t-test |
| 54 | 0.000963±0.0005 | 0.001519±0.0005 | 0.657514 | 4.16E-08 | Two Sample t-test |
| 55 | 0.003135±0.0015 | 0.004283±0.0012 | 0.450156 | 2.76E-05 | Mann-Whitney test |
| 56 | 0.002526±0.0009 | 0.002882±0.0008 | 0.190216 | 0.0270 | Two Sample t-test |
| 57 | 0.000504±0.0003 | 0.000762±0.0003 | 0.596367 | 5.35E-06 | Mann-Whitney test |
| 58 | 0.000666±0.0005 | 0.001053±0.0006 | 0.660911 | 0.0001 | Mann-Whitney test |
| 59 | 0.002107±0.0016 | 0.003302±0.0014 | 0.648150 | 0.0002 | Mann-Whitney test |
| 60 | 0.001229±0.0004 | 0.001613±0.0005 | 0.392262 | 9.16E-06 | Two Sample t-test |
| 61 | 0.00084±0.0009 | 0.001577±0.0012 | 0.908721 | 0.0001 | Mann-Whitney test |
| 62 | 0.001359±0.0009 | 0.002078±0.0009 | 0.612650 | 2.75E-05 | Mann-Whitney test |

**Table 5:**
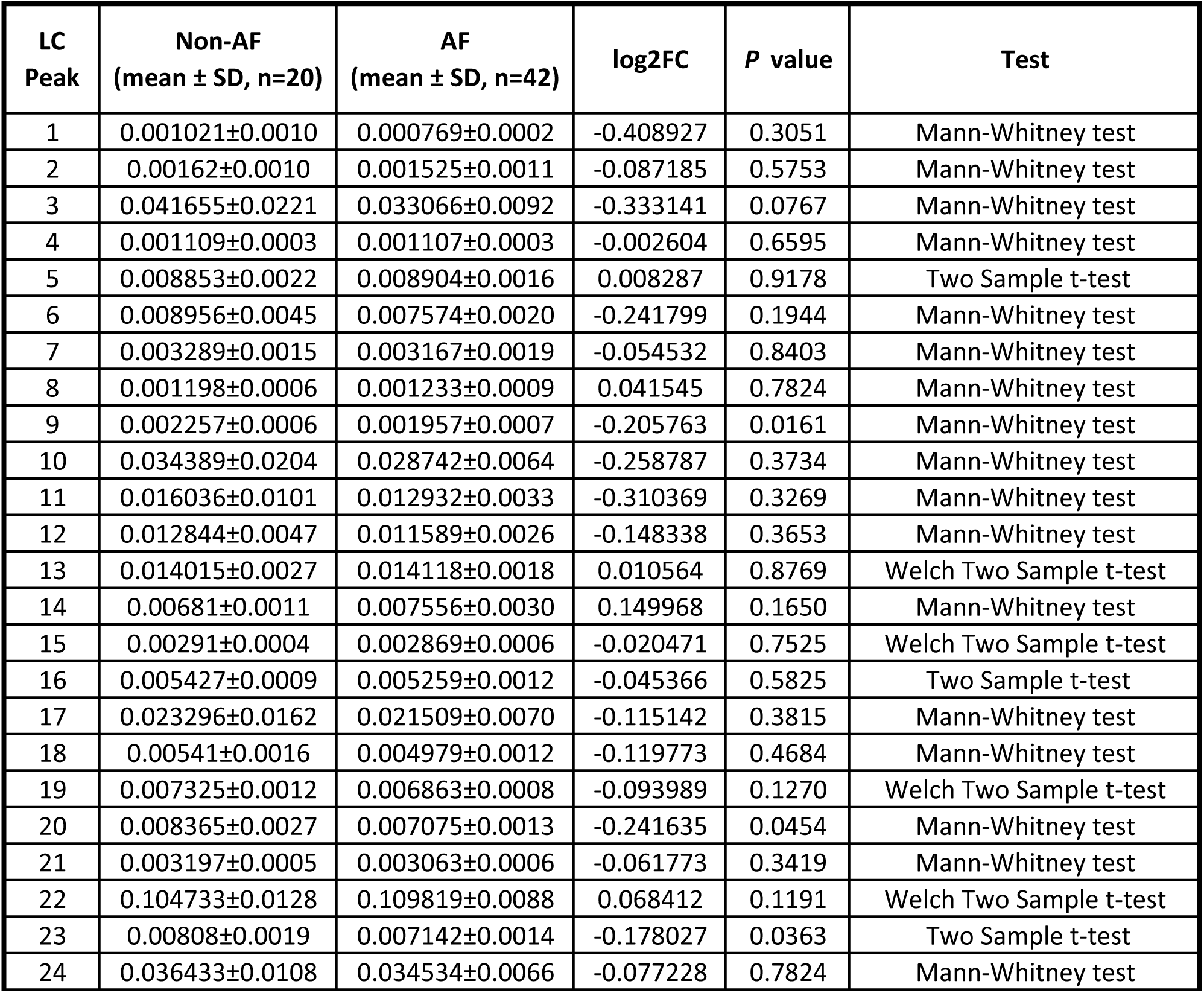
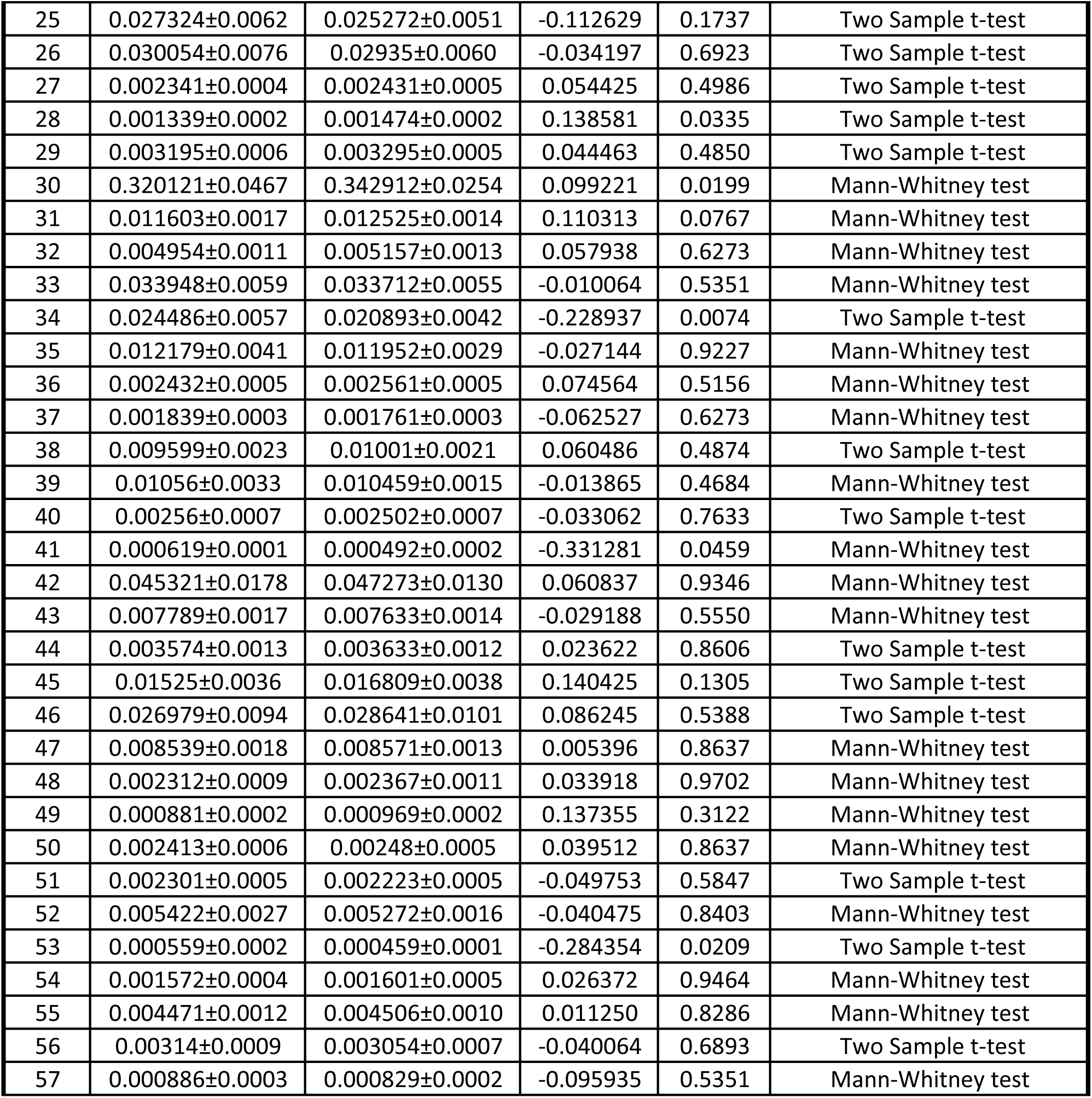
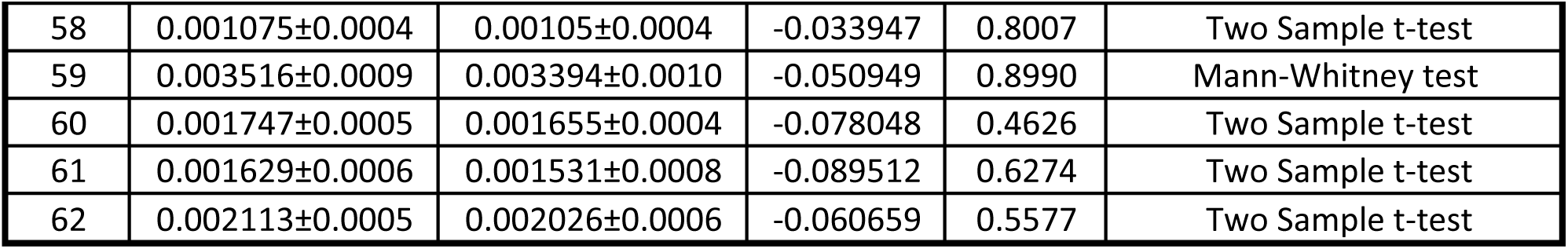
Glycan profiling of baseline CS serum samples. Glycan LC peaks were extracted from the fluorescence data analysis and quantified using HappyTools. AF, atrial fibrillation; log2FC, log2 fold change;

| LC Peak | Non-AF<br>(mean ± SD, n=20) | AF<br>(mean ± SD, n=42) | log2FC | P value | Test |
| --- | --- | --- | --- | --- | --- |
| 1 | 0.001021±0.0010 | 0.000769±0.0002 | -0.408927 | 0.3051 | Mann-Whitney test |
| 2 | 0.00162±0.0010 | 0.001525±0.0011 | -0.087185 | 0.5753 | Mann-Whitney test |
| 3 | 0.041655±0.0221 | 0.033066±0.0092 | -0.333141 | 0.0767 | Mann-Whitney test |
| 4 | 0.001109±0.0003 | 0.001107±0.0003 | -0.002604 | 0.6595 | Mann-Whitney test |
| 5 | 0.008853±0.0022 | 0.008904±0.0016 | 0.008287 | 0.9178 | Two Sample t-test |
| 6 | 0.008956±0.0045 | 0.007574±0.0020 | -0.241799 | 0.1944 | Mann-Whitney test |
| 7 | 0.003289±0.0015 | 0.003167±0.0019 | -0.054532 | 0.8403 | Mann-Whitney test |
| 8 | 0.001198±0.0006 | 0.001233±0.0009 | 0.041545 | 0.7824 | Mann-Whitney test |
| 9 | 0.002257±0.0006 | 0.001957±0.0007 | -0.205763 | 0.0161 | Mann-Whitney test |
| 10 | 0.034389±0.0204 | 0.028742±0.0064 | -0.258787 | 0.3734 | Mann-Whitney test |
| 11 | 0.016036±0.0101 | 0.012932±0.0033 | -0.310369 | 0.3269 | Mann-Whitney test |
| 12 | 0.012844±0.0047 | 0.011589±0.0026 | -0.148338 | 0.3653 | Mann-Whitney test |
| 13 | 0.014015±0.0027 | 0.014118±0.0018 | 0.010564 | 0.8769 | Welch Two Sample t-test |
| 14 | 0.00681±0.0011 | 0.007556±0.0030 | 0.149968 | 0.1650 | Mann-Whitney test |
| 15 | 0.00291±0.0004 | 0.002869±0.0006 | -0.020471 | 0.7525 | Welch Two Sample t-test |
| 16 | 0.005427±0.0009 | 0.005259±0.0012 | -0.045366 | 0.5825 | Two Sample t-test |
| 17 | 0.023296±0.0162 | 0.021509±0.0070 | -0.115142 | 0.3815 | Mann-Whitney test |
| 18 | 0.00541±0.0016 | 0.004979±0.0012 | -0.119773 | 0.4684 | Mann-Whitney test |
| 19 | 0.007325±0.0012 | 0.006863±0.0008 | -0.093989 | 0.1270 | Welch Two Sample t-test |
| 20 | 0.008365±0.0027 | 0.007075±0.0013 | -0.241635 | 0.0454 | Mann-Whitney test |
| 21 | 0.003197±0.0005 | 0.003063±0.0006 | -0.061773 | 0.3419 | Mann-Whitney test |
| 22 | 0.104733±0.0128 | 0.109819±0.0088 | 0.068412 | 0.1191 | Welch Two Sample t-test |
| 23 | 0.00808±0.0019 | 0.007142±0.0014 | -0.178027 | 0.0363 | Two Sample t-test |
| 24 | 0.036433±0.0108 | 0.034534±0.0066 | -0.077228 | 0.7824 | Mann-Whitney test |
| 25 | 0.027324±0.0062 | 0.025272±0.0051 | -0.112629 | 0.1737 | Two Sample t-test |
| 26 | 0.030054±0.0076 | 0.02935±0.0060 | -0.034197 | 0.6923 | Two Sample t-test |
| 27 | 0.002341±0.0004 | 0.002431±0.0005 | 0.054425 | 0.4986 | Two Sample t-test |
| 28 | 0.001339±0.0002 | 0.001474±0.0002 | 0.138581 | 0.0335 | Two Sample t-test |
| 29 | 0.003195±0.0006 | 0.003295±0.0005 | 0.044463 | 0.4850 | Two Sample t-test |
| 30 | 0.320121±0.0467 | 0.342912±0.0254 | 0.099221 | 0.0199 | Mann-Whitney test |
| 31 | 0.011603±0.0017 | 0.012525±0.0014 | 0.110313 | 0.0767 | Mann-Whitney test |
| 32 | 0.004954±0.0011 | 0.005157±0.0013 | 0.057938 | 0.6273 | Mann-Whitney test |
| 33 | 0.033948±0.0059 | 0.033712±0.0055 | -0.010064 | 0.5351 | Mann-Whitney test |
| 34 | 0.024486±0.0057 | 0.020893±0.0042 | -0.228937 | 0.0074 | Two Sample t-test |
| 35 | 0.012179±0.0041 | 0.011952±0.0029 | -0.027144 | 0.9227 | Mann-Whitney test |
| 36 | 0.002432±0.0005 | 0.002561±0.0005 | 0.074564 | 0.5156 | Mann-Whitney test |
| 37 | 0.001839±0.0003 | 0.001761±0.0003 | -0.062527 | 0.6273 | Mann-Whitney test |
| 38 | 0.009599±0.0023 | 0.01001±0.0021 | 0.060486 | 0.4874 | Two Sample t-test |
| 39 | 0.01056±0.0033 | 0.010459±0.0015 | -0.013865 | 0.4684 | Mann-Whitney test |
| 40 | 0.00256±0.0007 | 0.002502±0.0007 | -0.033062 | 0.7633 | Two Sample t-test |
| 41 | 0.000619±0.0001 | 0.000492±0.0002 | -0.331281 | 0.0459 | Mann-Whitney test |
| 42 | 0.045321±0.0178 | 0.047273±0.0130 | 0.060837 | 0.9346 | Mann-Whitney test |
| 43 | 0.007789±0.0017 | 0.007633±0.0014 | -0.029188 | 0.5550 | Mann-Whitney test |
| 44 | 0.003574±0.0013 | 0.003633±0.0012 | 0.023622 | 0.8606 | Two Sample t-test |
| 45 | 0.01525±0.0036 | 0.016809±0.0038 | 0.140425 | 0.1305 | Two Sample t-test |
| 46 | 0.026979±0.0094 | 0.028641±0.0101 | 0.086245 | 0.5388 | Two Sample t-test |
| 47 | 0.008539±0.0018 | 0.008571±0.0013 | 0.005396 | 0.8637 | Mann-Whitney test |
| 48 | 0.002312±0.0009 | 0.002367±0.0011 | 0.033918 | 0.9702 | Mann-Whitney test |
| 49 | 0.000881±0.0002 | 0.000969±0.0002 | 0.137355 | 0.3122 | Mann-Whitney test |
| 50 | 0.002413±0.0006 | 0.00248±0.0005 | 0.039512 | 0.8637 | Mann-Whitney test |
| 51 | 0.002301±0.0005 | 0.002223±0.0005 | -0.049753 | 0.5847 | Two Sample t-test |
| 52 | 0.005422±0.0027 | 0.005272±0.0016 | -0.040475 | 0.8403 | Mann-Whitney test |
| 53 | 0.000559±0.0002 | 0.000459±0.0001 | -0.284354 | 0.0209 | Two Sample t-test |
| 54 | 0.001572±0.0004 | 0.001601±0.0005 | 0.026372 | 0.9464 | Mann-Whitney test |
| 55 | 0.004471±0.0012 | 0.004506±0.0010 | 0.011250 | 0.8286 | Mann-Whitney test |
| 56 | 0.00314±0.0009 | 0.003054±0.0007 | -0.040064 | 0.6893 | Two Sample t-test |
| 57 | 0.000886±0.0003 | 0.000829±0.0002 | -0.095935 | 0.5351 | Mann-Whitney test |
| 58 | 0.001075±0.0004 | 0.00105±0.0004 | -0.033947 | 0.8007 | Two Sample t-test |
| 59 | 0.003516±0.0009 | 0.003394±0.0010 | -0.050949 | 0.8990 | Mann-Whitney test |
| 60 | 0.001747±0.0005 | 0.001655±0.0004 | -0.078048 | 0.4626 | Two Sample t-test |
| 61 | 0.001629±0.0006 | 0.001531±0.0008 | -0.089512 | 0.6274 | Two Sample t-test |
| 62 | 0.002113±0.0005 | 0.002026±0.0006 | -0.060659 | 0.5577 | Two Sample t-test |

To assess the overall *N*-glycosylation patterns, we grouped individual peaks with similar properties and identified 12 derived traits (**Table 1**). 7 traits were significantly altered in AF in PV serum (**Fig. 1d** and **Table 6**). In particular, S4, G4, S2 and G2 traits were upregulated, while O, SB and B traits were downregulated in AF. These glycosylation patterns were consistent with the observed differences in the individual *N*-glycan peaks. In the CS serum samples, 5 derived traits were altered in AF (**Fig. 1e** and **Table 7**), including higher levels of S2 and G2, and lower levels of SB, B and CFu traits. All significantly altered traits in the CS serum, with the exception of CFu, were overlapped in the PV pools.

**Table 6:**
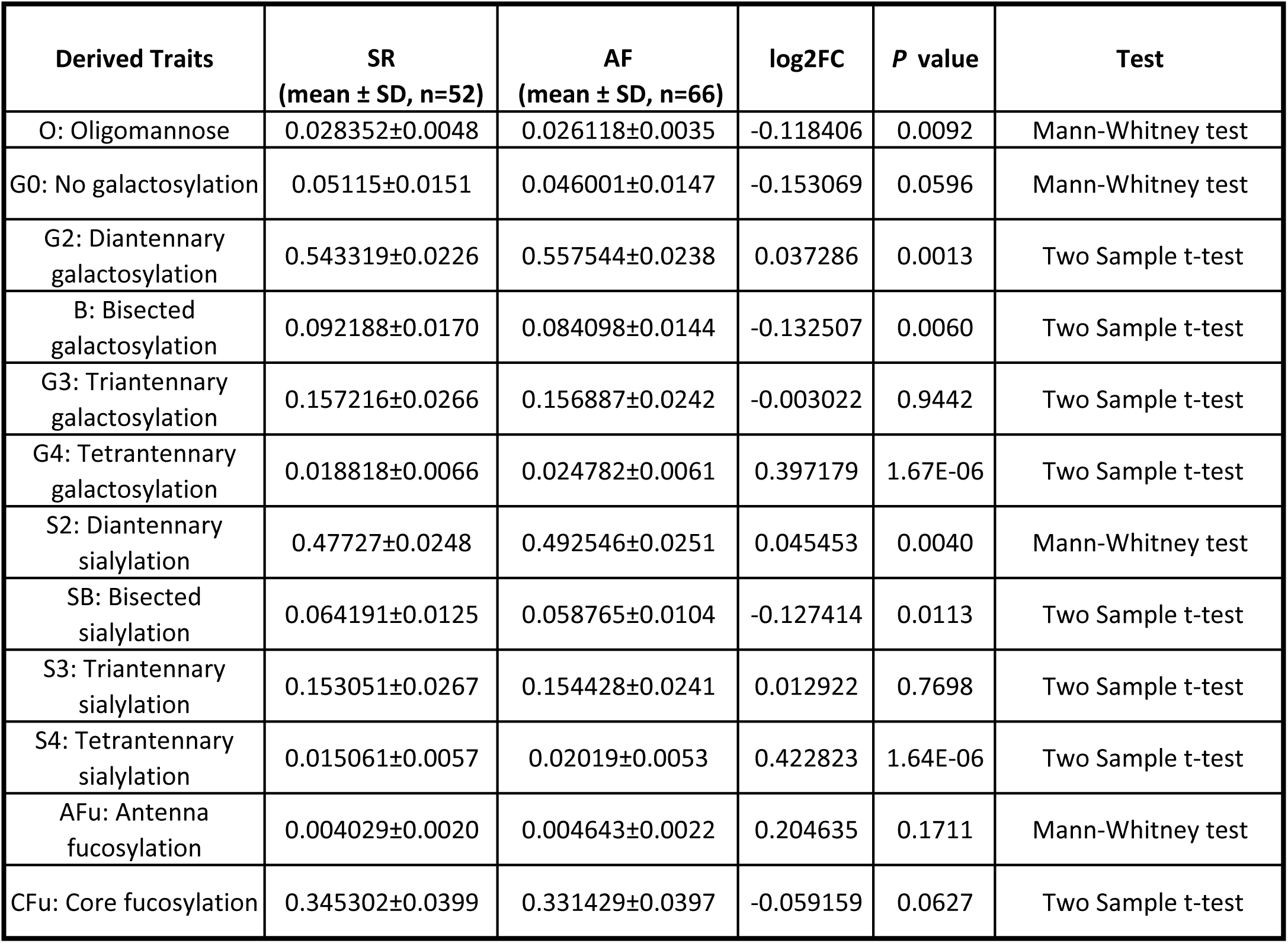
Associations of derived *N* -glycosylation traits with AF in baseline PV serum samples. AF, atrial fibrillation; log2FC, log2 fold change; SR, sinus rhythm

| Derived Traits | SR<br>(mean ± SD, n=52) | AF<br>(mean ± SD, n=66) | log2FC | *P* value | Test |
| --- | --- | --- | --- | --- | --- |
| O: Oligomannose | 0.028352±0.0048 | 0.026118±0.0035 | -0.118406 | 0.0092 | Mann-Whitney test |
| G0: No galactosylation | 0.05115±0.0151 | 0.046001±0.0147 | -0.153069 | 0.0596 | Mann-Whitney test |
| G2: Diantennary galactosylation | 0.543319±0.0226 | 0.557544±0.0238 | 0.037286 | 0.0013 | Two Sample t-test |
| B: Bisected galactosylation | 0.092188±0.0170 | 0.084098±0.0144 | -0.132507 | 0.0060 | Two Sample t-test |
| G3: Triantennary galactosylation | 0.157216±0.0266 | 0.156887±0.0242 | -0.003022 | 0.9442 | Two Sample t-test |
| G4: Tetrantennary galactosylation | 0.018818±0.0066 | 0.024782±0.0061 | 0.397179 | 1.67E-06 | Two Sample t-test |
| S2: Diantennary sialylation | 0.47727±0.0248 | 0.492546±0.0251 | 0.045453 | 0.0040 | Mann-Whitney test |
| SB: Bisected sialylation | 0.064191±0.0125 | 0.058765±0.0104 | -0.127414 | 0.0113 | Two Sample t-test |
| S3: Triantennary sialylation | 0.153051±0.0267 | 0.154428±0.0241 | 0.012922 | 0.7698 | Two Sample t-test |
| S4: Tetrantennary sialylation | 0.015061±0.0057 | 0.02019±0.0053 | 0.422823 | 1.64E-06 | Two Sample t-test |
| AFu: Antenna fucosylation | 0.004029±0.0020 | 0.004643±0.0022 | 0.204635 | 0.1711 | Mann-Whitney test |
| CFu: Core fucosylation | 0.345302±0.0399 | 0.331429±0.0397 | -0.059159 | 0.0627 | Two Sample t-test |

**Table 7:** Associations of derived *N* -glycosylation traits with AF in baseline CS serum samples. AF, atrial fibrillation; log2FC, log2 fold change; SR, sinus rhythm

| Derived Traits | Non-AF<br>(mean ± SD, n=20) | AF<br>(mean ± SD, n=42) | log2FC | *P* value | Test |
| --- | --- | --- | --- | --- | --- |
| O: Oligomannose | 0.026065±0.0051 | 0.026085±0.0034 | 0.001107 | 0.9877 | Welch Two Sample t-test |
| G0: No galactosylation | 0.05274±0.0275 | 0.04255±0.0110 | -0.309738 | 0.0767 | Mann-Whitney test |
| G2: Diantennary galactosylation | 0.54374±0.0287 | 0.562097±0.0234 | 0.047902 | 0.0095 | Two Sample t-test |
| B: Bisected galactosylation | 0.088116±0.0110 | 0.081017±0.0105 | -0.121179 | 0.0176 | Two Sample t-test |
| G3: Triantennary galactosylation | 0.15247±0.0338 | 0.157579±0.0216 | 0.047550 | 0.4964 | Mann-Whitney test |
| G4: Tetrantennary galactosylation | 0.026372±0.0059 | 0.025574±0.0044 | -0.044329 | 0.5511 | Two Sample t-test |
| S2: Diantennary sialylation | 0.475298±0.0509 | 0.497658±0.0242 | 0.066322 | 0.0471 | Mann-Whitney test |
| SB: Bisected sialylation | 0.061116±0.0082 | 0.05649±0.0066 | -0.113555 | 0.0204 | Two Sample t-test |
| S3: Triantennary sialylation | 0.150625±0.0338 | 0.155462±0.0213 | 0.045601 | 0.5060 | Mann-Whitney test |
| S4: Tetrantennary sialylation | 0.021658±0.0051 | 0.020871±0.0038 | -0.053400 | 0.4946 | Two Sample t-test |
| AFu: Antenna fucosylation | 0.004425±0.0013 | 0.004393±0.0015 | -0.010471 | 0.7482 | Mann-Whitney test |
| CFu: Core fucosylation | 0.351014±0.0752 | 0.320785±0.0303 | -0.129922 | 0.0349 | Mann-Whitney test |

### Multivariate N-glycan scores for AF

In the PV dataset, LASSO logistic regression identified 11 glycans with non-zero coefficients at the optimal penalty adjusting for clinical covariates (age, sex, BMI, hypertension and IHD): peak 14, peak 17, peak 21, peak 28, peak 31, peak 32, peak 36, peak 41, peak 54, peak 60 and peak 62 (**Fig. 2a**). The model achieved an AUROC of 0.934 in discriminating AF from non-AF individuals (**Fig. 2b**). A glycan score derived solely from the selected glycans also demonstrated strong discrimination (AUROC = 0.9) as compared to the clinical model (AUROC = 0.716). Addition of the glycan score to the clinical model yielded significant improvements in predictive performance, with a continuous NRI of 1.337 (95% CI: 1.069–1.59; *P* < 0.0001), IDI of 0.316 (95% CI: 0.251-0.387, *P* < 0.0001), and Brier score of 0.1105. For multiclass classification of AF subtypes in PV dataset, multinomial LASSO regression retained 9 glycans with non-zero coefficients, adjusting for the clinical covariates: peak 7, peak 19, peak 28, peak 36, peak 40, peak 41, peak 54, peak 60 and peak 62 (**Supplementary Fig. 3a**). The multinomial model achieved excellent discrimination among AF subtypes, with macro-averaged AUROC of 0.852 (**Supplementary Fig. 3b**). The model also showed good macro-averaged NRI of 1.177, alongside macro-averaged IDI of 0.237 and multiclass Brier score of 0.3635.

**Figure 2.**
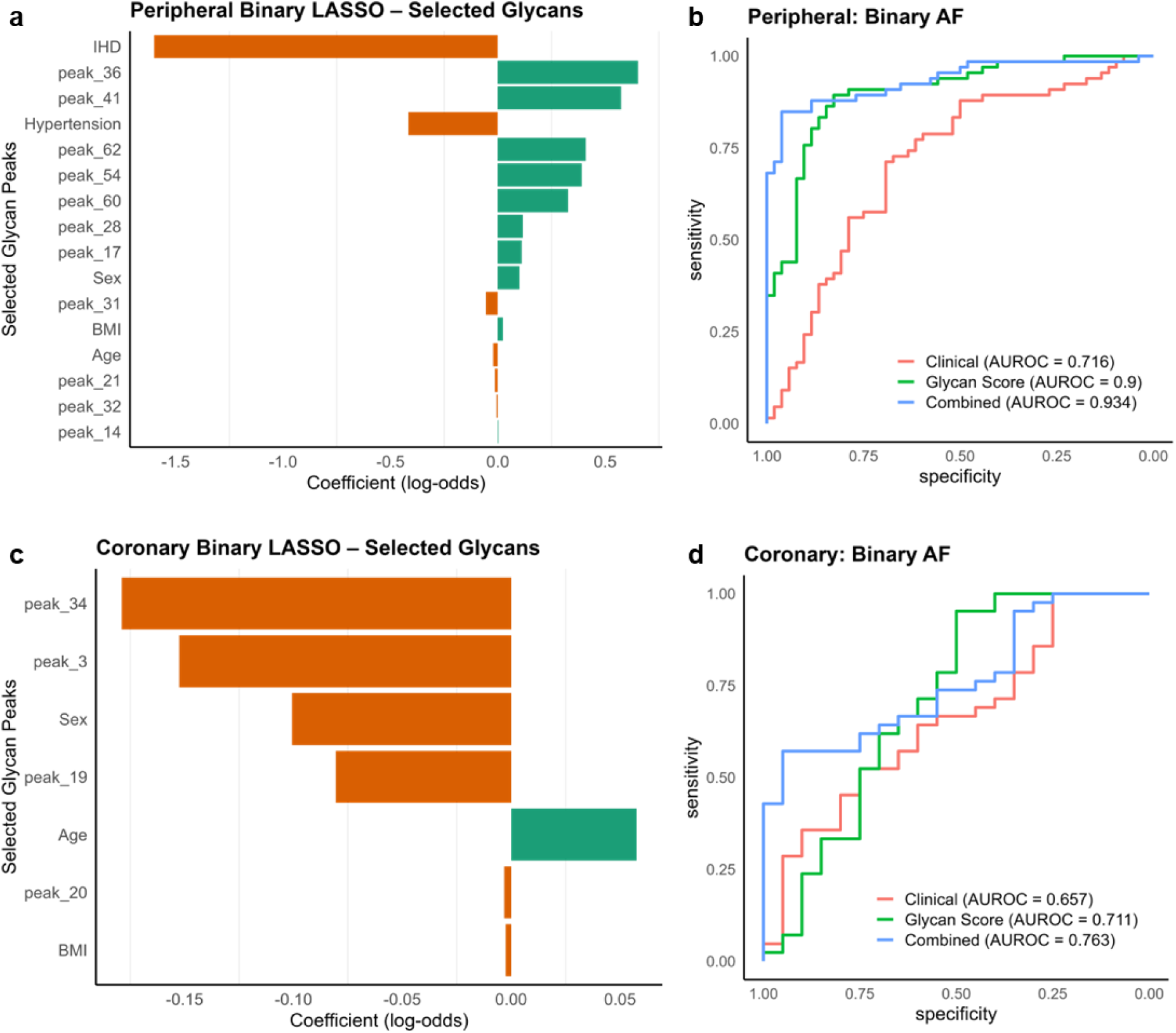
Multivariate *N*-glycan scores for binary classification of AF. (**a**) LASSO model was fitted to normalised PV glycan peaks with age, sex, BMI, hypertension and IHD specified as unpenalised covariates (N=118). (**b**) AUROC curves in PV dataset comparing three LASSO models for binary AF (no AF vs. any AF): clinical covariates only, glycan-only, and the combined model. (**c**) LASSO model was fitted to normalised CS glycan peaks with age, sex and BMI specified as unpenalised covariates (N=62). (**d**) AUROC curves in CS dataset comparing three LASSO models for binary AF (no AF vs. any AF): clinical covariates only, glycan-only, and the combined model. Bar plots display non-zero standardised coefficients (log-odds) retained at the optimal penalty, ordered by absolute magnitude (a, c). N represents individual biological donors (c, d). AF, atrial fibrillation; AUROC, area under the receiver operating characteristic curve; BMI, body mass index; CS, coronary sinus; IHD, ischemic heart disease; LASSO, least absolute shrinkage and selection operator; PV, peripheral venous.

In the CS dataset, LASSO logistic regression identified 4 glycans with non-zero coefficients: peak 3, peak 19, peak 20 and peak 34, adjusting for age, sex and BMI (**Fig. 2c**). The combined model achieved an AUROC of 0.763, while the glycan score achieved an AUROC of 0.711 and clinical model of 0.657 (**Fig. 2d**). The continuous NRI indicated reclassification improvement (NRI = 0.538, 95% CI: 0.049-1.04; *P* = 0.051) and the IDI confirmed improved discrimination (IDI = 0.05, 95% CI: 0.013-0.092; *P* = 0.011), along with the improved Brier score of 0.1782. For the multiclass CS model, the model achieved strong discrimination among AF subtypes, with macro-averaged AUROC = 0.741 (**Supplementary Fig. 3c**). Multinomial LASSO regression adjusted for clinical covariates retained 8 glycans with non-zero coefficients: peak 3, peak 9, peak 14, peak 19, peak 20, peak 28, peak 34 and peak 41(**Supplementary Fig. 3d**). The model also showed macro-averaged NRI of 0.613, IDI of 0.1 and Brier score of 0.5165. The combined models consistently outperformed the clinical and glycan-only models across all metrics supporting the additive predictive value of the glycan score.

### Correlation of PV glycan score for binary AF classification with PV plasma protein expression

To gain insights into circulating proteins that are likely to be *N*-glycosylated in AF, we evaluated whether any PV plasma proteins are correlated with the PV *N*-glycan score derived from the binary AF classification. Olink plasma proteomics data were available for a subgroup of patients from the serum *N*-glycome cohort (N=37). Prior to correlation analysis, 179 proteins were selected from the 1034 proteins included in the Olink Reveal panel based on two UniProt criteria: 1) presence of “*N*-glycosylation” under post-translational modifications and 2) designation as “Secreted” under subcellular location. Among the 179 filtered proteins, 11 showed significant correlation with PV glycan score (**Figure 3**). 7 proteins were positively correlated with PV glycan score, in which ISM1 exhibiting the strongest positive correlation (*r* = 0.4374, *P* = 0.0073). In contrast, FGF5 was demonstrated as the strongest negative correlation protein with PV glycan score (*r* = -0.4282, *P* = 0.0087). The correlation results for the complete set of filtered proteins are provided in **Table 8**.

**Figure 3.**
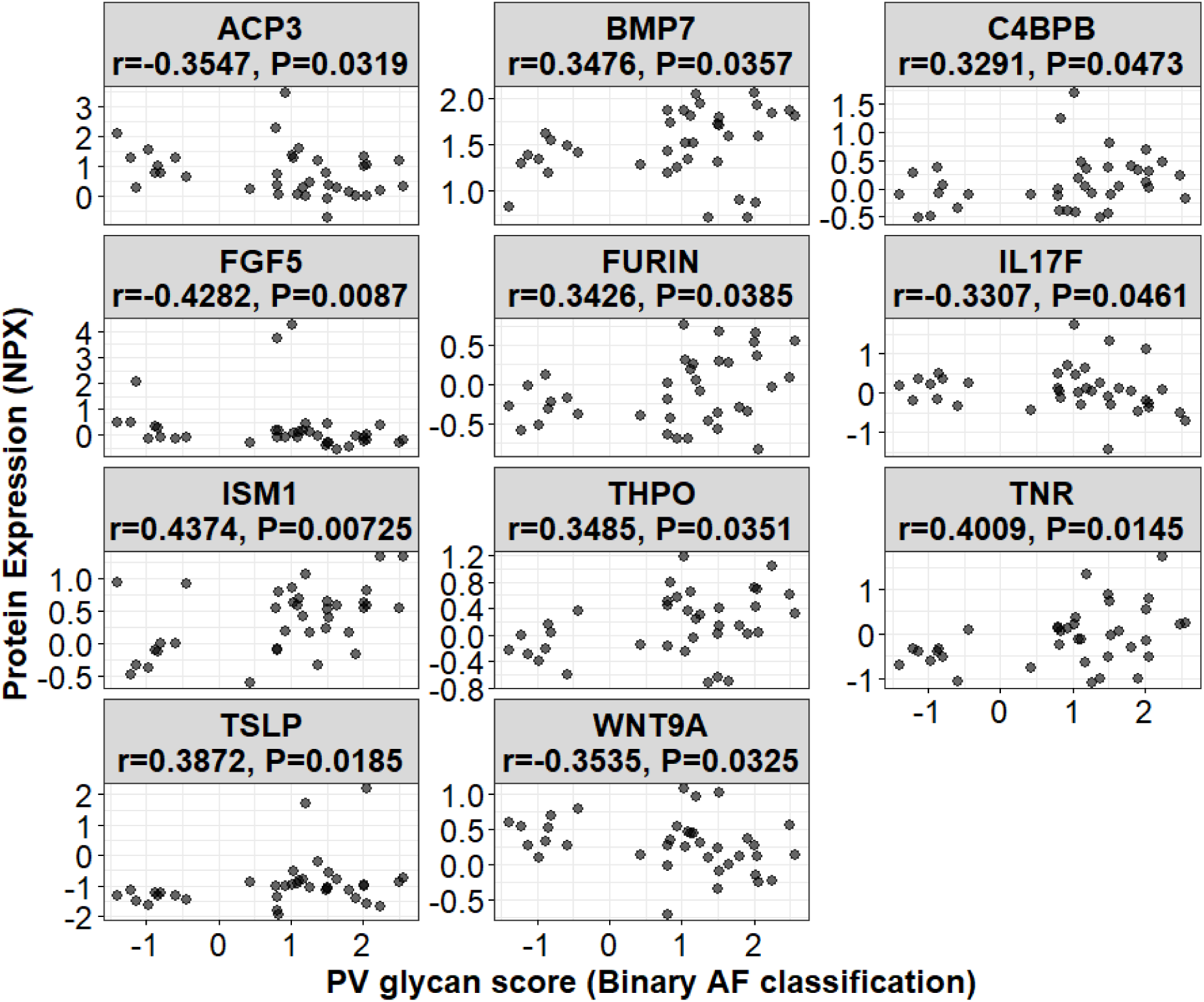
Correlation between PV glycan score and plasma protein levels at baseline. Dot plots showing the correlation between glycan score for binary AF classification in PV serum dataset and baseline levels of plasma proteins (N=37). Plasma protein levels are expressed as NPX values from the Olink assay. *P* values are determined by Spearman’s correlation test. Only significantly correlated proteins are plotted (*P* < 0.05). AF, atrial fibrillation; NPX, normalised protein expression; PV, peripheral venous.

**Table 8:**
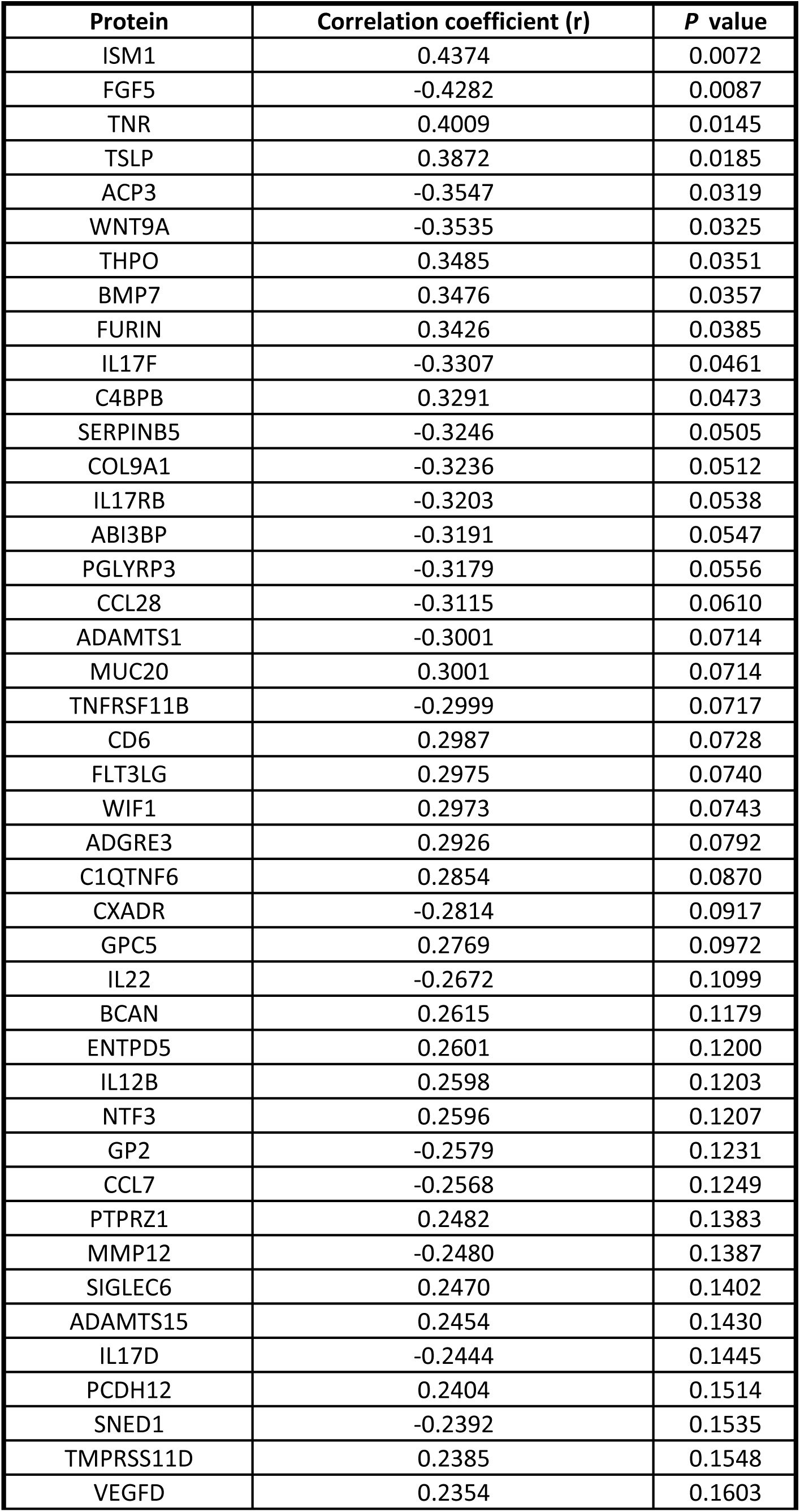
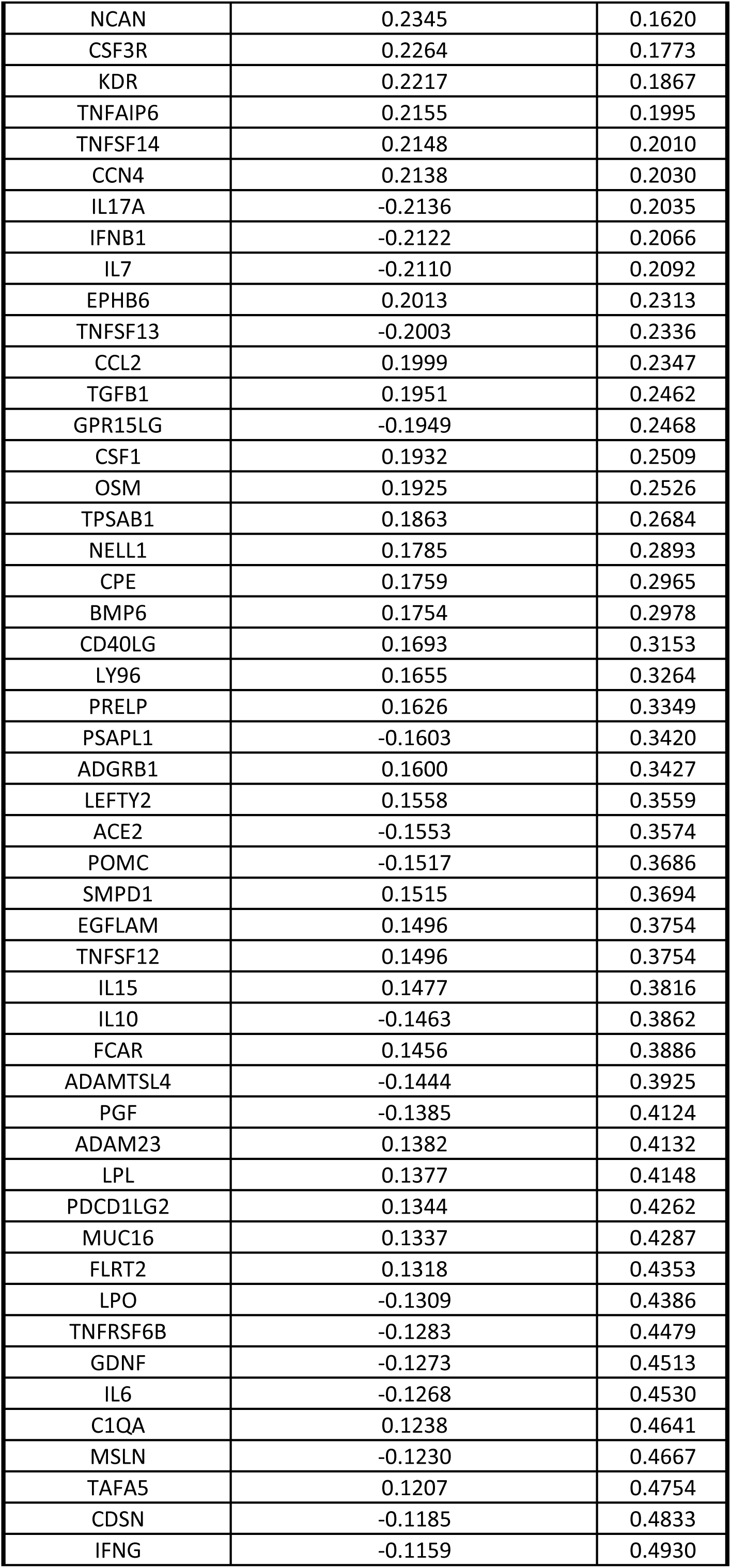
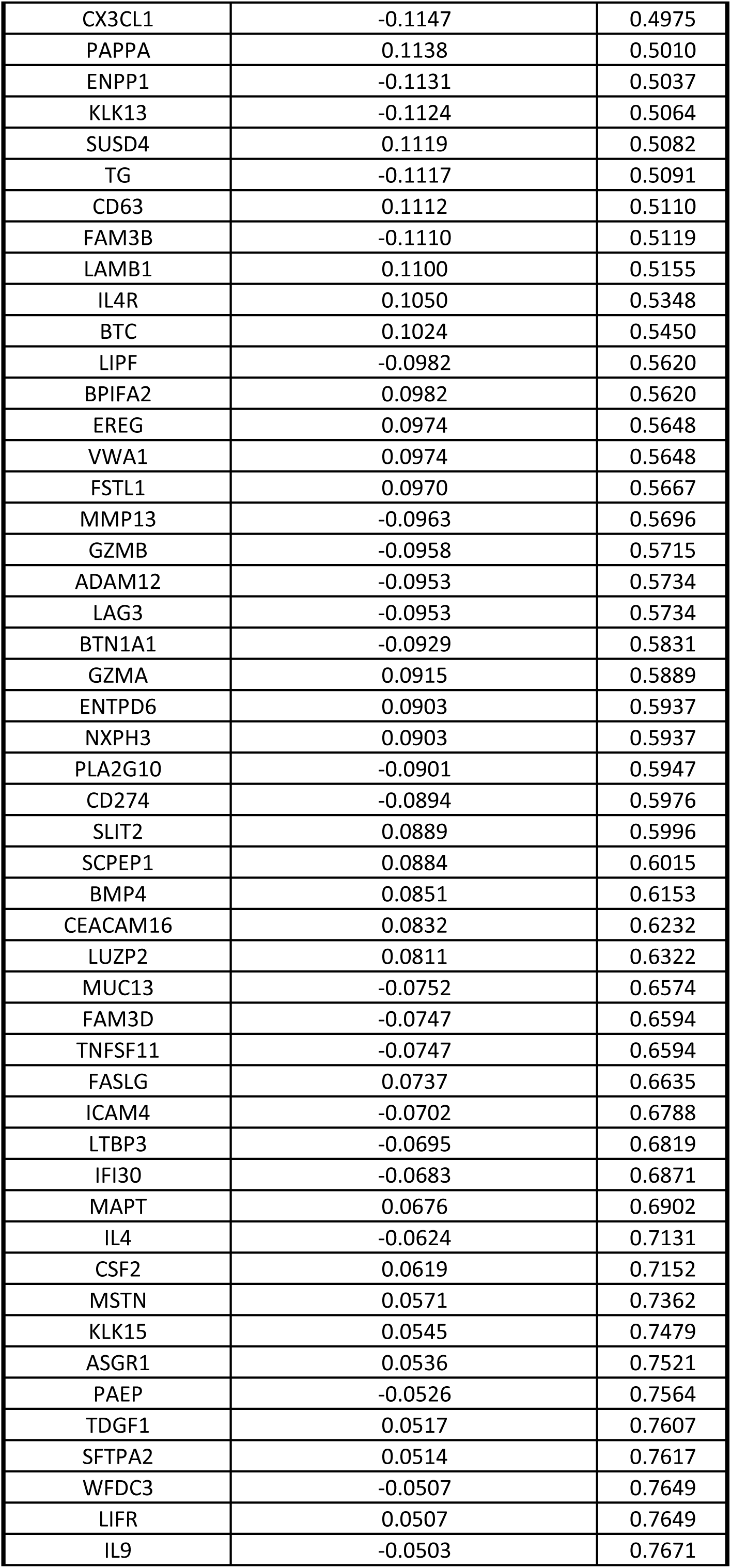
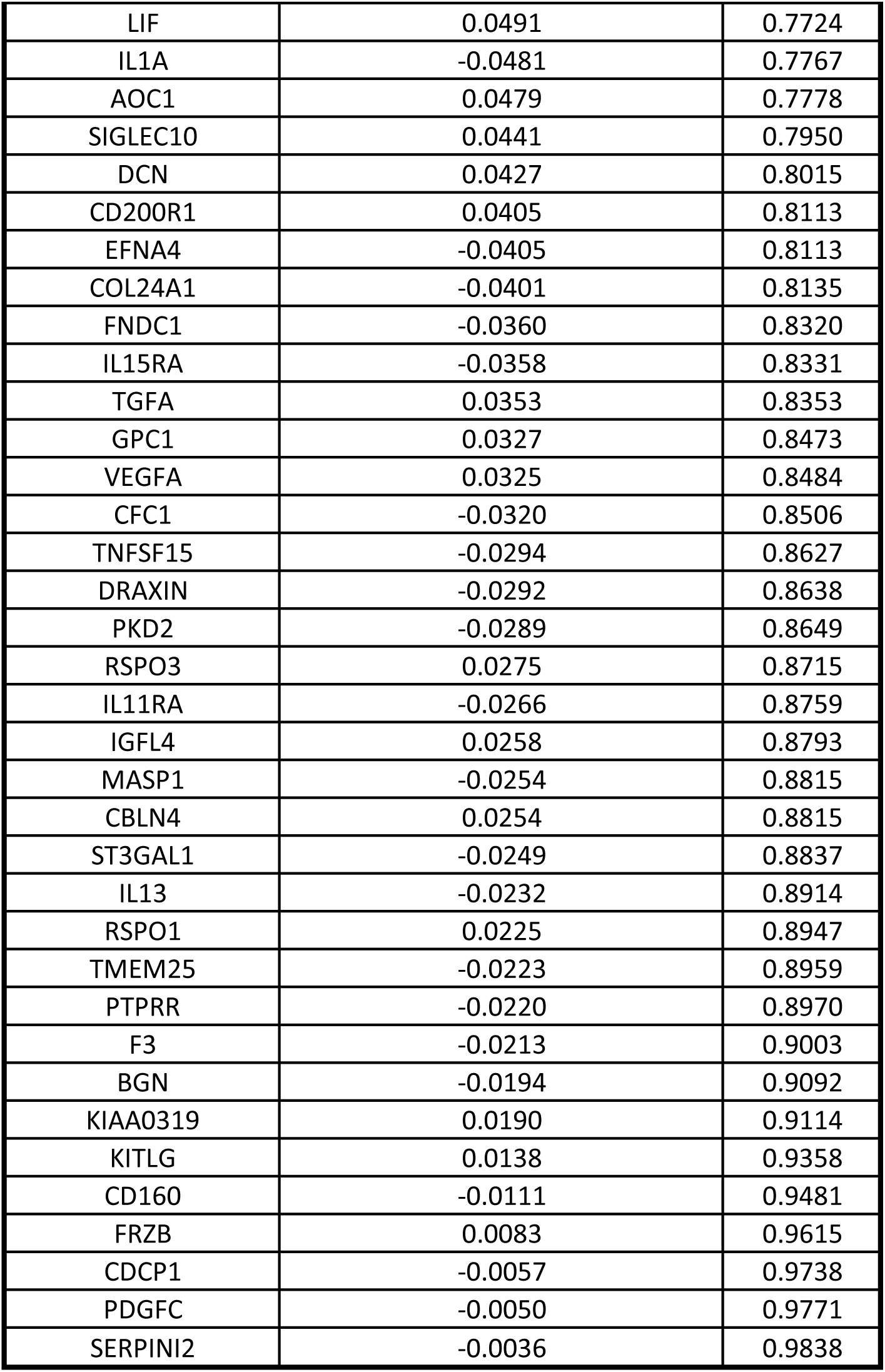
Correlation between serum PV glycan score and PV plasma proteins levels at baseline. Correlation coefficients were calculated by Spearman’s Correlation test

### Changes in circulating and cardiac serum levels of A2G2S2 N-glycan in AF

Peak 30 represented the most abundant glycan in human serum with a robust detection curve (**Fig. 1a**). It was upregulated in AF in both circulating PV and cardiac CS serum pools (**Fig. 1b-1c**) and contributed to the elevated levels of S2 and G2 derived traits (**Fig. 1e-1f**). Peak 30 was identified as the A2G2S2 *N*-glycan (**Fig. 4a** and **Table 9**). We found that baseline A2G2S2 abundance in the PV serum was comparable to that in CS pool (*P* = 0.2533, **Fig. 4b**). Moreover, PV and CS levels of A2G2S2 were positively correlated (*r* = 0.972, *P* < 0.0001, **Fig. 4c**), indicating that peripheral A2G2S2 largely reflects its cardiac levels. After five-fold cross-validation, the averaged AUROC for PV A2G2S2 (SR vs AF groups) was 0.7016 (± 0.1236 SD), while the averaged AUROC for CS A2G2S2 (non-AF vs AF groups) was 0.6539 (± 0.1685 SD) (**Fig. 4d**).

**Figure 4.**
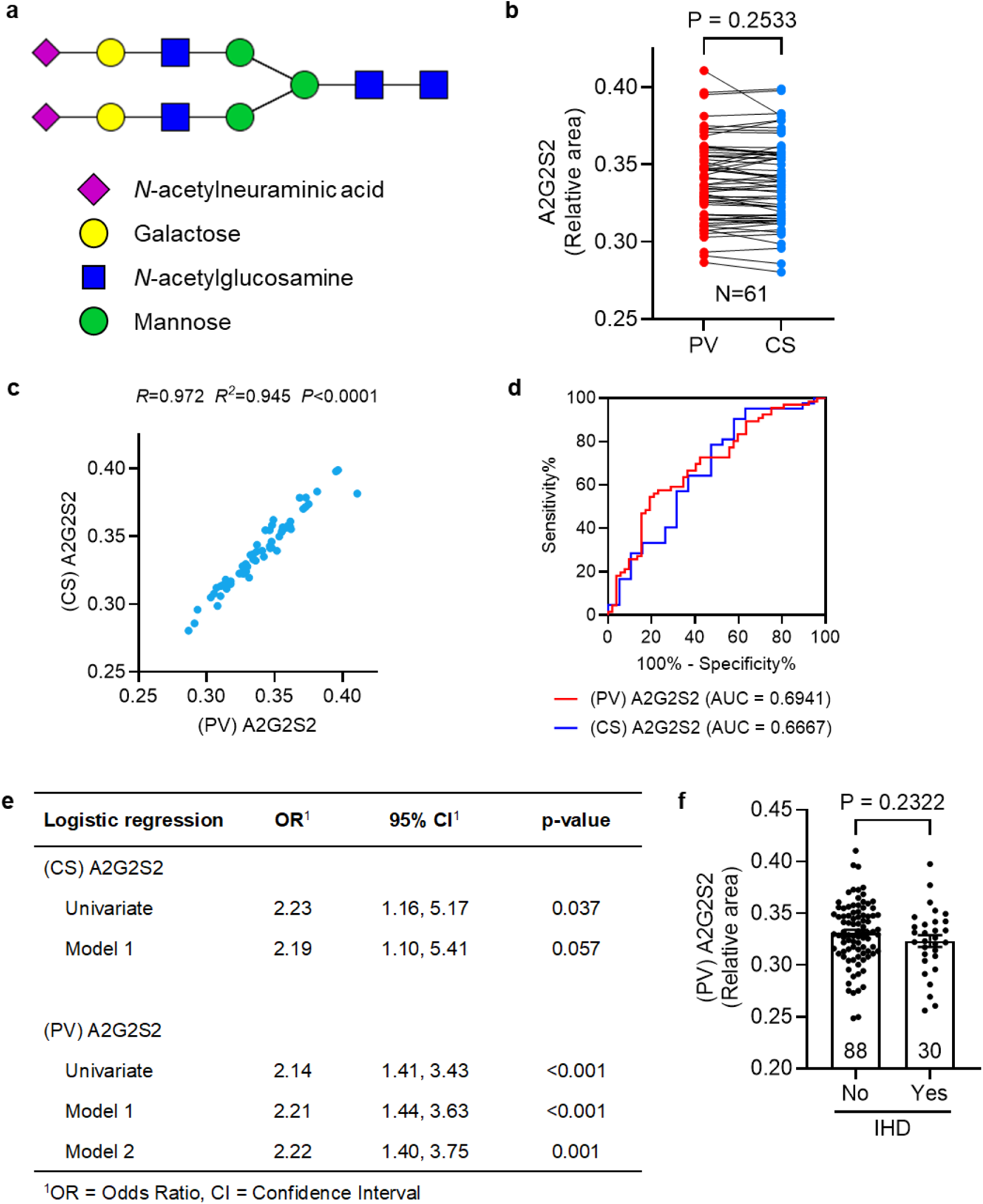
A2G2S2 glycan as a potential predictor of AF. (**a**) Illustration of peak 30 *N*-glycan composition and structure. (**b**) A2G2S2 level between PV and CS serum. (**c**) Correlation between PV A2G2S2 and CS A2G2S2 levels. (**d**) ROC curves of PV and CS A2G2S2 for predicting AF. (**e**) Univariate and multivariate logistic regression analysis for PV and CS A2G2S2 levels in association with AF. Results for model 1 are adjusted for age, sex and BMI. Results in model 2 are adjusted for age, sex, BMI, hypertension and IHD. (**f**) PV A2G2S2 levels in patients with and without IHD. Data are expressed as mean ± SEM (f); N represents individual biological donors (b, c, f). *P* values are determined by paired t-test (b), Pearson correlation test (c) and unpaired t-test (f). AF, atrial fibrillation; AUC, area under curve; BMI, body mass index; CS, coronary sinus; IHD, ischemic heart disease; PV, peripheral venous; ROC, receiver operating characteristic; SR, sinus rhythm.

**Table 9:**
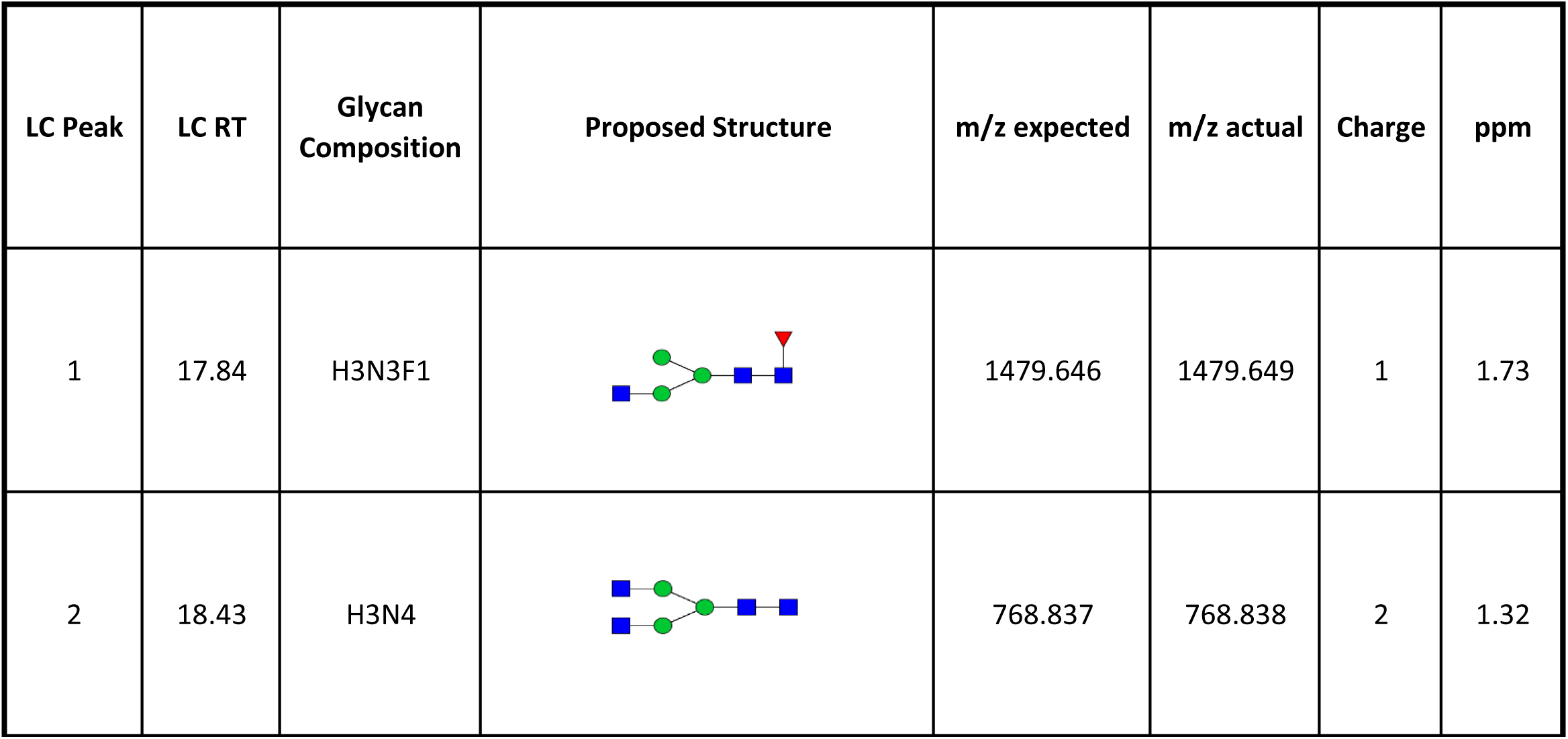
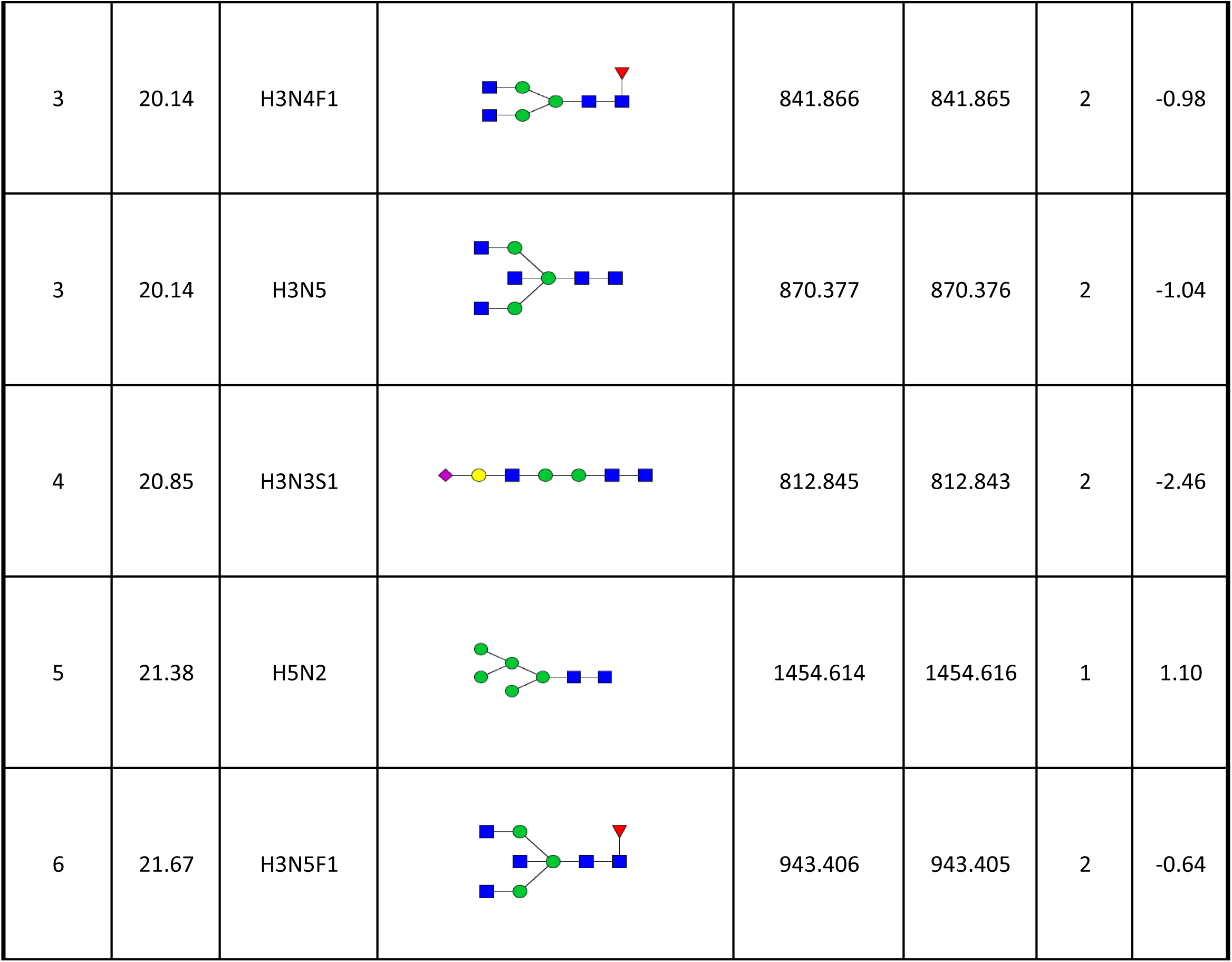
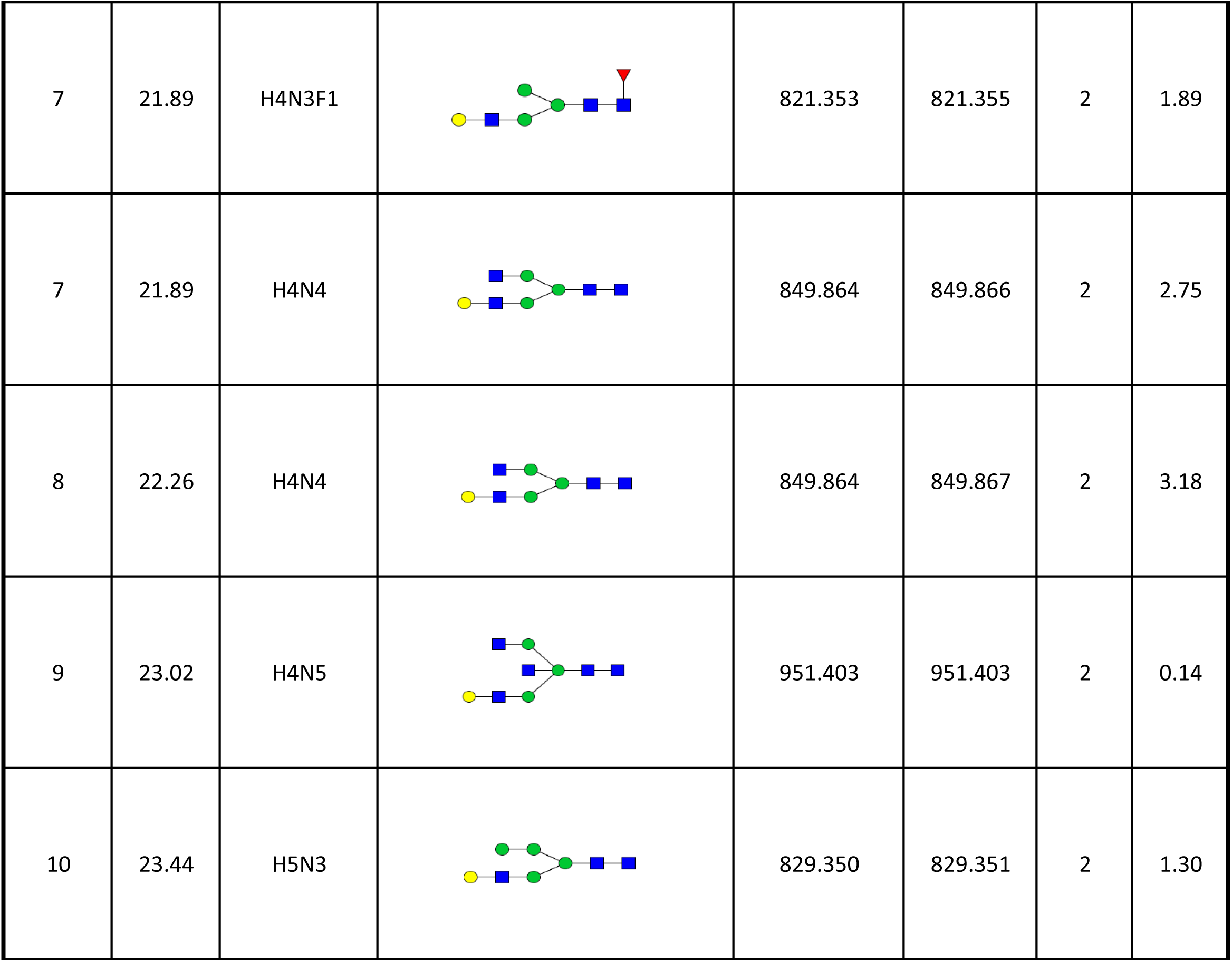
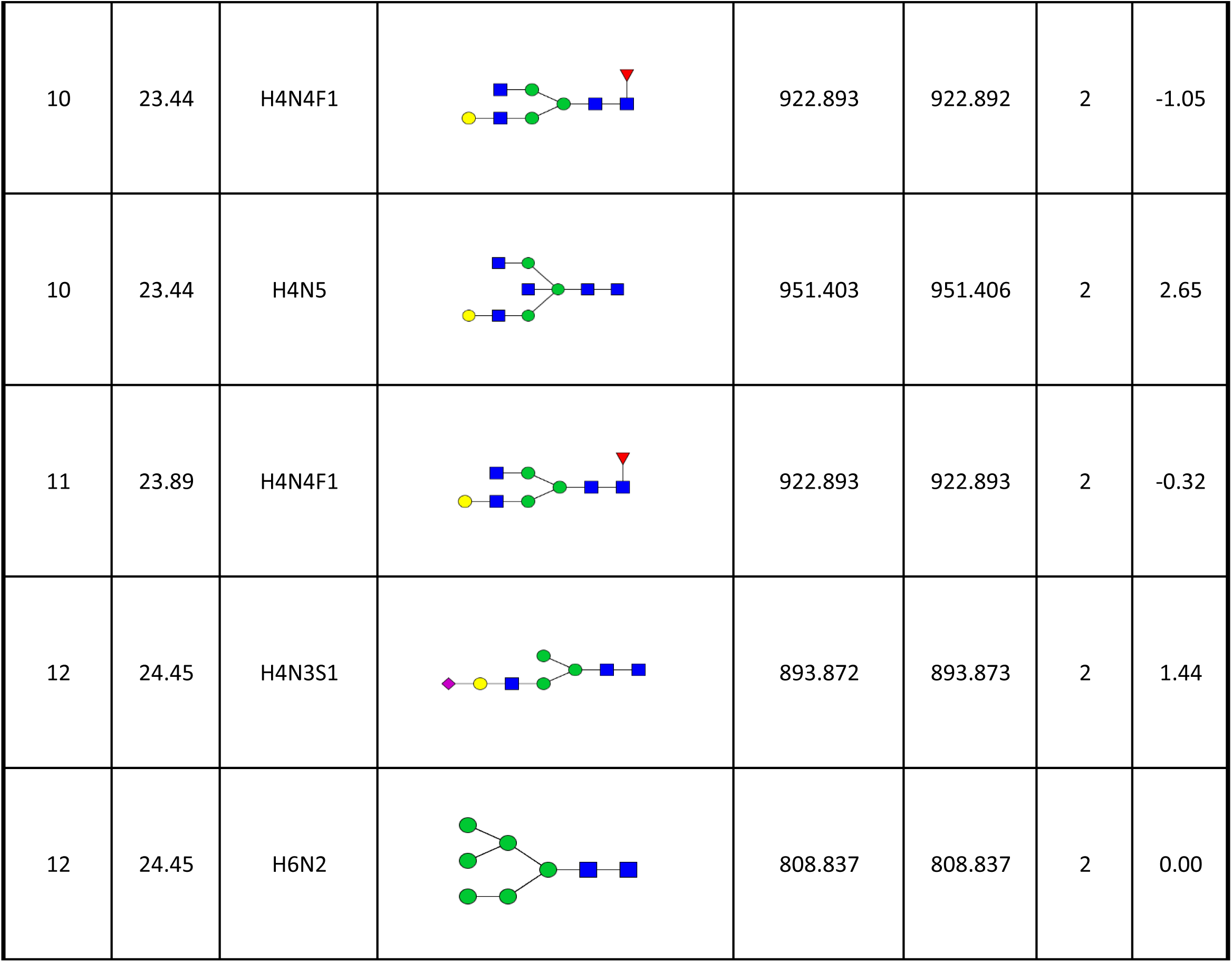
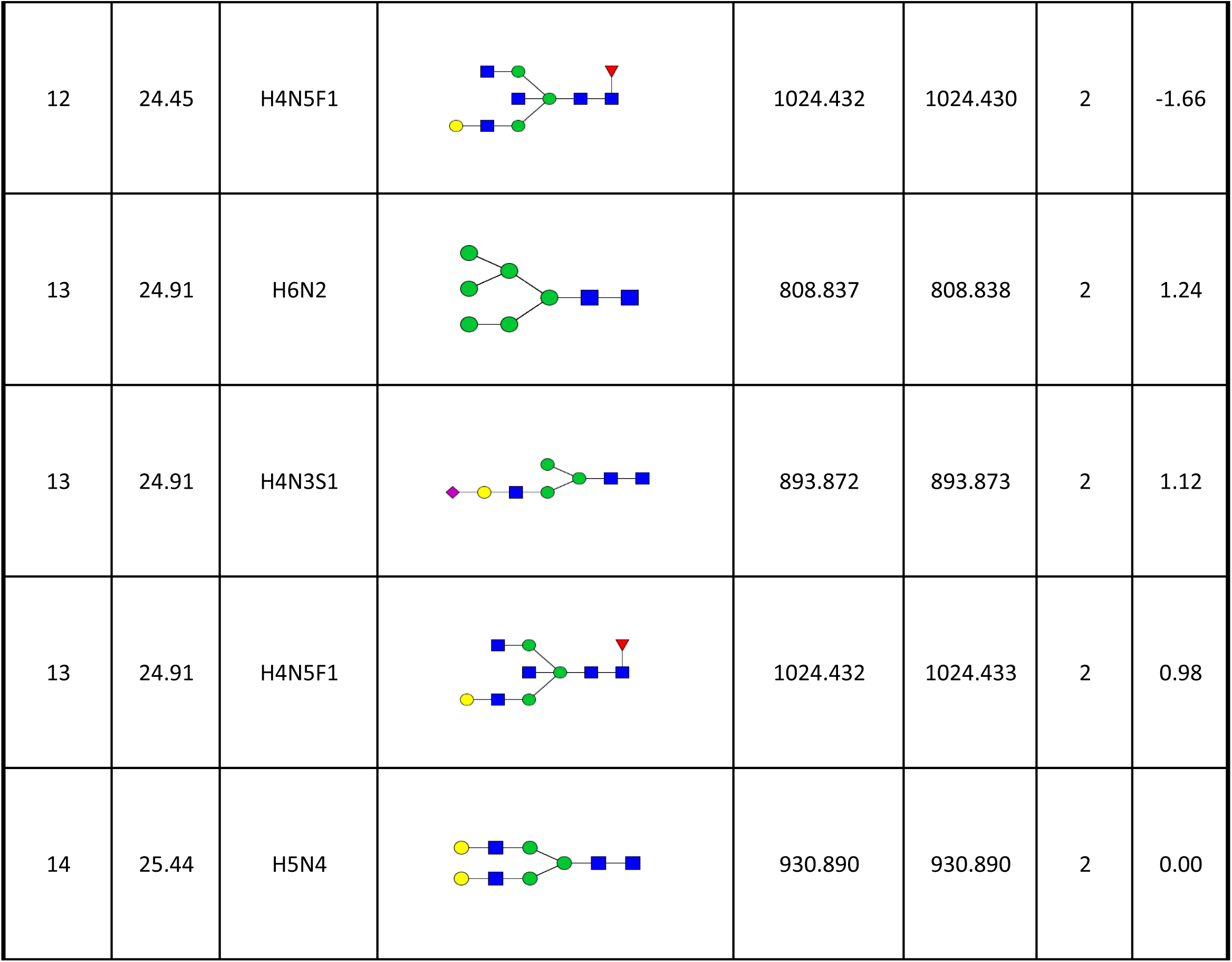
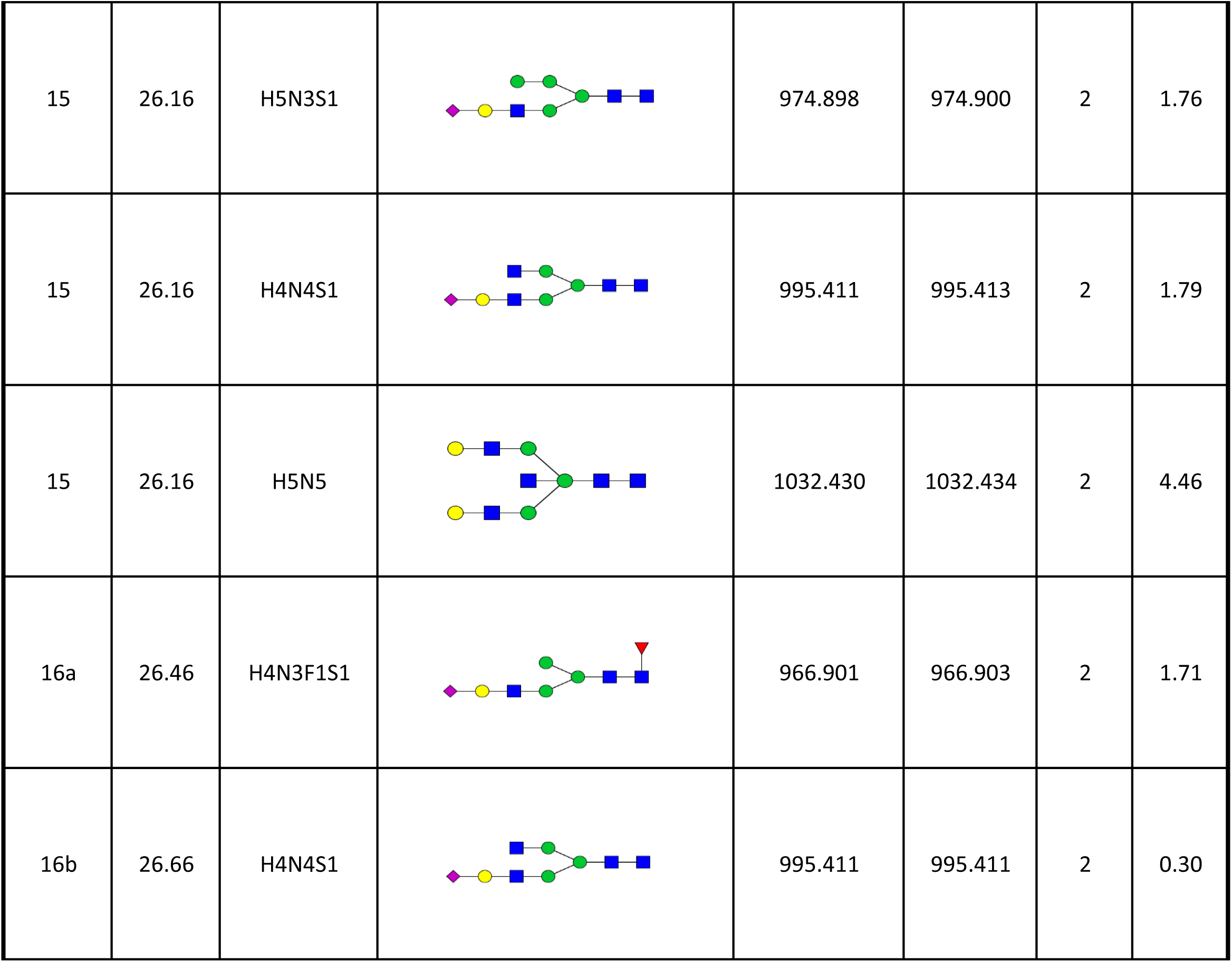
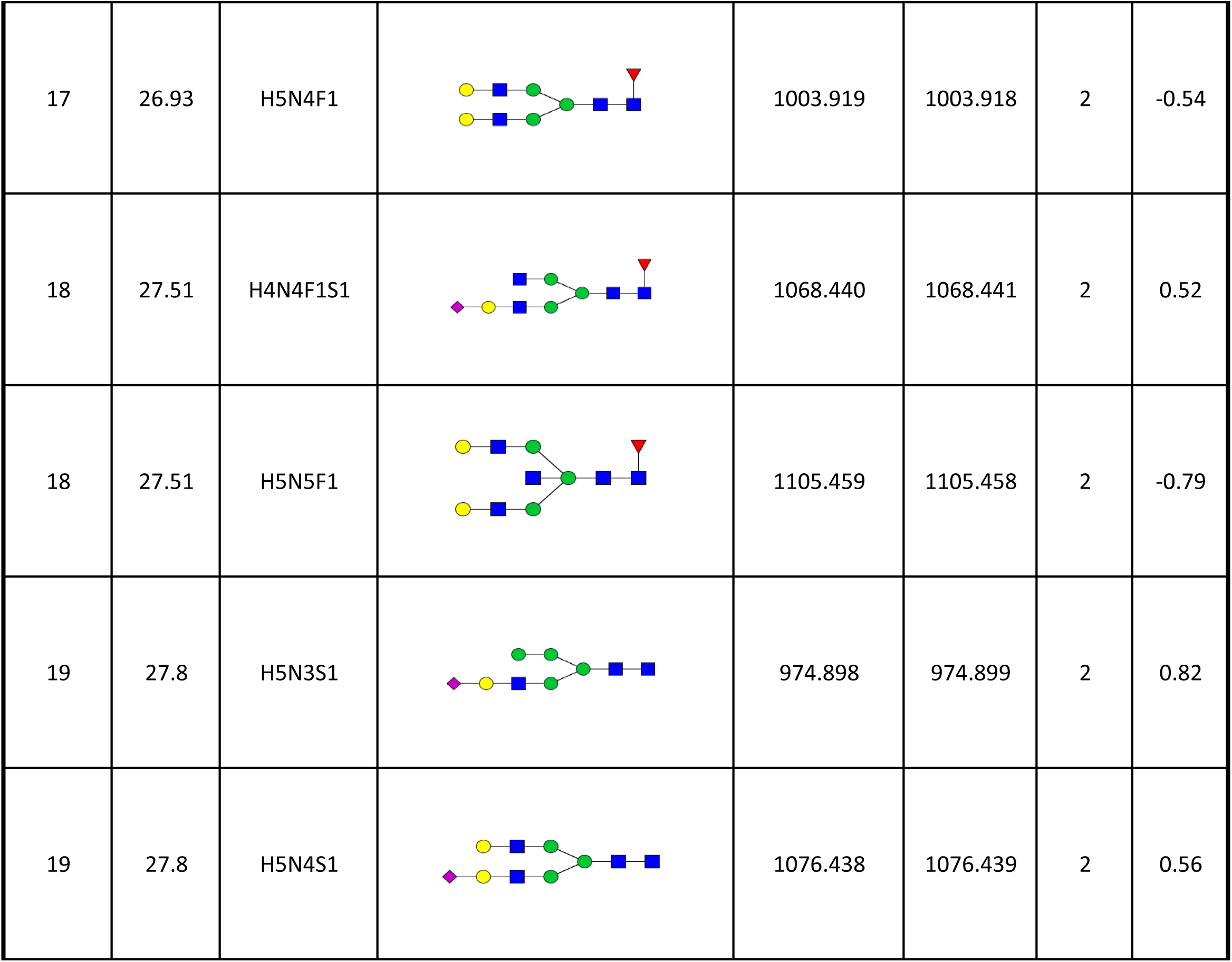
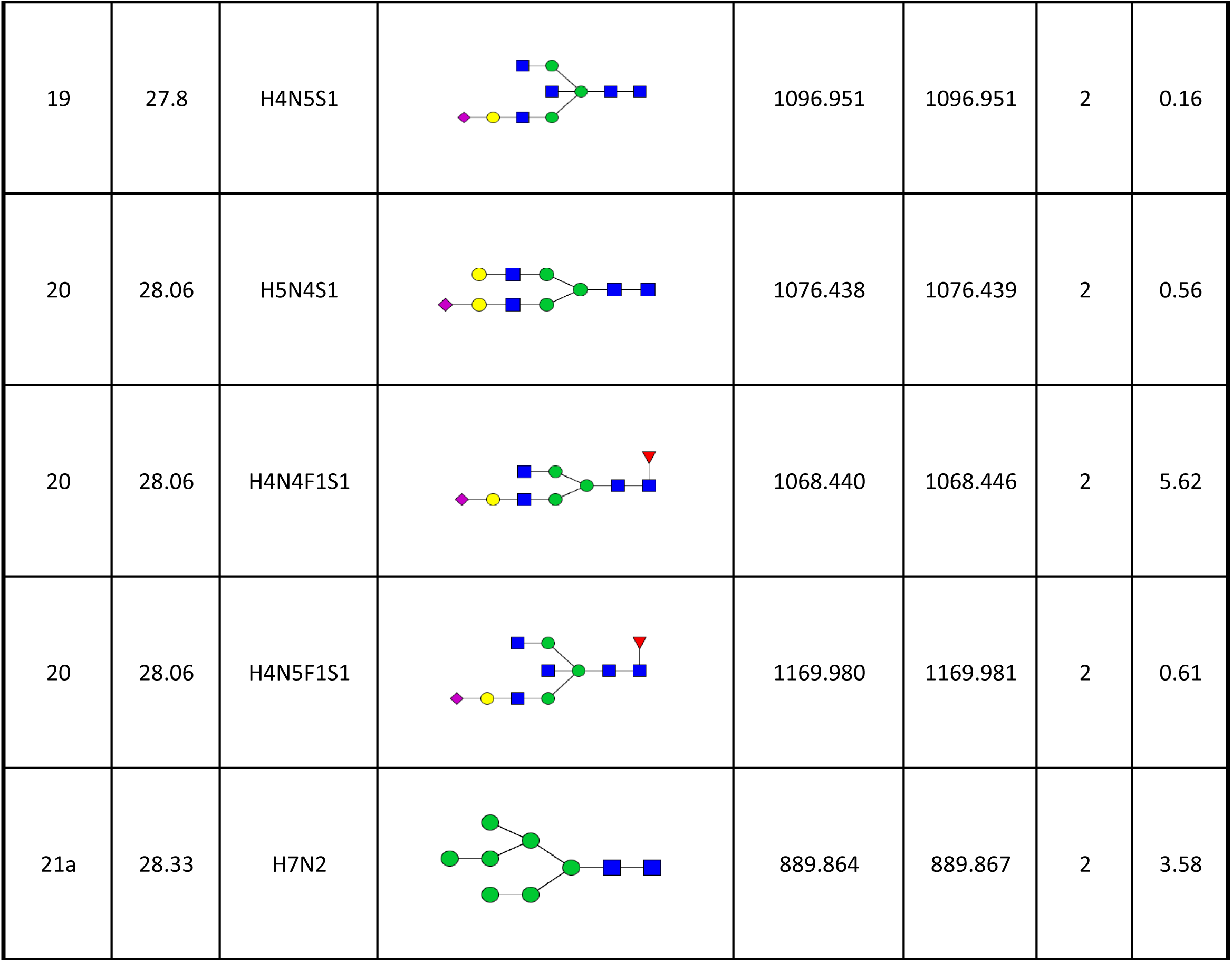
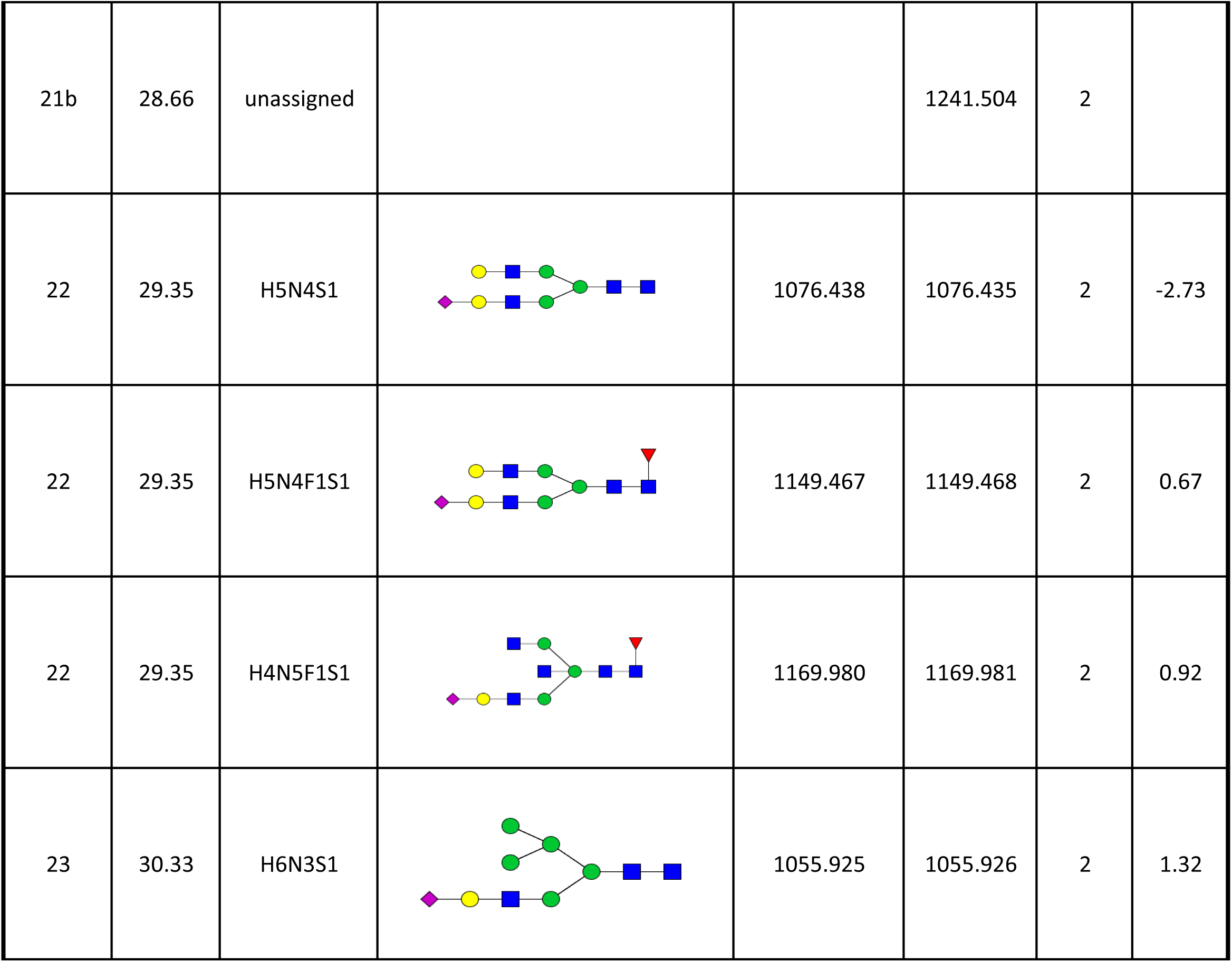
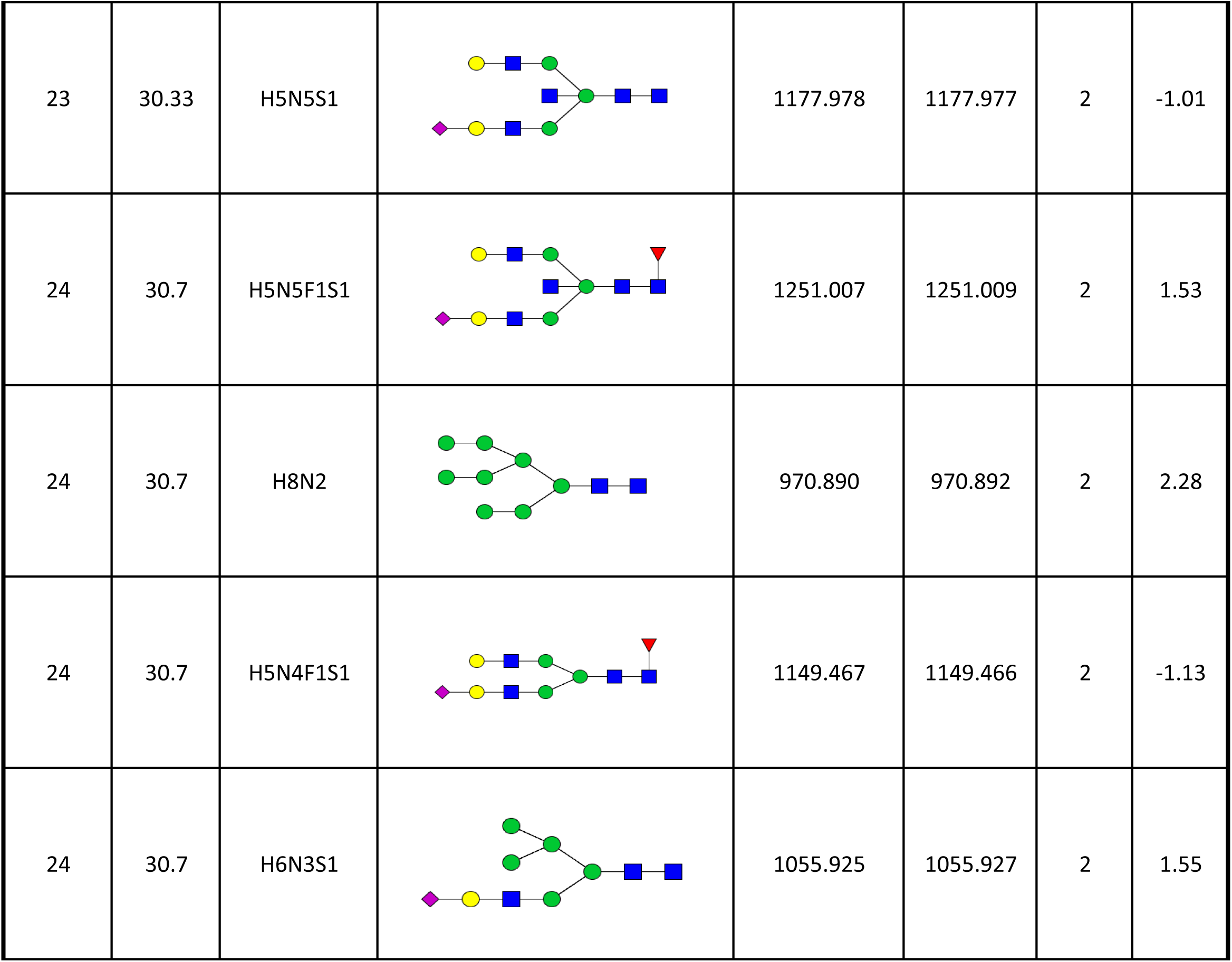
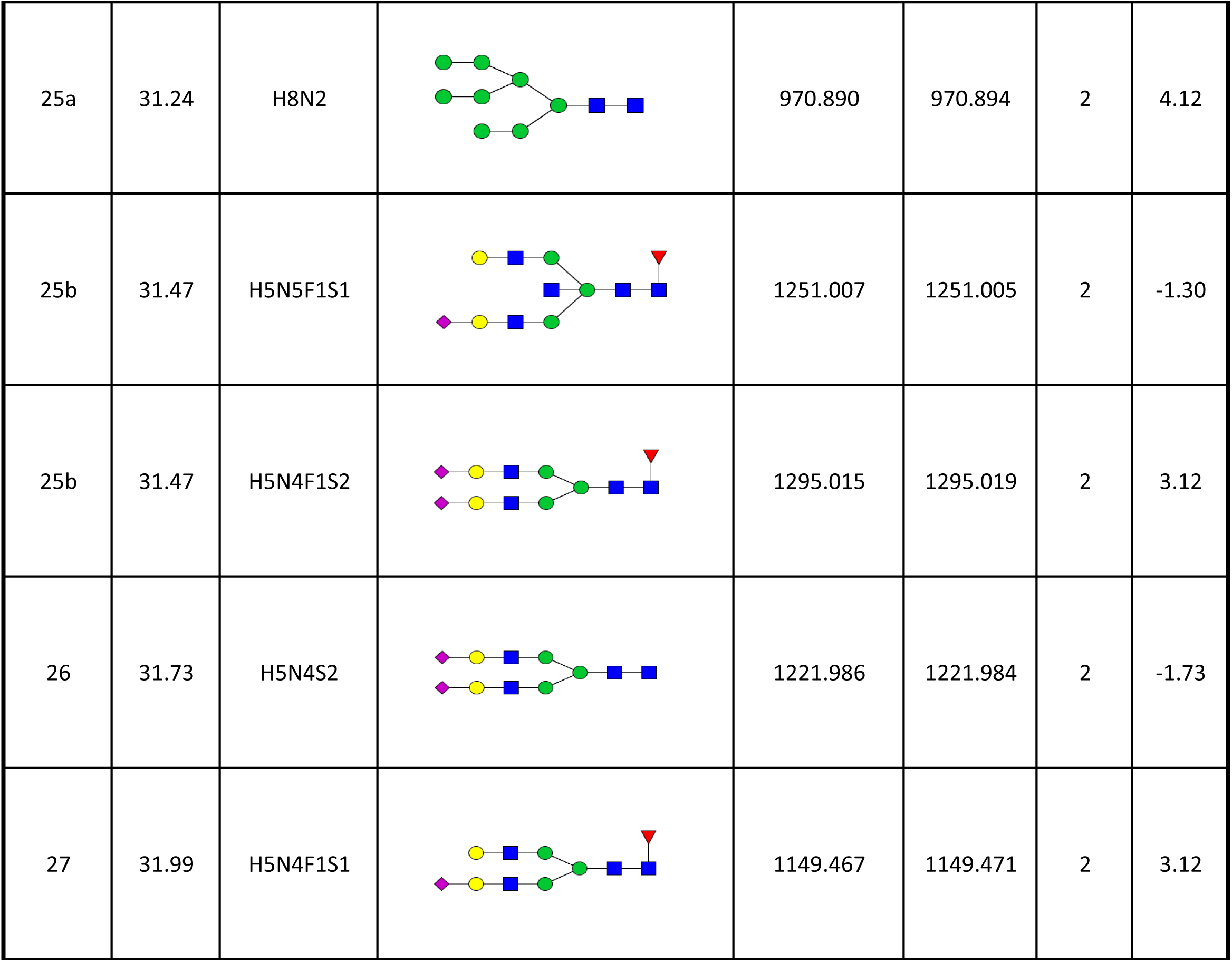
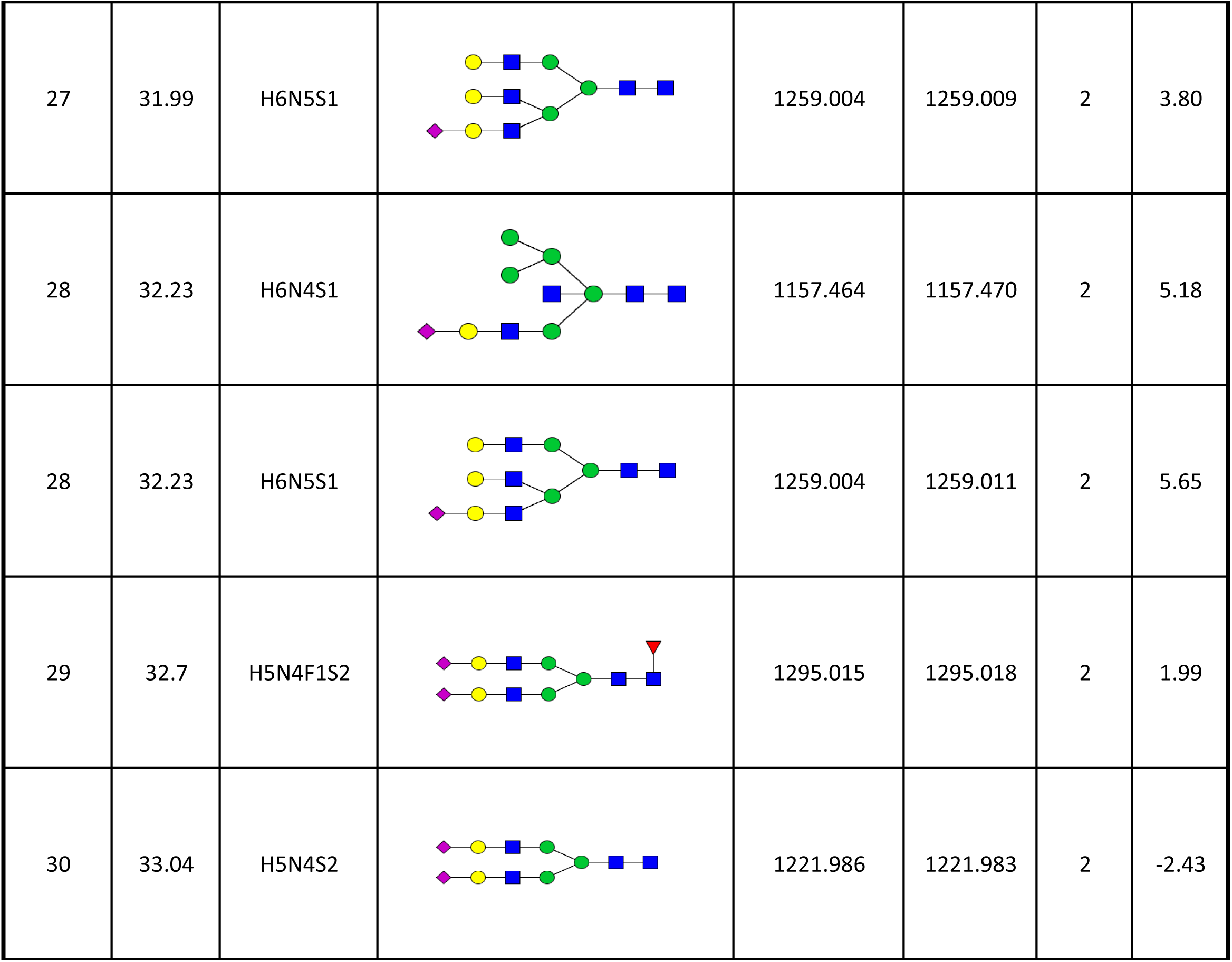
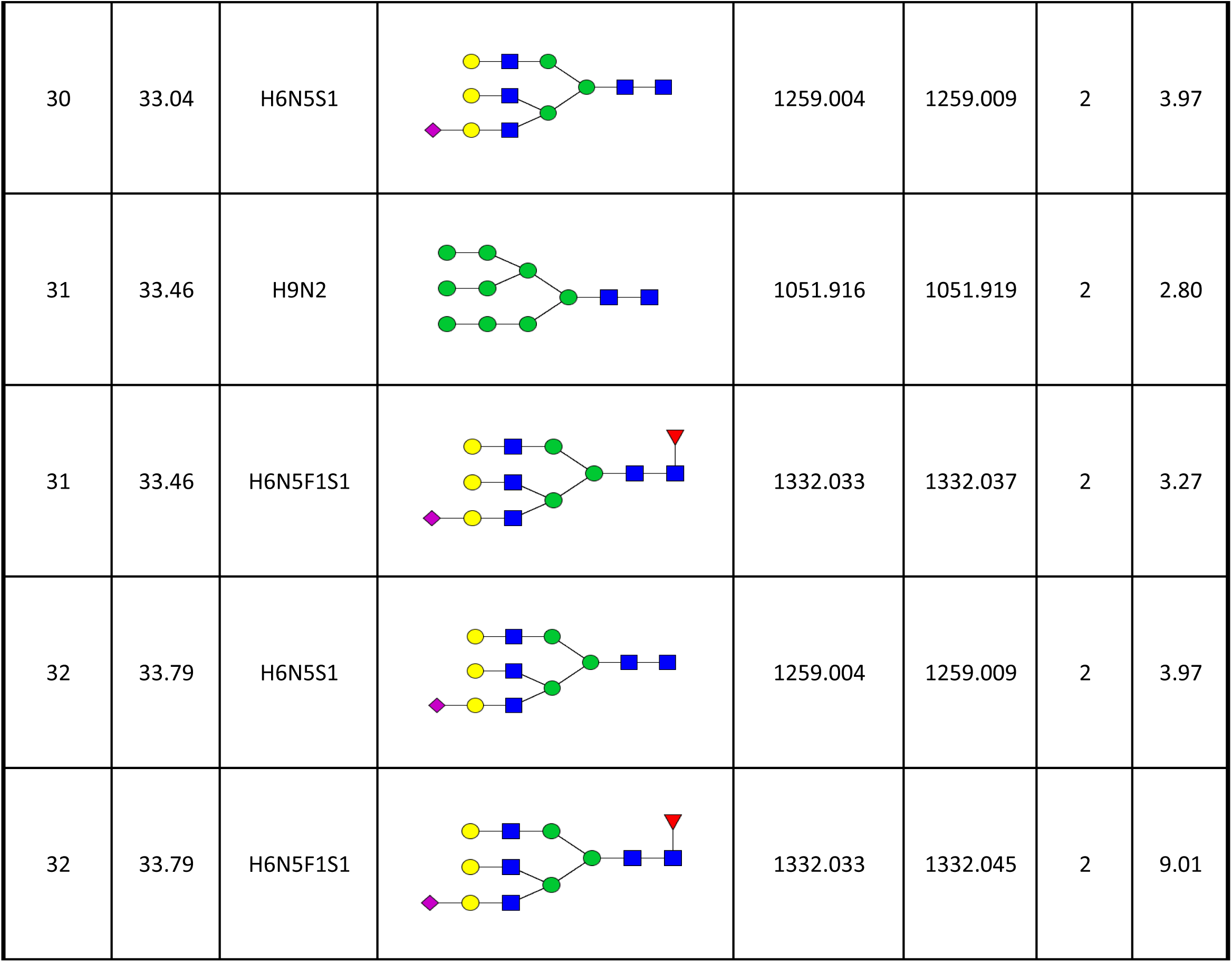
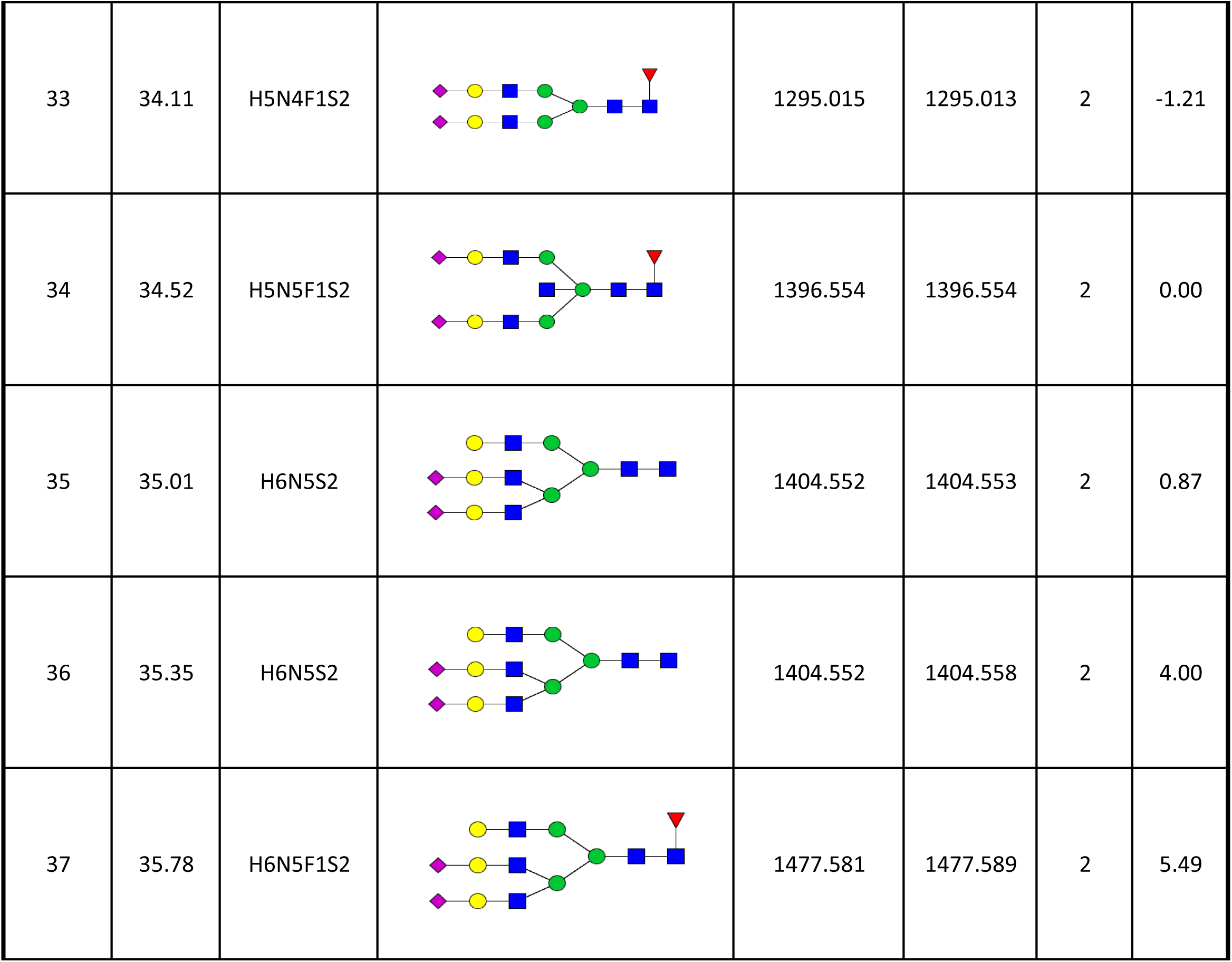
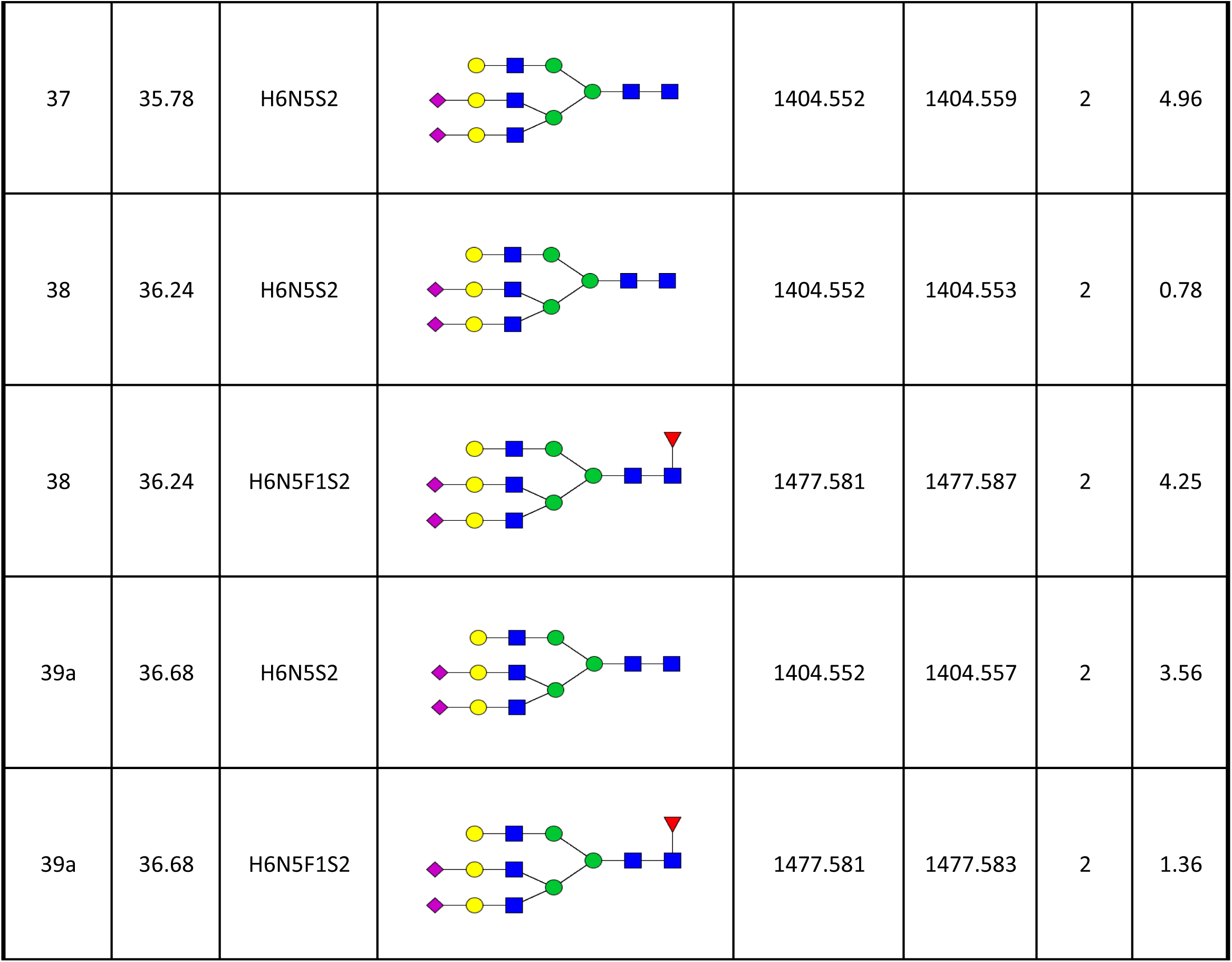
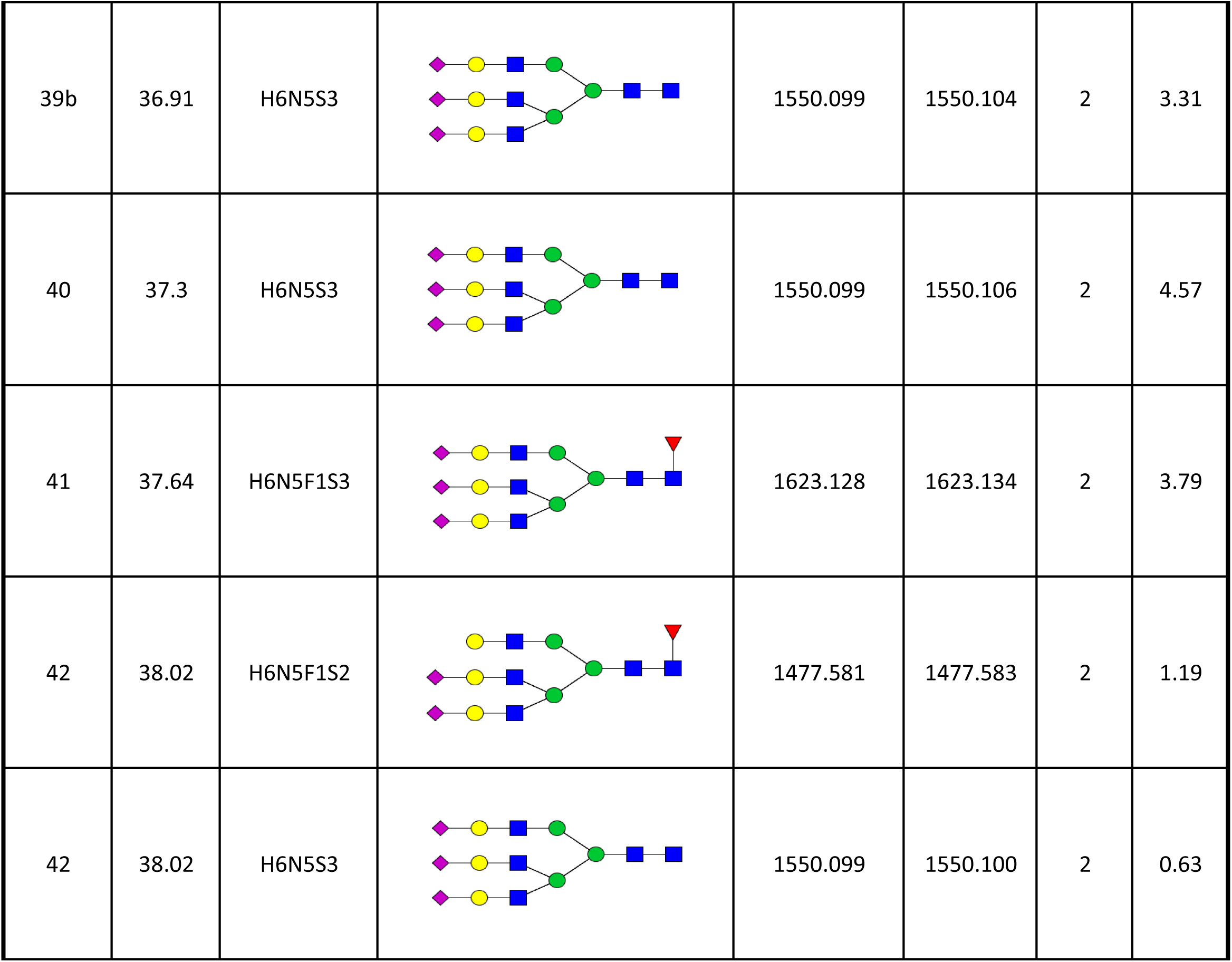
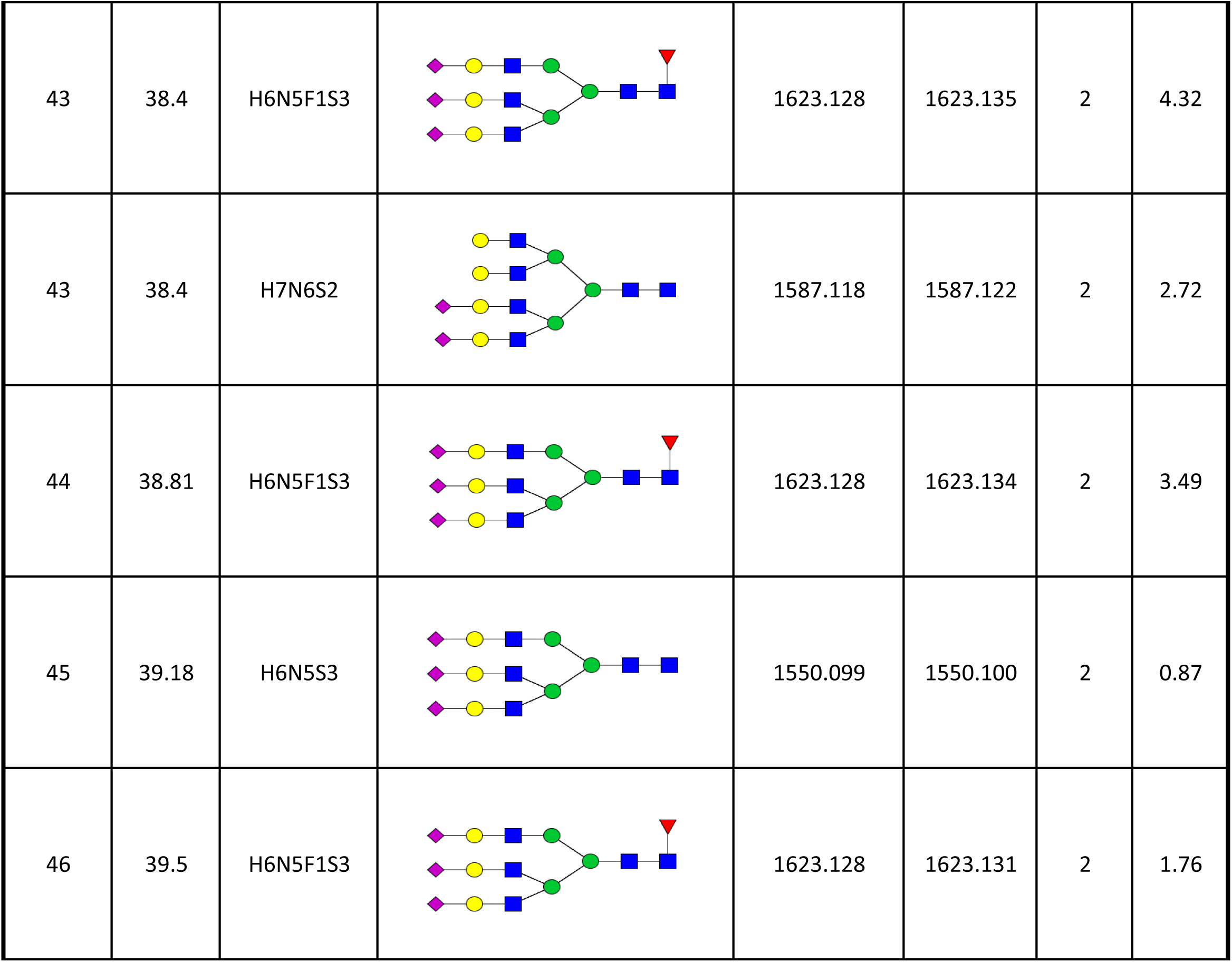
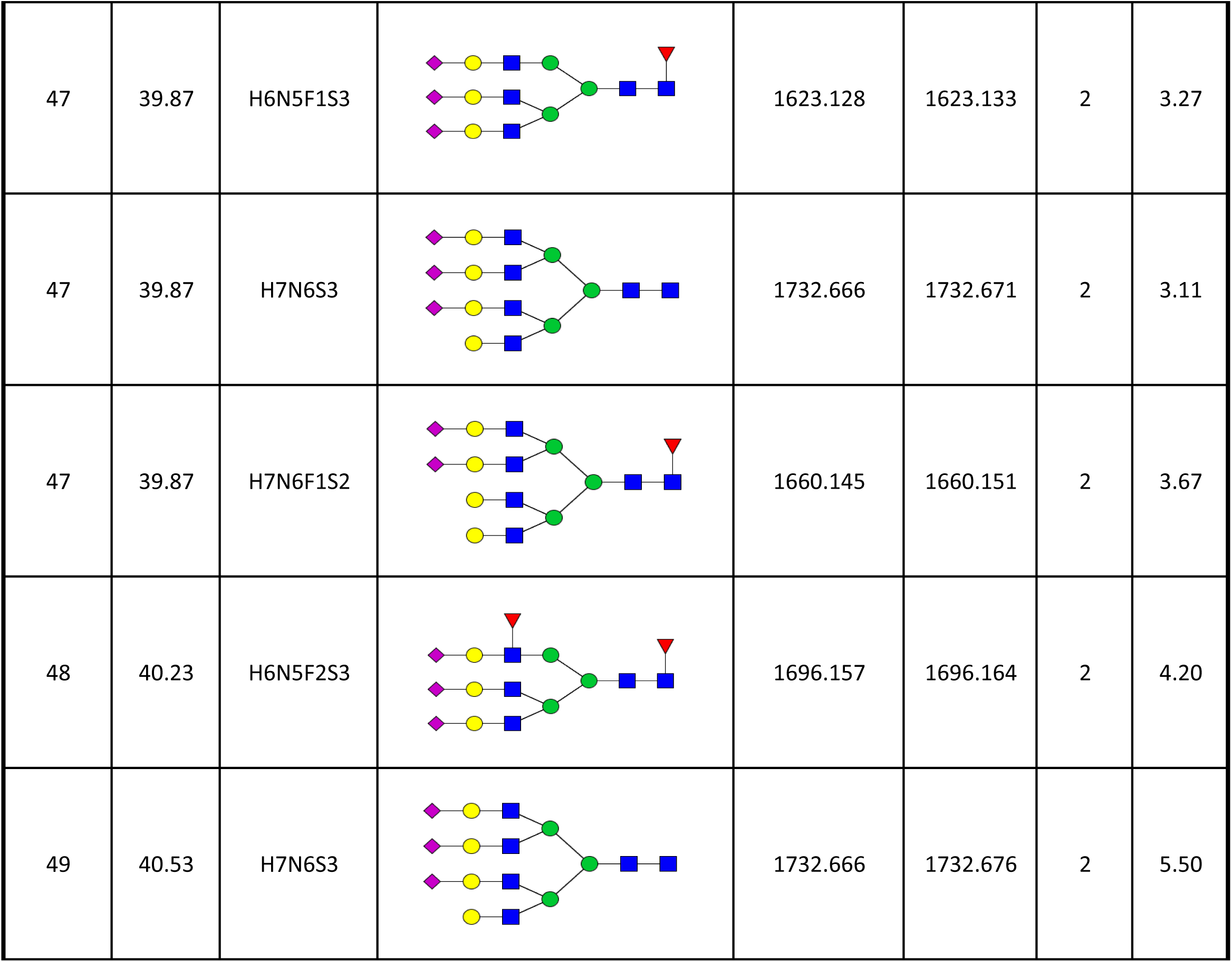
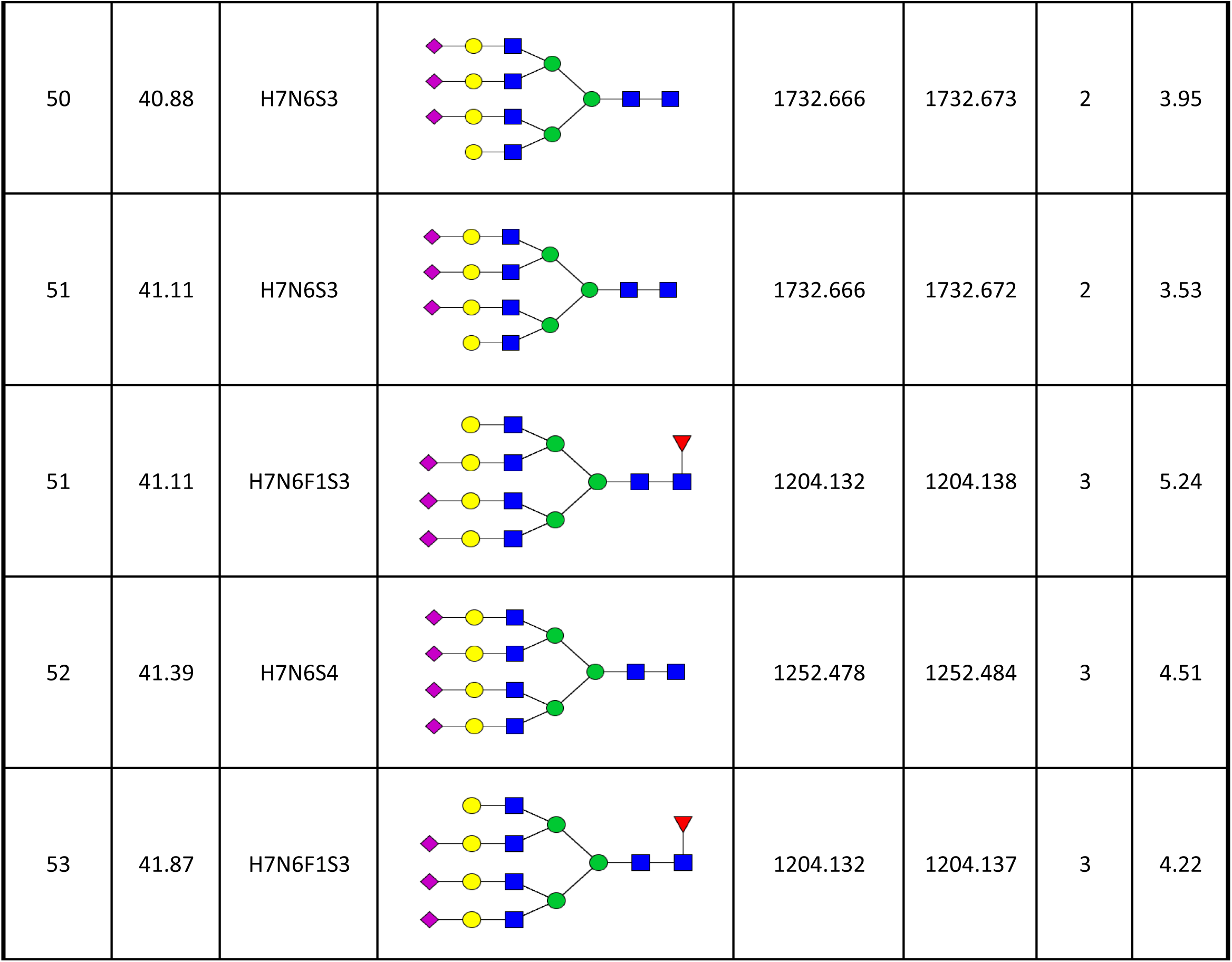
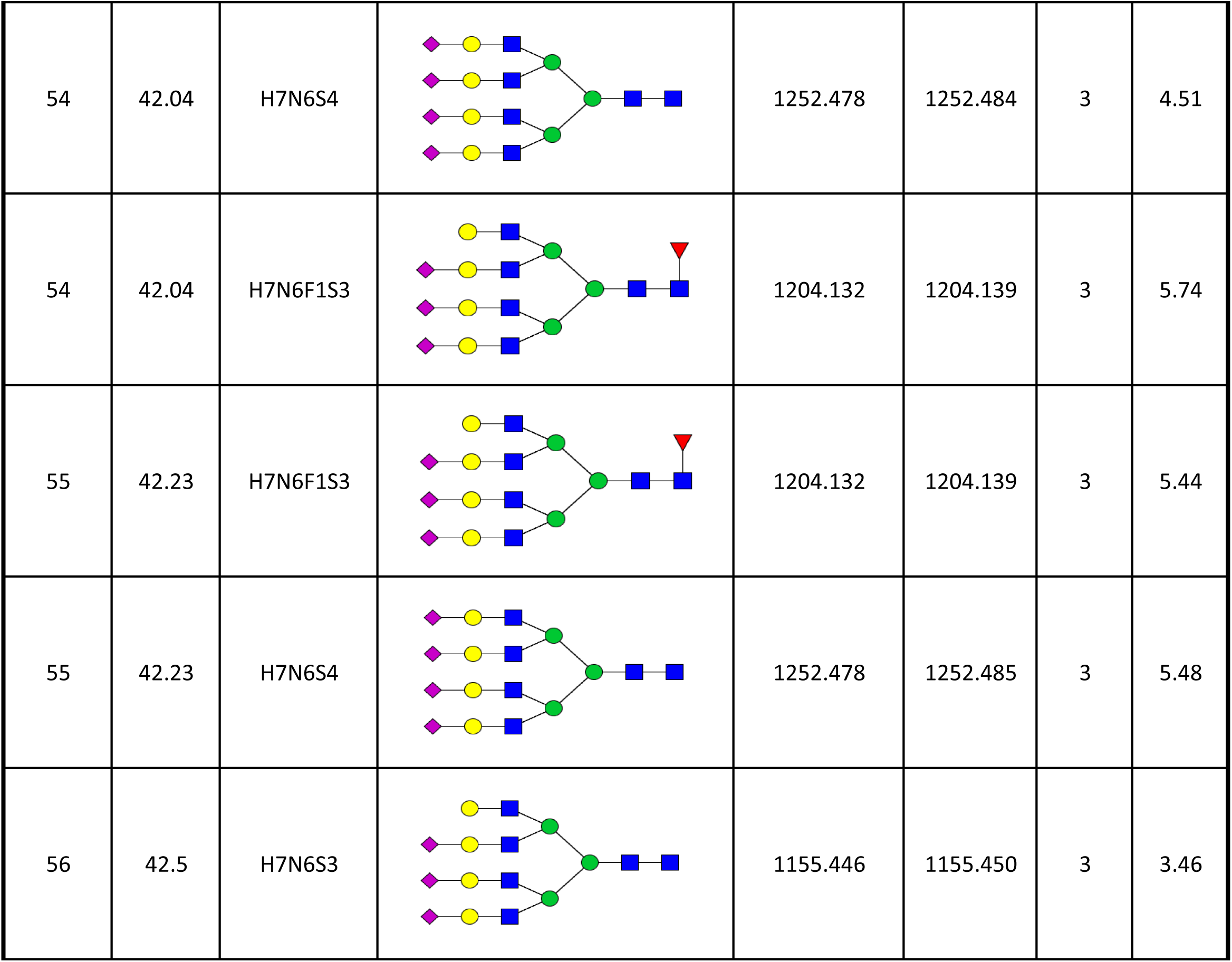
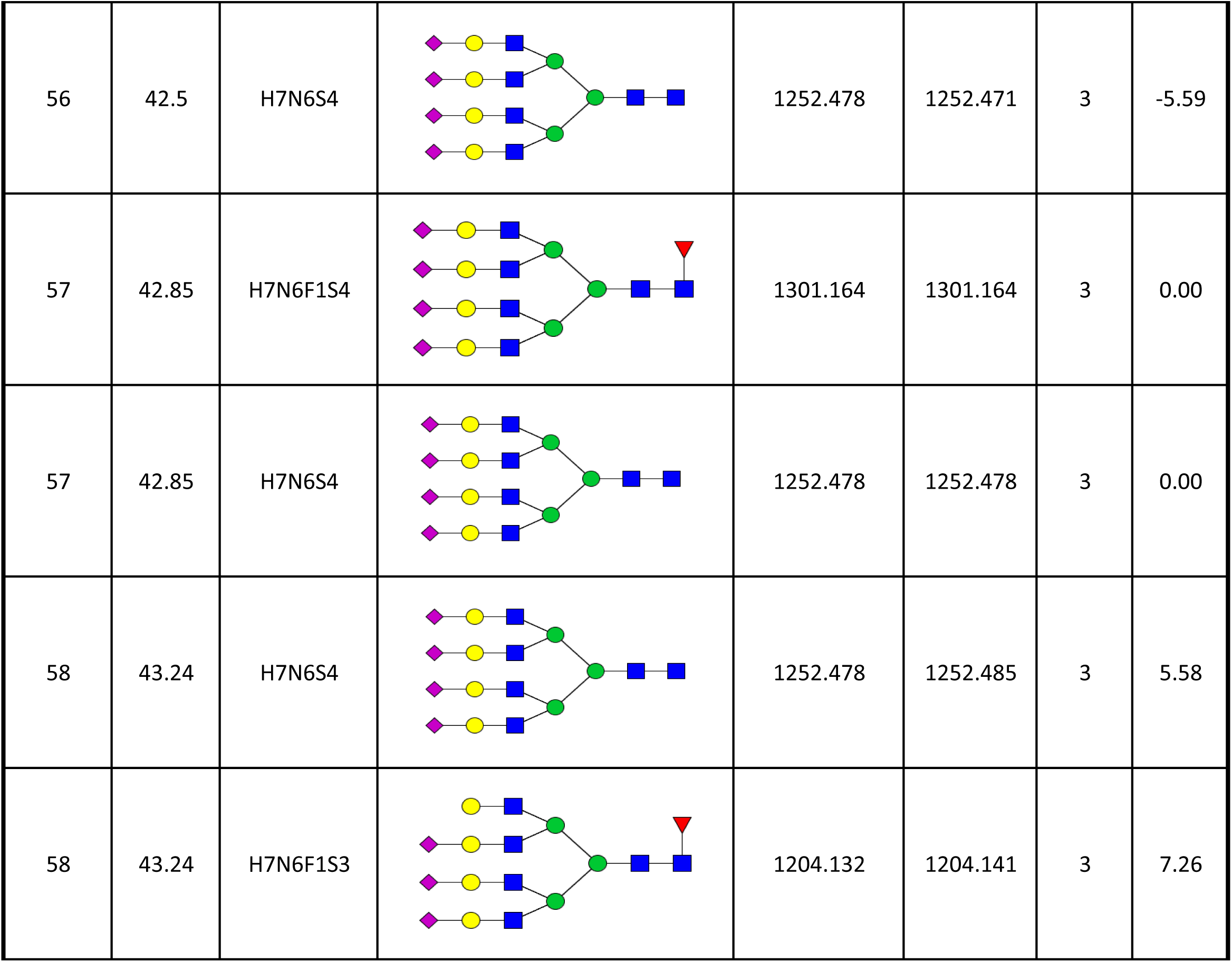
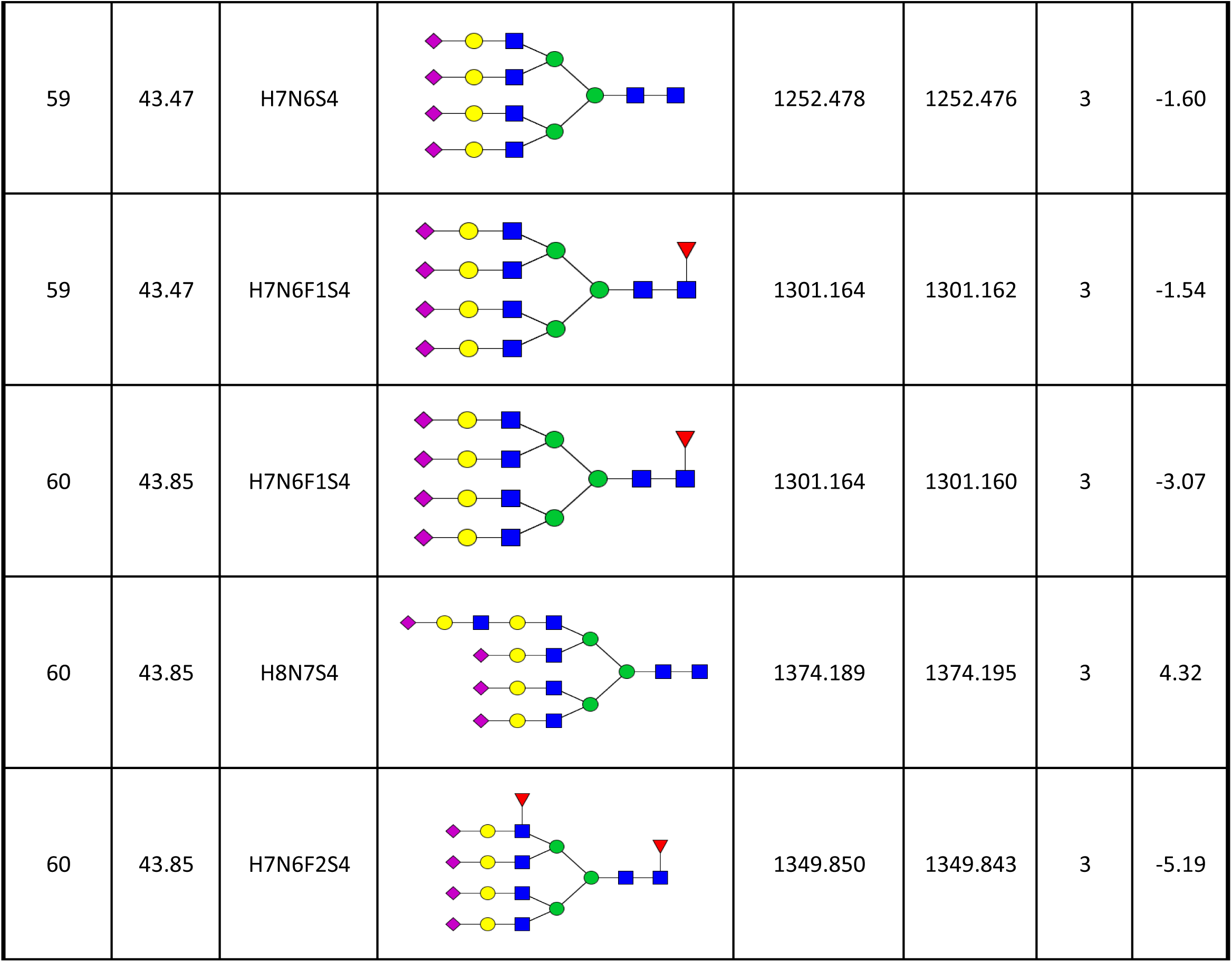
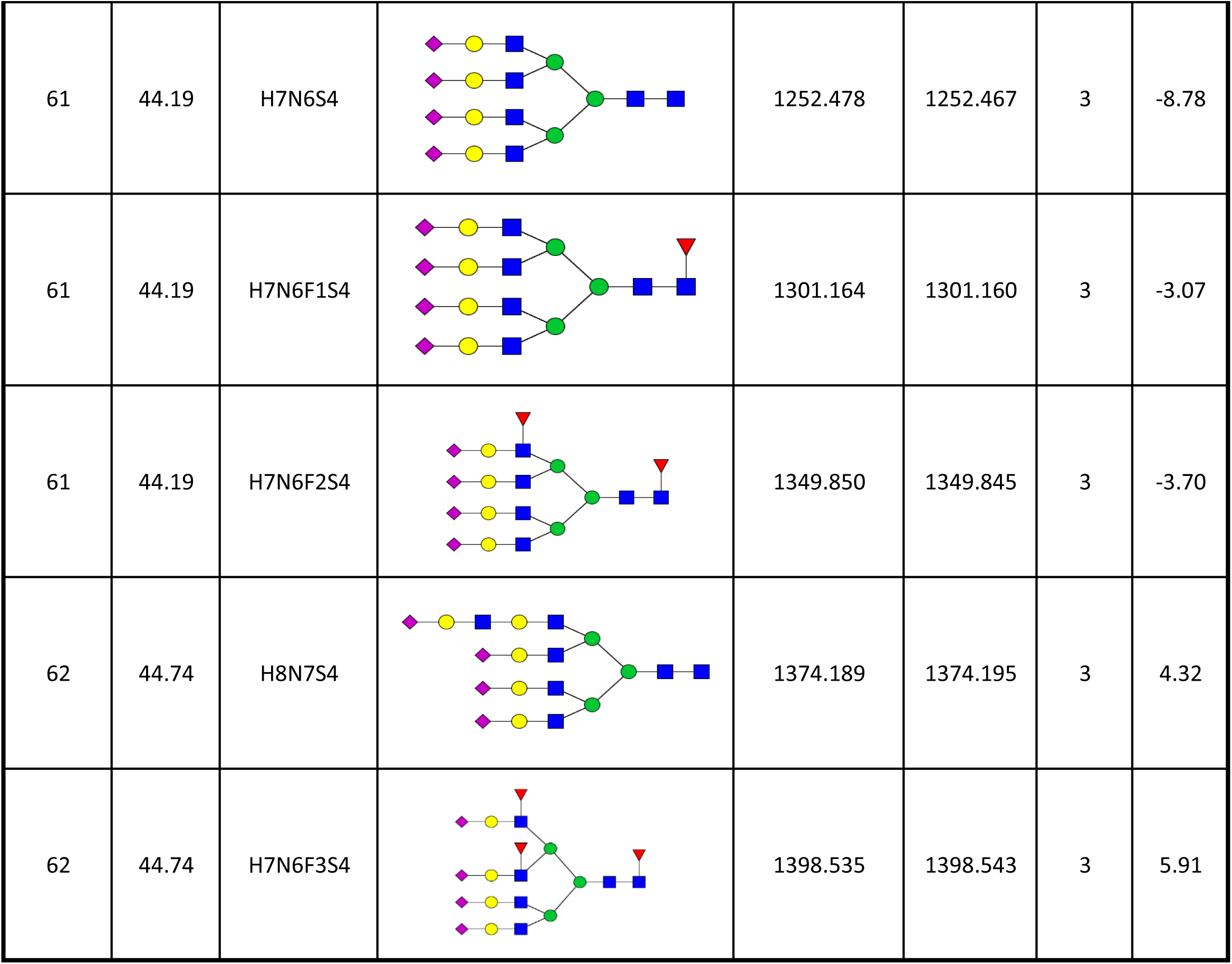
Alignment of the HPLC-derived glycan peaks (LC peaks) with LC-MS-derived glycans for total serum *N* -glycans. Structures were confirmed through consideration of the expected and actual M/Z, coupled with the ppm and comparison of retention times across LC (LC RT) and MS machinery. The proposed structures of glycans are represented, with CFG notation as shown in the methodology section. Glycan names are represented by their components, including abbreveations: hexose (H), N-acetylhexosamine (N), fucose (F), N-actylneuraminicic acid (S). LC, liquid chromatography; MS, mass spectrometry; RT, retention time

Next, logistic regression analysis showed that baseline PV A2G2S2 (OR 2.14, 95% CI: 1.41-3.43; *P* < 0.001) and CS A2G2S2 (OR 2.23, 95% CI: 1.16-5.17; *P* = 0.037) were both associated with higher odds of AF (**Fig. 4e** and **Table 10-11**). After adjusting for clinical risk factors and confounders, only PV A2G2S2 remained associated with increased odds of developing AF (OR 2.22, 95% CI: 1.40-3.75; *P* = 0.001) (**Fig. 4e** and **Table 10-11**). Although IHD was linked to lower odds of AF (OR 0.16, 95% CI: 0.05-0.45, *P* < 0.0001) (**Table 10**), PV A2G2S2 levels did not differ between patients with and without IHD (**Fig. 4f**). Moreover, variance inflation factor (VIF) assessed for the adjusted models revealed no multicollinearity (**Table 12**).

**Table 10:** Logistic regression analysis for (PV) A2G2S2 association with AF. Model 1 adjusted for age, sex and BMI Model 2 adjusted for age, sex, BMI, hypertension, IHD BMI, body mass index; CI, confidence interval; IHD, ischemic heart disease; PV, peripheral vein; OR, odds ratio

| Univariate |  |  |  | Adjusted Model 1 |  |  |  | Adjusted Model 2 |  |  |  |
| --- | --- | --- | --- | --- | --- | --- | --- | --- | --- | --- | --- |
| Characteristics | OR<sup>2</sup> | 95% CI<sup>2</sup> | P-value | Characteristics | OR<sup>2</sup> | 95% CI<sup>2</sup> | P-value | Characteristics | OR<sup>2</sup> | 95% CI<sup>2</sup> | P-value |
| (PV) A2G2S2 | 2.14 | 1.41, 3.43 | <0.001 | (PV) A2G2S2 | 2.21 | 1.44, 3.63 | <0.001 | (PV) A2G2S2 | 2.22 | 1.40, 3.75 | 0.001 |
|  |  |  |  | Age | 0.98 | 0.94, 1.02 | 0.309 | Age | 0.99 | 0.95, 1.04 | 0.713 |
|  |  |  |  | Sex (female) | 1.06 | 0.45, 2.55 | 0.890 | Sex (female) | 0.79 | 0.31, 2.05 | 0.632 |
|  |  |  |  | BMI | 0.97 | 0.89, 1.05 | 0.435 | BMI | 0.98 | 0.90, 1.07 | 0.659 |
|  |  |  |  |  |  |  |  | Hypertension | 0.63 | 0.26, 1.56 | 0.319 |
|  |  |  |  |  |  |  |  | IHD | 0.16 | 0.05, 0.45 | <0.001 |

**Table 11:** Logistic regression analysis for (CS) A2G2S2 association with AF. Model 1 adjusted for age, sex and BMI BMI, body mass index; CI, confidence interval; CS, coronary sinus; IHD, ischemic heart disease; OR, odds ratio

| Univariate |  |  |  | Adjusted Model 1 |  |  |  |
| --- | --- | --- | --- | --- | --- | --- | --- |
| Characteristics | OR<sup>2</sup> | 95% CI<sup>2</sup> | P-value | Characteristics | OR<sup>2</sup> | 95% CI<sup>2</sup> | P-value |
| (CS) A2G2S2 | 2.23 | 1.16, 5.17 | 0.037 | (CS) A2G2S2 | 2.19 | 1.10, 5.41 | 0.057 |
|  |  |  |  | Age | 1.05 | 1.00, 1.10 | 0.056 |
|  |  |  |  | Sex (female) | 1.01 | 0.29, 3.75 | 0.985 |
|  |  |  |  | BMI | 0.99 | 0.90, 1.06 | 0.730 |

**Table 12:** Multicollinearity check for adjusted logistic regression models. BMI, body mass index; CS, coronary sinus; IHD, ischemic heart disease; PV, peripheral vein; VIF, variance inflation factor

| Peripheral |  |  | Coronary |  |
| --- | --- | --- | --- | --- |
| Characteristics | VIF (Adjusted Model 1) | VIF (Adjusted Model 2) | Characteristics | VIF (Adjusted Model 1) |
| (PV) A2G2S2 | 1.0664 | 1.0682 | (CS) A2G2S2 | 1.0484 |
| Age | 1.0312 | 1.0752 | Age | 1.0210 |
| Sex | 1.0156 | 1.0544 | Sex | 1.0179 |
| BMI | 1.1113 | 1.1209 | BMI | 1.0603 |
| Hypertension | N/A | 1.1139 |  |  |
| IHD | N/A | 1.1139 |  |  |

### Clinical relevance of A2G2S2 glycan with baseline features

Baseline levels of PV and CS A2G2S2 were positively correlated with BMI but were independent of age (**Table 13**). Analysis of routinely measured circulating blood markers found that PV CRP levels in patients positively correlated with CS A2G2S2 (*R* = 0.4320; *P* = 0.0275), but not with PV A2G2S2 (*r* = 0.1017; *P* = 0.3663) (**Table 13**). ROC curve analysis on CRP levels for discriminating AF patients from the SR controls found an averaged AUROC value of 0.5498 (± 0.1272 SD), indicating poor predictive performance for AF. No other circulating markers showed correlations with A2G2S2 levels. We also examined potential associations between baseline A2G2S2 baseline levels and ECG parameters (**Table 13**). The analysis found a negative correlation between QT interval and CS A2G2S2 (*r* = -0.3332; *P* = 0.0311). In contrast, this association was not significant for PV A2G2S2 (*r* = -0.1495; *P* = 0.1619), nor were any correlations detected between PV A2G2S2 and other ECG measurements.

**Table 13:**
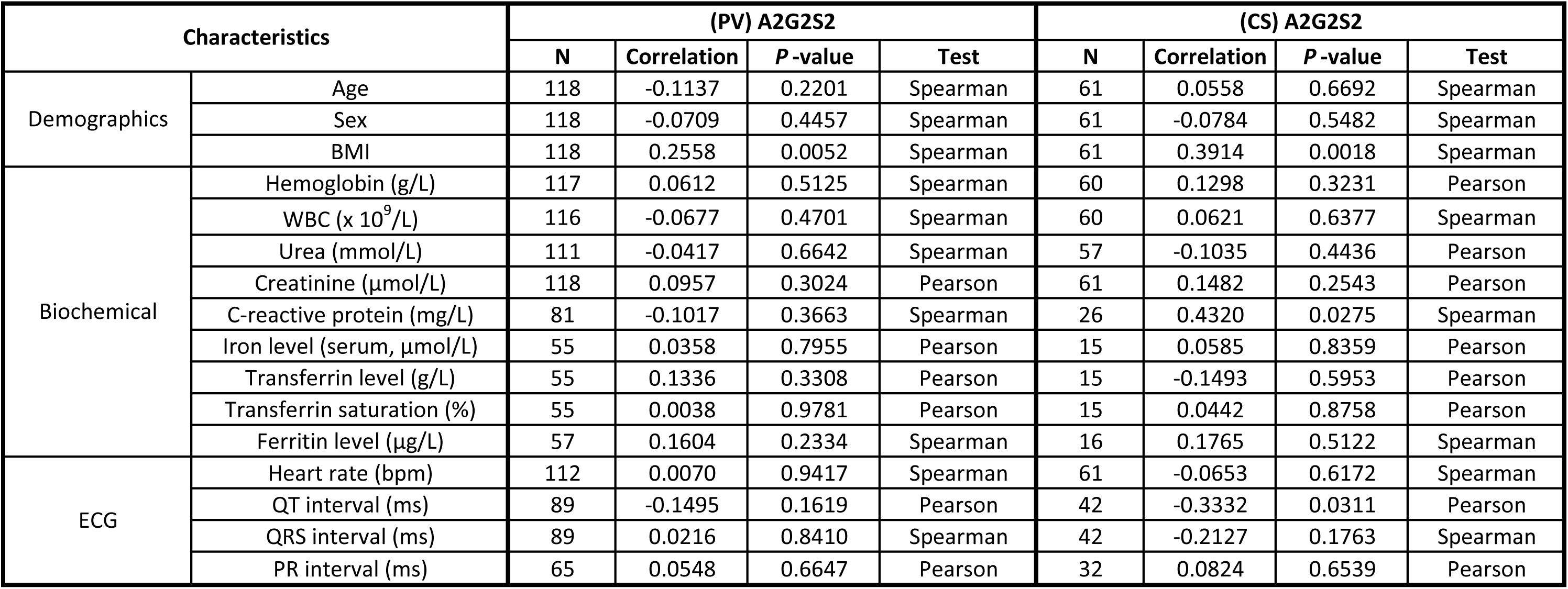
Baseline A2G2S2 glycan correlation with demographics, biochemical and ECG measurements. BMI, body mass index; CS, coronary sinus; ECG, Electrocardiogram; PV, peripheral vein; WBC, white blood cell

We investigated if A2G2S2 glycan levels were associated with the echocardiographic parameters of cardiac remodeling. Left atrial enlargement (LAE), commonly observed in AF (29), is determined by left atrial diameter ≥ 4.2 cm (men) or ≥ 4.0 cm (women) (30), or left atrial volume > 38 ml/m^2^ (31), or dilated left atria annotated in echocardiography report. Patients with LAE had increased CS (but not PV) A2G2S2 levels (*P* = 0.0354) (**Fig. 5a**) in the absence of correlation with the left atrial dimensions (*r* = 0.2905, *P* = 0.2276) (**Fig. 5b**). In contrast to the LAE, both PV and CS levels of A2G2S2 were not altered in patients with impaired LVEF (< 50%) (**Fig. 5c**) or left ventricular hypertrophy (LVH) (**Fig. 5d**). LVH is defined as indexed left ventricular mass ≥ 111 g/m^2^ (men) or ≥ 100 g/m^2^ (women) (31). These findings suggest that the elevated levels of CS A2G2S2 may be more representative of the remodeling in the atria rather than ventricle. Following this, we identified that patients who experienced AF for over a year have higher PV A2G2S2 levels compared with those who were in AF for shorter, less than 12 months, period (**Fig. 5e**); this pattern was not observed for the CS A2G2S2 abundance (**Fig. 5f**).

**Figure 5.**
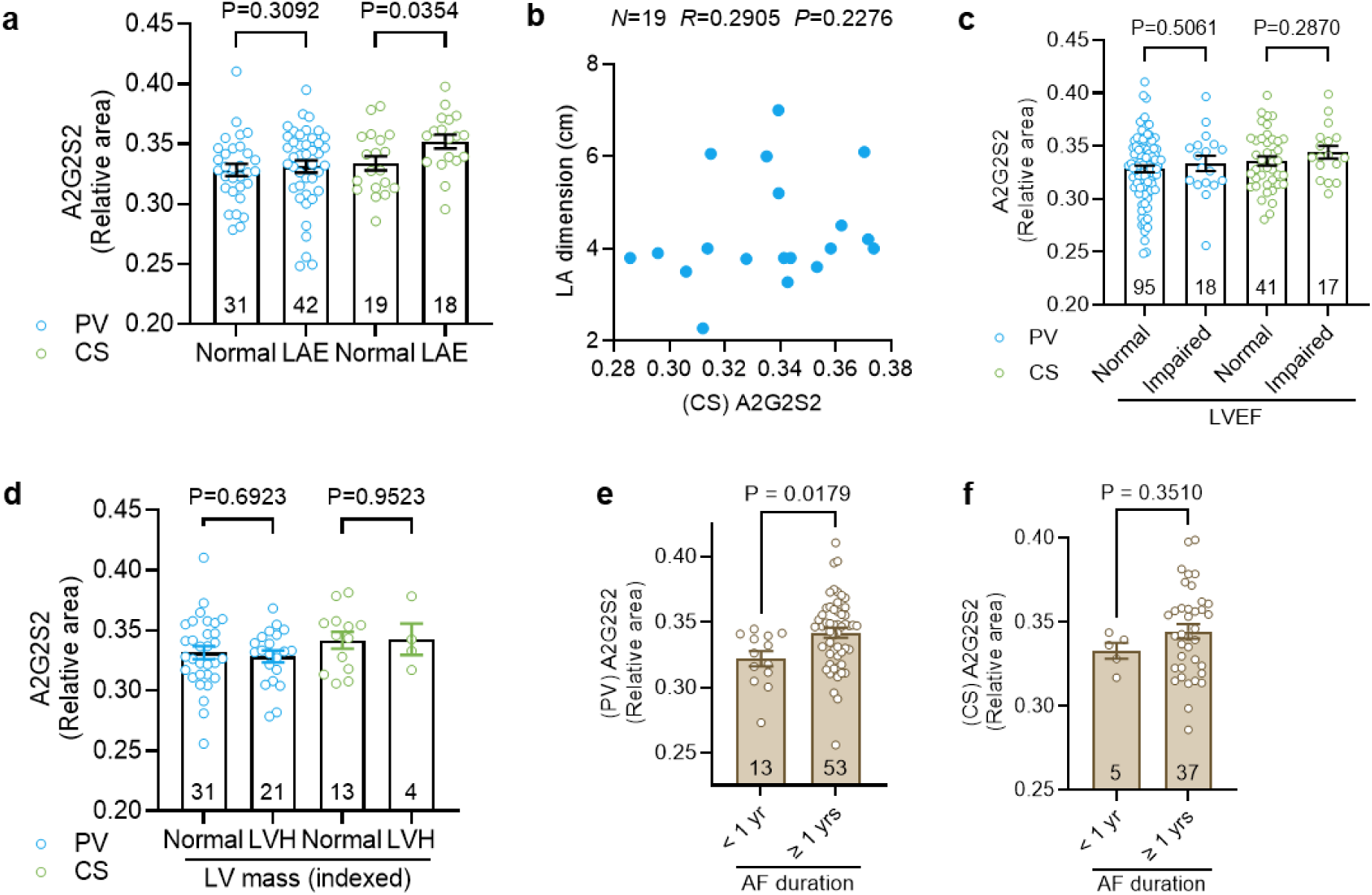
Associations of A2G2S2 levels with cardiac parameters. (**a**) A2G2S2 levels between patients with normal and enlarged left atria (LAE). (**b**) Correlation between CS A2G2S2 levels and LA dimension. (**c**) A2G2S2 levels between patients with normal and impaired LVEF. (**d**) A2G2S2 levels between patients with normal and hypertrophic left ventricle LVH, assessed by indexed LV mass. (**e-f**) Differences in A2G2S2 levels in AF patients with AF duration longer or shorter than one year. Data are expressed as mean ± SEM (a, c-f); N represents individual biological donors. *P* values are determined by Mann-Whitney test (a blue), unpaired t-test (a green, c-f) and Spearman’s correlation test (b). AF, atrial fibrillation; CS, coronary sinus; LA, left atrial; LAE, left atrial enlargement; LV, left ventricular; LVEF, left ventricular ejection fraction; PV, peripheral venous.

### Post-procedural and long-term changes of the circulating and cardiac A2G2S2 levels

In the cardiac surgery cohort, we found that the PV A2G2S2 was elevated on day 2 post-surgery (compared to the baseline levels) and remained elevated on day 4 (**Supplementary Fig. 4a**). In the ablation cohort, the A2G2S2 levels 2 hours post-procedure were significantly reduced in CS serum (*P* = 0.0010), but unchanged in PV serum (*P* = 0.3173) (**Supplementary Fig. 4b**). This suggests that ablation procedure may suppress local cardiac levels of A2G2S2, which, within the 2-hour period between sampling, is unlikely to exert an effect on A2G2S2 levels in the much larger peripheral serum pool.

We followed-up A2G2S2 abundance in the PV serum of AF patients at 3- and 12-months post-ablation. No differences were observed in PV A2G2S2 between patients who relapsed into AF or remained in SR after successful ablation procedure (**Supplementary Fig. 4c-4d**). PV A2G2S2 levels at 3-month post-ablation were not significantly altered in paroxysmal AF (*P* = 0.2462) and persistent AF patients (*P* = 0.5882) compared to the pre-ablation levels (**Supplementary Fig. 4e**). Similar findings were observed in persistent AF patients at the 12-month follow-up (*P* = 0.9624), whereas a trend toward a reduced PV A2G2S2 levels was noted in patients with paroxysmal AF (*P* = 0.0730) (**Supplementary Fig. 4f**).

## Discussion

To our knowledge, this is the first study to comprehensively profile the serum N-glycome from both peripheral venous and coronary sinus sources in patients with AF, highlighting novel cardiac and systemic N-glycosylation signatures linked to atrial remodeling. We identified individual and composite *N*-glycan traits associated with AF and further explored their clinical relevance by integrating clinical parameters into the analysis, including measures related to atrial remodeling.

Post-translational *N*-glycosylation has been linked to a variety of immuno-inflammatory diseases, including CVDs (23). Two case-control cardiovascular studies found an association of IgG *N*-glycan profile, agalactosylated and asialylation traits with increased incidence of CVD. Additionally, sialylation of glycoproteins is identified as an important predictor of atherosclerotic diseases (32). Furthermore, the inhibitors of the sialic acid regulating enzymes are in development for the treatment of atherosclerosis (32). Here, we found that AF patients display several upregulated derived traits, mainly composed of diantennary and tetrantennary glycans with either terminating galactose or sialic acid (S2, G2, S4, G4). Glycosylation involving these traits is highly associated with acute phase proteins such as alpha-1-acid glycoprotein and hemopexin (33). These proteins were upregulated in inflammation (34,35), one of the key pathogenetic mechanisms underlying and triggering AF (7), which may partially explain the elevation of galactosylated and sialylated traits in the serum *N*-glycome in patients with AF.

We established two unique glycan scores for AF classification in the PV and CS datasets. This represents the first composite serum *N*-glycan-based scoring approach for AF classification. Our findings indicate that coordinated alterations across multiple N-glycans, rather than individual glycans, reflect AF pathology and may provide a foundation for novel biomarker-based patient stratification strategies. A Similar strategy of generating a glycan score has previously been applied using IgG glycans to predict CVD risk (23). Notably, the glycans retained in the PV and CS glycan score models were not identical and no individual glycan was shared between the two models. This suggests that *N*-glycosylation changes in AF may differ between the serum source. Furthermore, it implies that *N*-glycosylation-mediated modulation of protein function contributing to the proarrhythmic phenotype may operate differently at the systemic level compared with the local cardiac environment. These findings underscore the importance of assessing *N*-glycans from both circulating and cardiac serum to uncover novel biological insights and improve AF prediction models. The translational potential of the proposed glycan scores would be further strengthened through validation in larger AF biomarker discovery cohorts.

To broaden the biological insights of systemic glycosylation alterations related to AF, we conducted a sub-analysis on 37 patients to correlate the PV glycan score with plasma protein levels detected from the same patient. The biological relevance of plasma proteins should be carefully considered, as many plasma glycoproteins, such as fibrinogen and Von Willebrand Factor (vWF), are established modulators of AF risk (36). These glycoproteins play crucial roles in coagulation, driving clot formation and increasing risk of thrombosis (37), a common condition coexisting with AF (38). Site specific alterations in the *N*-glycosylation of fibrinogen were observed in plasma from patients with AF and strongly associated with cardiovascular risk factors (37). These changes may support fibrinogen glycosylation as a potential mechanistic link and biomarker in AF.

The positive correlations of the AF-associated PV glycan score with FURIN and BMP7 suggest that the composite glycan signature may reflect active atrial structural remodeling processes. FURIN plays a critical role in atrial fibrosis by acting as a proteolytic enzyme to cleave and activate transforming growth factor-beta (TGF-β), the master regulator of fibrosis (39). Although BMP7 demonstrates anti-fibrotic effects in AF by antagonising TGF-β signalling (40), its positive correlation with PV glycan score suggests that BMP7 glycosylation may influence its stability and potentially modulate signalling potency. Beyond structural remodeling, another important aspect of AF pathology is the thrombo-inflammatory state, as reflected by the association between with PV glycan score and the proteins C4BPB, IL17F and THPO. C4BPB is a heavily glycosylated protein involving in the inhibition of complement activation (41). Pro-inflammatory cytokines, like tumour necrosis factor (TNF) and interleukin-6 (IL6) are well established markers of AF (42). Plasma level of IL17F, a member of the Th17-related cytokines, is associated with increased AF risk (43) and contributes to chronic tissue inflammation in psoriatic arthritis (44). THPO is a key growth factor to stimulate the activation of circulating platelets (45), and modification by *N*- or *O*-glycosylation at the C-terminal of this protein has been demonstrated to regulate its half-life and stability (46). Collectively, the significantly correlated proteins may serve as biomarkers of specific *N*-glycosylation changes in AF; however, targeted follow-up studies are warranted to understand how glycosylation modulates their functions in AF, for example through site-specific glycoproteomics or functional studies.

Notably, the serum *N*-glycans outperformed CRP alone for the prediction of AF. This is consistent with previous reports demonstrating that *N*-glycans are superior to CRP markers for predicting CVD and IBD (21,22). Although CRP is well-recognised for predicting systemic inflammation and future CVD events (47,48), it is not a specific marker for AF, as elevation in CRP can be triggered by a wide range of conditions, in which infection is the main player (49). This highlights the potential of *N*-glycans as clinical markers for AF, as they are less reactive than the acute phase proteins like CRP and exhibit greater stability during chronic inflammation (50,51).

In this study, we found that the A2G2S2 *N*-glycan is highly abundant in both circulating PV and cardiac CS serum and significantly upregulated in patients with AF. A2G2S is a diantennary sialylated glycan; notably, increased sialylation has been associated with inflammation and advanced CVD (22). Our findings of the elevated serum A2G2S2 in AF are consistent with a previous report showing serum A2G2S2 levels in IgG *N*-glycome in AF patients (24). Consistent with previous evidence (52–54), we found that A2G2S2 is not only the most abundant and reliably detected glycan in human serum, but may also serve as a stable molecular signature reflecting atrial remodeling and inflammation in AF patients. Importantly, previous studies have shown that variations in total plasma *N*-glycans abundance are minimal over five days period in healthy individuals (55), with the lowest variability (7.5% coefficients of variations) found for a glycan group (GP9) containing A2G2S2 glycan (55). These properties emphasise the potential value of A2G2S2 glycan to serve as a reliable and stable molecular signature, including in the context of AF.

To test the potential association of A2G2S2 with cardiac electrical changes, prominent in AF, the ECG data showed limited relevance of serum A2G2S2 changes to cardiac electrical signalling, except for QT intervals which may require further evaluation. The CS, but not PV, levels of A2G2S2 glycan were raised in patients with LAE, indicating that changes in A2G2S2 may reflect the left atrial remodeling. Left atrial diameter alone is shown to have poorer performance in predicting cardiovascular events compared to indexed left atrial volume (56), which may explain why CS A2G2S2 was not correlated with left atrial dimension, while being positively associated with patients with LAE. A less profound atrial structural remodeling observed in patients with arrhythmias, such as SVTs and flutters (assigned to the non-AF group in this study), compared to those with AF (57) may potentially account for the CS A2G2S2 levels in AF group compared to the non-AF group.

The acute inflammatory response, accompanied by oxidative stress and an increased cytokine production, is commonly observed post-operatively in patients undergoing major, including open-heart, surgeries (58). Post-cardiac surgery levels of PV A2G2S2 were significantly elevated and sustained for four days, indicating that this might be triggered by the pro-inflammatory responses inflicted by major surgical trauma. By contrast, patients with ablation displayed reduced A2G2S2 levels in CS serum 2 hours post-procedure in the absence of changes in PV serum. Ablation, via ceasing the aberrant atrial electrical activity, may be sufficient to suppress the release of cardiac-derived A2G2S2 into the CS pool, with no impact, due to its localised (atrial, around pulmonary veins) nature, on the total circulating glycan levels. In contrast to the longer duration post-cardiac surgery (two to four days) for serum sampling, blood samples from the ablation cohort were collected 2 hours post-procedure which may be too soon for any detectable, inflammation-driven upregulation of A2G2S2 levels. However, these mechanistic links are yet to be explored.

While ablation offers an effective therapy for patients with paroxysmal and symptomatic persistent AF, AF recurrence after restoration of SR with ablation remains common and limits its efficacy (59,60). Interestingly, a recent study reported a reduction in A2G2S2 in IgG *N*-glycome 6 months post-ablation (24). Similarly, in our cohort, there was a reducing trend in PV A2G2S2 levels at 12-month post-ablation in paroxysmal AF patients with no changes observed at 3-month follow-up (relative to baseline). These results indicate that changes in this glycan might be time-dependent in patients with AF and may reflect the differences in the arrhythmogenic substrate between paroxysmal (short, self-terminating) and persistent (chronic, not self-terminating) AF. It is conceivable that a more complex atrial remodeling in persistent AF may account for the non-significant changes in A2G2S2 in peripheral circulation after ablation, in contrast to a reducing trend in paroxysmal AF typically associated with a less advanced atrial remodeling and better outcomes at 12-month follow-up.

It is important to mention that although the SR control patients had no AF history, they expectedly (i.e., indications for cardiac surgery) had other CVDs or conditions, such as IHD, myocardial infarction and chronic kidney disease. Nonetheless, we accounted for these confounding factors in the regression analyses. A further validation of the current findings in larger cohorts and in serum samples obtained from healthier controls would be desirable to aid a better understanding of the role of *N*-glycans in AF. While the multivariate glycan scores demonstrated strong discriminative performance in AF, integrating them with additional biomarkers or clinical parameters in future studies could further improve their predictive power and provide deeper insights into their biological roles of *N*-glycans in AF. A better understanding of the glycoprotein biology alongside the *N*-glycosylation profiling, e.g., through glycoproteomics, could further advance our knowledge of the biological processes contributing to AF pathogenesis. Previous studies identified acute phase proteins mainly contributing to *the N*-glycans in human plasma and serum (33). These proteins play a prominent biological role in inflammation, and could, in conjunction with other inflammatory markers, be explored further in AF.

In summary, we profiled cardiac and circulating serum *N*-glycosylation patterns in AF patients at three levels: individual glycan, grouped glycans (derived traits) and composite glycan scores. This study extends previous characterisation of the *N*-glycome in AF, particularly highlighting dysregulated cardiac *N*-glycan signatures. Validation in larger cohorts will be critical to confirm these N-glycan signatures as mechanistic markers of AF and explore their potential for improving patient stratification, risk prediction, and development of targeted therapies.

## Acknowledgements

C.H.K.Y., J.C., D.I.R.S. and S.R. conceived and designed the analyses. Patient consent was obtained by C.S.M., L.M.M., A.M.J., N.M. and K.C.; human blood samples were collected by the clinicians T.R.B., K.R., M.G., M.P., Y.B., R.W., R.S., G.K., V.S. and A.K. under ethical approval granted to S.R.; C.H.K.Y., C.S.M., L.M.M., A.M.J., N.M. and K.C. processed serum samples. C.H.K.Y carried out the selection of samples with the assistance of C.S.M. and A.M.J.; J.C. was responsible for the sample processing for *N*-glycome profiling. J.C. and G.EH. contributed to data processing. C.H.K.Y., G.EH. and A.B. were responsible for statistical analysis. S.R. secured funding. S.R. and D.I.R.S. supervised the work. C.H.K.Y., J.C., G.EH., C.S.M., D.I.R.S. and S.R. wrote and edited the manuscript. We are grateful for all participants who provided the blood samples, and all staff (the nurse teams) and investigators who contributed to this study. We thank the Multiomics Technology Platforms Group at the Centre of Human Genetics for the generation and initial processing of the sequencing data.

## Funding

This work was funded by the British Heart Foundation (BHF) Intermediate (FS/SBSRF/22/31026) and Senior (FS/SBSRF/22/31033) Fellowships (to S.R.), Oxford BHF Centre of Research Excellence (CRE) grants (to S.R.), the British Research Council (BRC4) NIHR Oxford Biomedical Research Centre grant (to S.R. and C.H.K.Y.), the BHF Clinical Research Training Fellowship (FS/CRTF/25/24786; to C.S.M.), the BHF PhD Studentship (FS/20/7/34992; to L.M.M.), the BHF Studentship grant (FS/4yPhD/F/22/34177; to A.M.J.), and The Royal Society University Research Fellowship (URF\R1\221314; to A.B.).

## Disclosure of interest

J.C., G.EH. and D.I.R.S. are employed by Ludger Ltd, a company that commercialises glycan analysis tools and services. The remaining authors declare no competing interests.

## Data availability statement

All data related to the manuscript is included in the manuscript. The metadata of the patient cohorts that support the findings are available from the first author (C.H.K.Y.) and corresponding author (S.R.) upon request.

## Notes

### Competing Interest Statement

The authors have declared no competing interest.

### Author Declarations

This study was approved by the South Central-Berkshire B Research Ethics Committee (UK, 18/SC/0404 and 18/SC/0304) at John Radcliffe Hospital, Oxford.

## References

1. Roth GA, Mensah GA, Johnson CO, Addolorato G, Ammirati E, Baddour LM, et al. Global Burden of Cardiovascular Diseases and Risk Factors, 1990–2019: Update From the GBD 2019 Study. J Am Coll Cardiol [Internet]. 2020 Dec 22 [cited 2025 Jun 7];76(25):2982. Available from: https://pmc.ncbi.nlm.nih.gov/articles/PMC7755038/

2. Odutayo A, Wong CX, Hsiao AJ, Hopewell S, Altman DG, Emdin CA. Atrial fibrillation and risks of cardiovascular disease, renal disease, and death: systematic review and meta-analysis. BMJ [Internet]. 2016 Sep 6 [cited 2025 Jun 13];354:i4482. Available from: https://www.bmj.com/content/354/bmj.i4482

3. Koh YH, Lew LZW, Franke KB, Elliott AD, Lau DH, Thiyagarajah A, et al. Predictive role of atrial fibrillation in cognitive decline: a systematic review and meta-analysis of 2.8 million individuals. EP Europace [Internet]. 2022 Sep 1 [cited 2025 Jun 7];24(8):1229–39. Available from: 10.1093/europace/euac003

4. Zhang W, Liang J, Li C, Gao D, Ma Q, Pan Y, et al. Age at Diagnosis of Atrial Fibrillation and Incident Dementia. JAMA Netw Open [Internet]. 2023 Nov 1 [cited 2025 Jun 7];6(11):e2342744–e2342744. Available from: https://jamanetwork.com/journals/jamanetworkopen/fullarticle/2811523

5. Chen Q, Yi Z, Cheng J. Atrial fibrillation in aging population. Aging Medicine [Internet]. 2018 Jan 1 [cited 2025 Jun 7];1(1):67–74. Available from: /doi/pdf/10.1002/agm2.12015

6. Middeldorp ME, Sandhu RK, Mao J, Gencer B, Danik JS, Moorthy V, et al. Risk Factors for the Development of New-Onset Persistent Atrial Fibrillation: Subanalysis of the VITAL Study. Circ Arrhythm Electrophysiol [Internet]. 2023 Dec 1 [cited 2025 Jun 7];16(12):651–62. Available from: /doi/pdf/10.1161/CIRCEP.123.012334?download=true

7. Sagris M, Vardas EP, Theofilis P, Antonopoulos AS, Oikonomou E, Tousoulis D. Atrial Fibrillation: Pathogenesis, Predisposing Factors, and Genetics. International Journal of Molecular Sciences 2022, Vol 23, Page 6 [Internet]. 2021 Dec 21 [cited 2025 Jun 7];23(1):6. Available from: https://www.mdpi.com/1422-0067/23/1/6/htm

8. Dzeshka MS, Lip GYH, Snezhitskiy V, Shantsila E. Cardiac Fibrosis in Patients With Atrial Fibrillation: Mechanisms and Clinical Implications. J Am Coll Cardiol [Internet]. 2015 Aug 25 [cited 2025 Jun 7];66(8):943–59. Available from: /doi/pdf/10.1016/j.jacc.2015.06.1313?download=true

9. McGann C, Akoum N, Patel A, Kholmovski E, Revelo P, Damal K, et al. Atrial fibrillation ablation outcome is predicted by left atrial remodeling on MRI. Circ Arrhythm Electrophysiol [Internet]. 2014 [cited 2025 Jun 13];7(1):23–30. Available from: /doi/pdf/10.1161/CIRCEP.113.000689?download=true

10. Clift CL, Mehta A, Drake RR, Angel PM. Multiplexed Imaging Mass Spectrometry of Histological Staining, N-Glycan and Extracellular Matrix from One Tissue Section: A Tool for Fibrosis Research. Methods in Molecular Biology [Internet]. 2021 [cited 2025 Jun 7];2350:313–29. Available from: https://link.springer.com/protocol/10.1007/978-1-0716-1593-5_20

11. Reily C, Stewart TJ, Renfrow MB, Novak J. Glycosylation in health and disease. Nature Reviews Nephrology 2019 15:6 [Internet]. 2019 Mar 11 [cited 2025 Jun 7];15(6):346–66. Available from: https://www.nature.com/articles/s41581-019-0129-4

12. Radovani B, Gudelj I. N-Glycosylation and Inflammation; the Not-So-Sweet Relation. Front Immunol [Internet]. 2022 Jun 27 [cited 2025 Jun 7];13:893365. Available from: www.frontiersin.org

13. Tian M, Li X, Yu L, Qian JX, Bai XY, Yang J, et al. Glycosylation as an intricate post-translational modification process takes part in glycoproteins related immunity. Cell Communication and Signaling 2025 23:1 [Internet]. 2025 May 5 [cited 2025 Jun 7];23(1):1–18. Available from: https://biosignaling.biomedcentral.com/articles/10.1186/s12964-025-02216-w

14. Pinho SS, Alves I, Gaifem J, Rabinovich GA. Immune regulatory networks coordinated by glycans and glycan-binding proteins in autoimmunity and infection. Cellular & Molecular Immunology 2023 20:10 [Internet]. 2023 Aug 15 [cited 2025 Jun 7];20(10):1101–13. Available from: https://www.nature.com/articles/s41423-023-01074-1

15. Adua E, Afrifa-Yamoah E, Peprah-Yamoah E, Anto EO, Acheampong E, Awuah-Mensah KA, et al. Multi-block data integration analysis for identifying and validating targeted N-glycans as biomarkers for type II diabetes mellitus. Sci Rep [Internet]. 2022 Dec 1 [cited 2025 Jun 7];12(1):1–12. Available from: https://www.nature.com/articles/s41598-022-15172-z

16. Rudman N, Gornik O, Lauc G. Altered N-glycosylation profiles as potential biomarkers and drug targets in diabetes. FEBS Lett [Internet]. 2019 Jul 1 [cited 2025 Jun 7];593(13):1598–615. Available from: /doi/pdf/10.1002/1873-3468.13495

17. Ruhaak LR, Miyamoto S, Lebrilla CB. Developments in the identification of glycan biomarkers for the detection of cancer. Molecular and Cellular Proteomics [Internet]. 2013 Apr 1 [cited 2025 Jun 7];12(4):846–55. Available from: https://www.mcponline.org/action/showFullText?pii=S1535947620311294

18. Cao X, Shang QH, Chi XL, Zhang W, Xiao HM, Sun MM, et al. Serum N-glycan markers for diagnosing liver fibrosis induced by hepatitis B virus. World J Gastroenterol [Internet]. 2020 Mar 14 [cited 2025 Jun 7];26(10):1067–79. Available from: https://www.wjgnet.com/1007-9327/full/v26/i10/1067.htm

19. Hanamatsu H, Suda G, Ohara M, Ogawa K, Tamaki N, Hikita H, et al. Elevated A2F bisect N-glycans of serum IgA reflect progression of liver fibrosis in patients with MASLD. J Gastroenterol [Internet]. 2025 Apr 1 [cited 2025 Jun 7];60(4):456–68. Available from: https://link.springer.com/article/10.1007/s00535-024-02206-8

20. Wittenbecher C, Štambuk T, Kuxhaus O, Rudman N, Vučković F, Štambuk J, et al. Plasma N-Glycans as Emerging Biomarkers of Cardiometabolic Risk: A Prospective Investigation in the EPIC-Potsdam Cohort Study. Diabetes Care. 2020 Mar 1;43(3):661–8.

21. Shubhakar A, Jansen BC, Adams AT, Reiding KR, Ventham NT, Kalla R, et al. Serum N-Glycomic Biomarkers Predict Treatment Escalation in Inflammatory Bowel Disease. J Crohns Colitis [Internet]. 2023 Jun 16 [cited 2025 Jun 7];17(6):919–32. Available from: 10.1093/ecco-jcc/jjad012

22. Cheeseman J, Badia C, Elgood-Hunt G, Gardner RA, Trinh DN, Monopoli MP, et al. Elevated concentrations of Neu5Ac and Neu5,9Ac2 in human plasma: potential biomarkers of cardiovascular disease. Glycoconj J [Internet]. 2023 Dec 1 [cited 2025 Jun 7];40(6):645–54. Available from: https://link.springer.com/article/10.1007/s10719-023-10138-3

23. Hoshi RA, Plavša B, Liu Y, Trbojević-Akmačić I, Glynn RJ, Ridker PM, et al. N-Glycosylation Profiles of Immunoglobulin G and Future Cardiovascular Events. Circ Res [Internet]. 2024 Mar 1 [cited 2025 Jun 7];134(5):E3–14. Available from: /doi/pdf/10.1161/CIRCRESAHA.123.323623?download=true

24. Plavša B, Szavits-Nossan J, Blivajs A, Rapčan B, Radovani B, Šesto I, et al. The N-Glycosylation of Total Plasma Proteins and IgG in Atrial Fibrillation. Biomolecules [Internet]. 2023 Apr 1 [cited 2025 Jun 7];13(4):605. Available from: https://www.mdpi.com/2218-273X/13/4/605/htm

25. Jansen BC, Hafkenscheid L, Bondt A, Gardner RA, Hendel JL, Wuhrer M, et al. HappyTools: A software for high-throughput HPLC data processing and quantitation. PLoS One [Internet]. 2018 Jul 1 [cited 2025 Jun 7];13(7):e0200280. Available from: https://journals.plos.org/plosone/article?id=10.1371/journal.pone.0200280

26. Saldova R, Asadi Shehni A, Haakensen VD, Steinfeld I, Hilliard M, Kifer I, et al. Association of N-glycosylation with breast carcinoma and systemic features using high-resolution quantitative UPLC. J Proteome Res [Internet]. 2014 May 2 [cited 2025 Jun 7];13(5):2314–27. Available from: /doi/pdf/10.1021/pr401092y

27. Varki A, Cummings RD, Aebi M, Packer NH, Seeberger PH, Esko JD, et al. Symbol Nomenclature for Graphical Representations of Glycans. Glycobiology [Internet]. 2015 Dec 1 [cited 2025 Jun 7];25(12):1323–4. Available from: 10.1093/glycob/cwv091

28. Wehrens R, Hageman JA, van Eeuwijk F, Kooke R, Flood PJ, Wijnker E, et al. Improved batch correction in untargeted MS-based metabolomics. Metabolomics [Internet]. 2016 May 1 [cited 2025 Jun 7];12(5):1–12. Available from: https://link.springer.com/article/10.1007/s11306-016-1015-8

29. Kirchhof P, Benussi S, Kotecha D, Ahlsson A, Atar D, Casadei B, et al. 2016 ESC Guidelines for the management of atrial fibrillation developed in collaboration with EACTS. Eur Heart J [Internet]. 2016 Oct 7 [cited 2025 Jun 7];37(38):2893–962. Available from: 10.1093/eurheartj/ehw210

30. Bouzas-Mosquera A, Broullón FJ, Álvarez-García N, Méndez E, Peteiro J, Gándara-Sambade T, et al. Left atrial size and risk for all-cause mortality and ischemic stroke. CMAJ Canadian Medical Association Journal [Internet]. 2011 Jul 12 [cited 2025 Jun 13];183(10):E657–64. Available from: https://www.cmaj.ca/content/183/10/E657

31. Harkness A, Ring L, Augustine DX, Oxborough D, Robinson S, Sharma V. Normal reference intervals for cardiac dimensions and function for use in echocardiographic practice: A guideline from the British Society of Echocardiography. Echo Res Pract [Internet]. 2020 Mar 1 [cited 2025 Jun 13];7(1):G1–18. Available from: https://echo.biomedcentral.com/articles/10.1530/ERP-19-0050

32. Wattchow NE, Pullen BJ, Indraratna AD, Nankivell V, Everest-Dass A, Psaltis PJ, et al. The emerging role of glycans and the importance of sialylation in cardiovascular disease. Atherosclerosis [Internet]. 2025 Apr 1 [cited 2025 Jun 7];403:119172. Available from: https://www.atherosclerosis-journal.com/action/showFullText?pii=S002191502500070X

33. Clerc F, Reiding KR, Jansen BC, Kammeijer GSM, Bondt A, Wuhrer M. Human plasma protein N-glycosylation. Glycoconjugate Journal 2016 33:3 [Internet]. 2015 Nov 10 [cited 2025 Jun 7];33(3):309–43. Available from: https://link.springer.com/article/10.1007/s10719-015-9626-2

34. Tolosano E, Altruda F. Hemopexin: Structure, function, and regulation. DNA Cell Biol [Internet]. 2002 Jul 6 [cited 2025 Jun 7];21(4):297–306. Available from: /doi/pdf/10.1089/104454902753759717?download=true

35. Blain P, Mucklow J, Rawlins M, Roberts D, Routledge P, Shand D. Determinants of plasma alpha 1-acid glycoprotein (AAG) concentrations in health. Br J Clin Pharmacol [Internet]. 1985 Nov 1 [cited 2025 Jun 7];20(5):500–2. Available from: /doi/pdf/10.1111/j.1365-2125.1985.tb05107.x

36. Rafaqat S, Gluscevic S, Patoulias D, Sharif S, Klisic A. The Association between Coagulation and Atrial Fibrillation. Biomedicines [Internet]. 2024 Feb 1 [cited 2025 Dec 17];12(2):274. Available from: https://pmc.ncbi.nlm.nih.gov/articles/PMC10887311/

37. Šoić D, Kifer D, Szavits-Nossan J, Blivajs A, Đerek L, Rudan D, et al. High-Throughput Site-Specific N-Glycosylation Profiling of Human Fibrinogen in Atrial Fibrillation. J Proteome Res [Internet]. 2025 Apr 4 [cited 2025 Dec 17];24(4):2121–34. Available from: /doi/pdf/10.1021/acs.jproteome.5c00096?ref=article_openPDF

38. Lutsey PL, Norby FL, Alonso A, Cushman M, Chen LY, Michos ED, et al. Atrial fibrillation and venous thromboembolism: evidence of bidirectionality in the Atherosclerosis Risk in Communities Study. J Thromb Haemost [Internet]. 2018 Apr 1 [cited 2025 Dec 17];16(4):670. Available from: https://pmc.ncbi.nlm.nih.gov/articles/PMC5893387/

39. Chen C, Chen P, Yu W, Zhao L, Yang Y, Qu H, et al. TGF-β-Driven Atrial Fibrosis in Atrial Fibrillation: From Mechanistic Insights to Targeted Therapies. Aging Dis [Internet]. 2025 Jul 31 [cited 2025 Dec 17];0-. Available from: https://www.aginganddisease.org/EN/10.14336/AD.2025.0564

40. Chen X, Xu J, Jiang B, Liu D. Bone Morphogenetic Protein-7 Antagonizes Myocardial Fibrosis Induced by Atrial Fibrillation by Restraining Transforming Growth Factor-β (TGF-β)/Smads Signaling. Med Sci Monit [Internet]. 2016 Sep 28 [cited 2025 Dec 17];22:3457. Available from: https://pmc.ncbi.nlm.nih.gov/articles/PMC5045133/

41. Kadavá T, Hevler JF, Kalaidopoulou Nteak S, Yin VC, Strasser J, Preiner J, et al. Higher-order structure and proteoforms of co-occurring C4b-binding protein assemblies in human serum. EMBO J [Internet]. 2024 Jul 15 [cited 2025 Dec 17];43(14):3009. Available from: https://pmc.ncbi.nlm.nih.gov/articles/PMC11251186/

42. Ihara K, Sasano T. Role of Inflammation in the Pathogenesis of Atrial Fibrillation. Front Physiol [Internet]. 2022 Apr 14 [cited 2025 Dec 17];13:862164. Available from: www.frontiersin.org

43. Wu N, Xu B, Liu Y, Chen X, Tang H, Wu L, et al. Elevated plasma levels of Th17-related cytokines are associated with increased risk of atrial fibrillation. Scientific Reports 2016 6:1 [Internet]. 2016 May 20 [cited 2025 Dec 17];6(1):26543-. Available from: https://www.nature.com/articles/srep26543

44. Glatt S, Baeten D, Baker T, Griffiths M, Ionescu L, Lawson ADG, et al. Dual IL-17A and IL-17F neutralisation by bimekizumab in psoriatic arthritis: Evidence from preclinical experiments and a randomised placebo-controlled clinical trial that IL-17F contributes to human chronic tissue inflammation. Ann Rheum Dis [Internet]. 2018 Apr 1 [cited 2025 Dec 17];77(4):523–32. Available from: https://ard.eular.org/action/showFullText?pii=S0003496724009324

45. Lupia E, Capuano M, Vizio B, Schiavello M, Bosco O, Gelardi M, et al. Thrombopoietin participates in platelet activation in COVID-19 patients. EBioMedicine [Internet]. 2022 Nov 1 [cited 2025 Dec 17];85:104305. Available from: https://pmc.ncbi.nlm.nih.gov/articles/PMC9556163/

46. Sarson-Lawrence KTG, Hardy JM, Iaria J, Stockwell D, Behrens K, Saiyed T, et al. Cryo-EM structure of the extracellular domain of murine Thrombopoietin Receptor in complex with Thrombopoietin. Nature Communications 2024 15:1 [Internet]. 2024 Feb 7 [cited 2025 Dec 17];15(1):1135-. Available from: https://www.nature.com/articles/s41467-024-45356-2

47. Dorresteijn JAN, Visseren FLJ, Wassink AMJ, Gondrie MJA, Steyerberg EW, Ridker PM, et al. Development and validation of a prediction rule for recurrent vascular events based on a cohort study of patients with arterial disease: the SMART risk score. Heart [Internet]. 2013 Jun 15 [cited 2025 Jun 7];99(12):866–72. Available from: https://heart.bmj.com/content/99/12/866

48. Ridker PM, Bhatt DL, Pradhan AD, Glynn RJ, MacFadyen JG, Nissen SE. Inflammation and cholesterol as predictors of cardiovascular events among patients receiving statin therapy: a collaborative analysis of three randomised trials. The Lancet [Internet]. 2023 Apr 15 [cited 2025 Jun 7];401(10384):1293–301. Available from: https://www.thelancet.com/action/showFullText?pii=S0140673623002155

49. Landry A, Docherty P, Ouellette S, Cartier LJ. Causes and outcomes of markedly elevated C-reactive protein levels. Canadian Family Physician [Internet]. 2017 Jun 1 [cited 2025 Jun 7];63(6):e316. Available from: https://pmc.ncbi.nlm.nih.gov/articles/PMC5471098/

50. Levine JA, Han JM, Wolska A, Wilson SR, Patel TP, Remaley AT, et al. Associations of GlycA and high-sensitivity C-reactive protein with measures of lipolysis in adults with obesity. J Clin Lipidol [Internet]. 2020 Sep 1 [cited 2025 Jun 7];14(5):667–74. Available from: https://www.lipidjournal.com/action/showFullText?pii=S1933287420302221

51. Crick DCP, Khandaker GM, Halligan SL, Burgner D, Mansell T, Fraser A. Comparison of the stability of glycoprotein acetyls and high sensitivity C-reactive protein as markers of chronic inflammation. Immunology [Internet]. 2024 Apr 1 [cited 2025 Jun 7];171(4):497–512. Available from: /doi/pdf/10.1111/imm.13739

52. Colombo M, Shehni AA, Thoma I, McGurnaghan SJ, Blackbourn LAK, Wilkinson H, et al. Quantitative levels of serum N-glycans in type 1 diabetes and their association with kidney disease. Glycobiology [Internet]. 2021 Jun 3 [cited 2025 Jun 7];31(5):613–23. Available from: 10.1093/glycob/cwaa106

53. Gebrehiwot AG, Melka DS, Kassaye YM, Rehan IF, Rangappa S, Hinou H, et al. Healthy human serum N-glycan profiling reveals the influence of ethnic variation on the identified cancer-relevant glycan biomarkers. PLoS One [Internet]. 2018 Dec 1 [cited 2025 Jun 7];13(12):e0209515. Available from: https://journals.plos.org/plosone/article?id=10.1371/journal.pone.0209515

54. Kita Y, Miura Y, Furukawa JI, Nakano M, Shinohara Y, Ohno M, et al. Quantitative glycomics of human whole serum glycoproteins based on the standardized protocol for liberating N-glycans. Molecular and Cellular Proteomics [Internet]. 2007 Aug 1 [cited 2025 Jun 7];6(8):1437–45. Available from: https://www.mcponline.org/action/showFullText?pii=S1535947620321617

55. Gornik O, Wagner J, Pučić M, Knežević A, Redžić I, Lauc G. Stability of N-glycan profiles in human plasma. Glycobiology [Internet]. 2009 Dec 1 [cited 2025 Jun 7];19(12):1547–53. Available from: 10.1093/glycob/cwp134

56. Tsang TSM, Abhayaratna WP, Barnes ME, Miyasaka Y, Gersh BJ, Bailey KR, et al. Prediction of Cardiovascular Outcomes With Left Atrial Size: Is Volume Superior to Area or Diameter? J Am Coll Cardiol [Internet]. 2006 Mar 7 [cited 2025 Jun 17];47(5):1018–23. Available from: /doi/pdf/10.1016/j.jacc.2005.08.077?download=true

57. Guichard JB, Naud P, Xiong F, Qi X, L’Heureux N, Hiram R, et al. Comparison of Atrial Remodeling Caused by Sustained Atrial Flutter Versus Atrial Fibrillation. J Am Coll Cardiol [Internet]. 2020 Jul 28 [cited 2025 Jun 18];76(4):374–88. Available from: /doi/pdf/10.1016/j.jacc.2020.05.062?download=true

58. Suleiman MS, Zacharowski K, Angelini GD. Inflammatory response and cardioprotection during open-heart surgery: the importance of anaesthetics. Br J Pharmacol [Internet]. 2008 Jan 1 [cited 2025 Jun 7];153(1):21–33. Available from: /doi/pdf/10.1038/sj.bjp.0707526

59. Dretzke J, Chuchu N, Agarwal R, Herd C, Chua W, Fabritz L, et al. Predicting recurrent atrial fibrillation after catheter ablation: a systematic review of prognostic models. EP Europace [Internet]. 2020 May 1 [cited 2025 Jun 13];22(5):748–60. Available from: 10.1093/europace/euaa041

60. Choi SH, Yu HT, Kim D, Park JW, Kim TH, Uhm JS, et al. Late recurrence of atrial fibrillation 5 years after catheter ablation: predictors and outcome. EP Europace [Internet]. 2023 May 19 [cited 2025 Jun 13];25(5):1–10. Available from: 10.1093/europace/euad113

